# Household COVID-19 risk and in-person schooling

**DOI:** 10.1101/2021.02.27.21252597

**Authors:** Justin Lessler, M. Kate Grabowski, Kyra H. Grantz, Elena Badillo-Goicoechea, C. Jessica E. Metcalf, Carly Lupton-Smith, Andrew S. Azman, Elizabeth A. Stuart

## Abstract

In-person schooling has proved contentious and difficult to study throughout the SARS-CoV-2 pandemic. Data from a massive online survey in the United States indicates an increased risk of COVID-19-related outcomes among respondents living with a child attending school in-person. School-based mitigation measures are associated with significant reductions in risk, particularly daily symptoms screens, teacher masking, and closure of extra-curricular activities. With seven or more mitigation measures, the association between in-person schooling and COVID-19-related outcomes all but disappears. Teachers working outside the home were more likely to report COVID-19-related outcomes, but this association is similar to other occupations (e.g., healthcare, office work). In-person schooling is associated with household COVID-19 risk, but this risk can likely be controlled with properly implemented school-based mitigation measures.

**One sentence summary:** Living with children attending in-person school is linked to a higher risk of COVID-19 outcomes, which school-based interventions can mitigate.

## Main Text

The role of schools in transmission, and the value of school closure, has been one of the most contentious issues over the course of the COVID-19 pandemic. There is ongoing debate about exactly how much SARS-CoV-2 risk is posed to individuals and communities by in-person schooling. While there is general consensus that it should be possible to open schools safely with adequate mitigation measures, there is little data and even less agreement as to what level of mitigation is needed.

Many ecological studies have shown an association between in-person schooling and the speed and extent of community SARS-CoV-2 transmission (*1*–*3*), though these results have been far from uniform (*4*). While there have been numerous outbreaks of SARS-CoV-2 in schools and school-like settings (*5*–*7*), studies conducted outside of outbreak settings have suggested that, when mitigation measures are in place, transmission within schools is limited and infection rates mirror that of the surrounding community (*8*, *9*).

However, the ways in which in-person schooling influences community SARS-CoV-2 incidence are complex. Schools play a unique role in the social fabric of the United States and other countries, and often create potential transmission connections between otherwise disparate communities. Even if transmission in classrooms is rare, activities surrounding in-person schooling, such as student pick-up and drop-off, teacher interactions, and broader changes to behavior when school is in session could lead to increases in community transmission.

There is also a growing body of evidence that younger children (e.g., those less than 10 years of age) are less susceptible to infection when exposed (*10*), though it is unclear if they are less likely to pass on the virus once infected (*11*, *12*), or if this reduced susceptibility is offset by the increased number of contacts students make when in school (*13*). Even when students are infected, the risk of severe disease and death among teenagers and young children is low (*14*). This means that one of the main reasons for a focus on schools is not the risk to students, but the risk that in-person schooling poses to teachers and family members, as well as its impact on the trajectory of the overall epidemic. Yet, few studies have focused on the risk in-person school poses to household members (*15*).

Different interpretations of the evidence and local politics have led to massive heterogeneity in approaches to schooling across the United States during the 2020-21 school year (*16*), running the gambit from complete cessation of in-person learning to opening completely with no mitigation measures in place. Most schools that have opened have made some efforts to mitigate transmission, but there is much diversity in the approaches adopted.

This hodgepodge of approaches to schooling creates a massive natural experiment from which we can learn about what does, and does not, work for controlling school associated SARS-CoV-2 spread. However, there is no central repository for the measures taken across the over 130,000 schools in the United States, nor is there a central repository for health outcomes in these schools, making studies difficult. Where data are available they are often restricted to traditional public school systems, though 28% of Pre-K through 12th grade students are in private or charter schools, and rarely can it be linked with individual- or household-level outcomes.

The COVID-19 Symptom Survey provides a unique opportunity to collect and analyze data on schooling behaviors and SARS-CoV-2 related outcomes from households throughout the United States. This survey is administered through the Facebook platform in partnership with Carnegie Mellon University and yields approximately 500,000 survey responses in the United States each week (*17*). It includes questions on symptoms related to COVID-19, testing and, since late November 2020, the schooling experience of any children in the household. Analysis weights adjust for non-response and coverage bias (see supplement for details).

We analyzed data collected over two time periods during the 2020-2021 school year (Nov. 24, 2020-Dec. 23, 2020 and Jan. 11 2021-Feb. 10, 2021). Of 2,142,887 total respondents in the 50 US states and Washington DC during this period, 576,051 (26.9%) reported at least one child in Pre-K through high school living in their household (Table S1-S2, Fig. 1A). Forty-nine percent (284,789/576,051) of these respondents reported a child living in the household engaged in either full- (68.8%) or part-time (46.0%) in-person schooling, with substantial variation both within and between states (Fig. 1, Table S3). Overall, in-person schooling increased between the two periods from 48% to 52%, though decreases were observed in some states (e.g., Arizona) (Fig. S1, Table S3).

**Figure 1.**
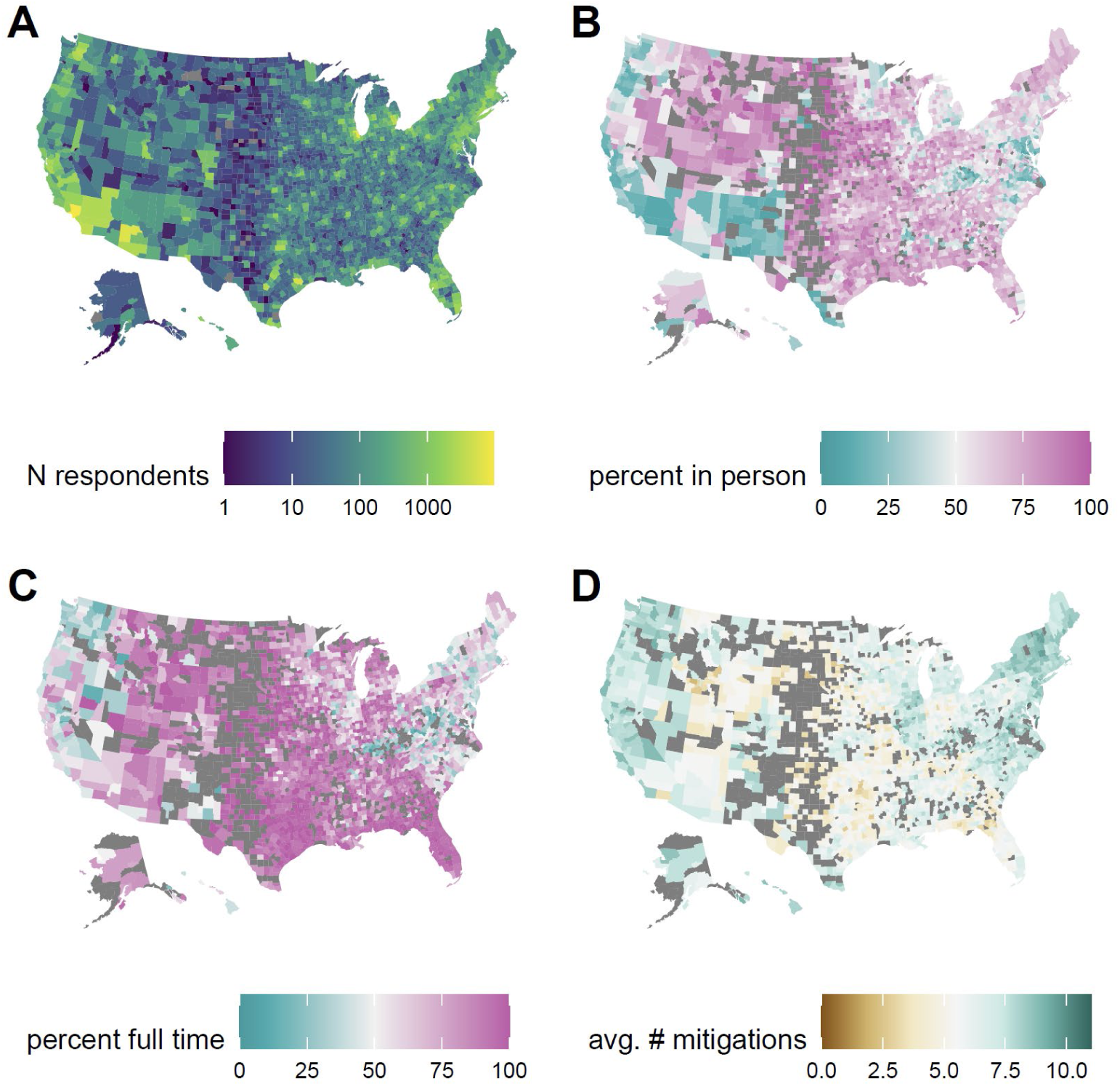
Spatial distribution of survey responses. (**A**) Number of survey respondents reporting a school age student in the household by county. (**B**) Percentage of households with school age children reporting any in-person schooling by county, excluding counties with fewer than 10 responses (excluded counties in dark grey). (**C**) Percentage of households with a child in in-person schooling reporting full-time in-person schooling, excluding counties with fewer than 10 reporting in-person schooling, (**D**) Average number of school-based mitigation measures reported for children with in-person schooling, excluding counties with fewer than 10 reporting in-person schooling.

After adjusting for county-level incidence and other individual- and county-level factors (but not school-based mitigation measures; Tables S1-S2, Fig. S2), living in a household with a child engaged in full-time in-person schooling is associated with a substantial increase in the odds (adjusted odds ratio [aOR] 1.38, 95% CI 1.30-1.47) of reporting COVID-19 like illness (CLI, fever of at least 100 °F, along with cough, shortness of breath, or difficulty breathing), loss of taste or smell (aOR 1.21, 95% CI 1.16-1.27), and report of a positive SARS-CoV-2 test result within the previous 14 days (aOR 1.30, 95% CI 1.24-1.35) (Fig. 2A, Table S4).

**Figure 2.**
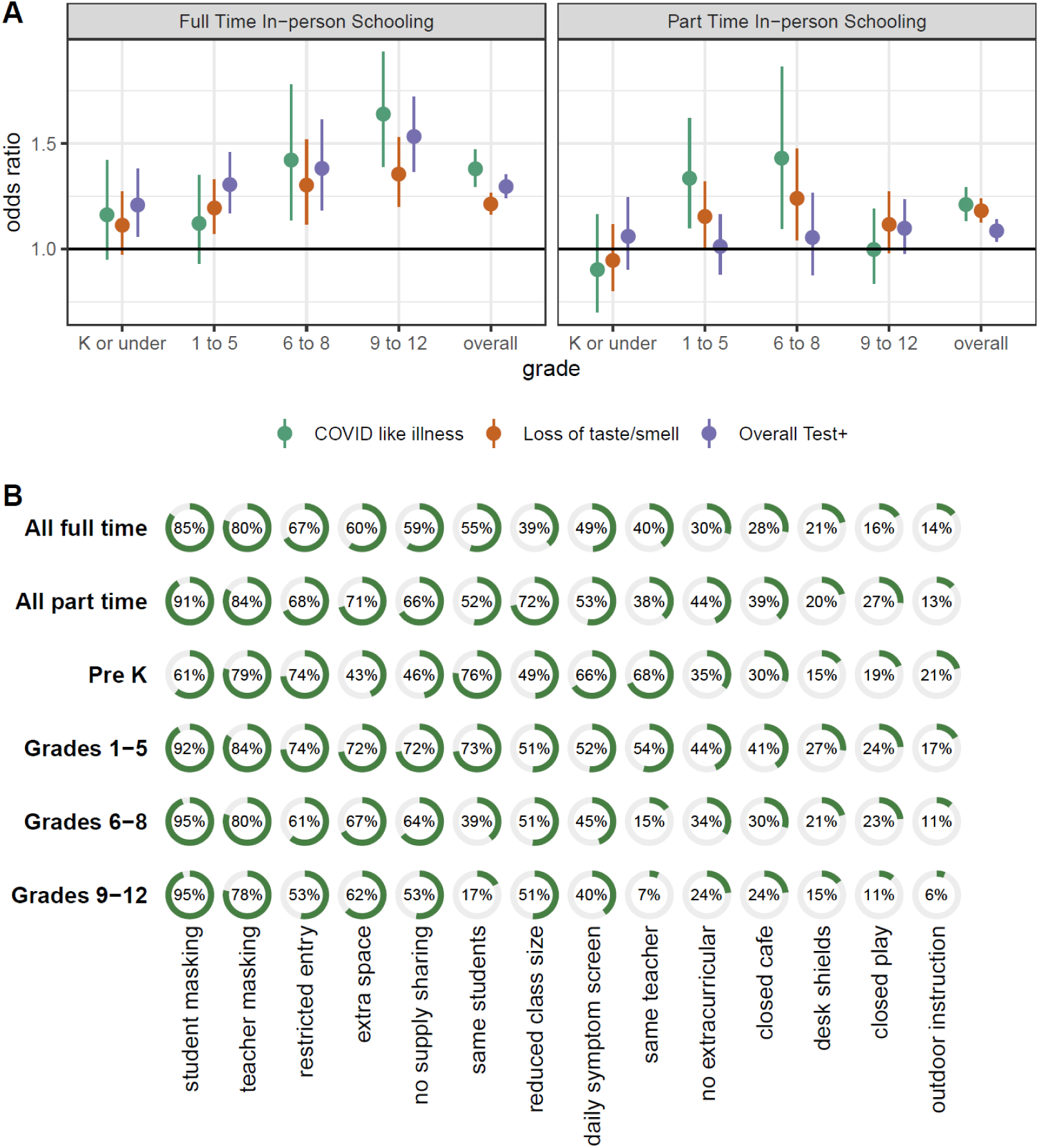
Risk from in-person schooling and distribution of mitigation measures by grade. (**A**) Odds ratio of COVID-19-related outcomes associated with full- and part-time in-person schooling by outcome and grade level, compared to individuals with children in their household not attending in-person schooling and adjusted for individual- and county-level covariates (but not number of mitigation measures) indicating that the strength of the association increases with grade level. (**B**) Distribution of mitigation measures by grade level and full- versus part-time in-person status across all grades.

When stratifying by grade level (restricted to households reporting a child/children in a single grade strata), we find that the strength of the associations increase with grade level, from no association in households with only Pre-K and Kindergarten students (Fig. 2A, Table S4) to a substantial positive association in households with only high school aged students.

The association between COVID-19 outcomes and reporting a child in the household engaged in part-time in-person schooling is attenuated but still statistically significant for CLI (aOR 1.21, 95% CI 1.13-1.29), loss of taste and smell (aOR 1.18, 95% CI 1.13-1.24) and reporting a positive test (aOR 1.09, 95% CI, 1.03-1.14). Among those reporting part-time schooling the association between grade and COVID-19-related outcomes is less clear (Fig. 2A, Table S4).

For those students engaged in any form of in-person learning, the most common mitigation measure reported was student mask mandates (88%), followed by teacher mask mandates (80%), restricted entry (66%) and extra space between desks (63%) (Table S5). The distribution of mitigation measures reported was similar between those reporting full- and part-time in-person schooling, though most measures were slightly more likely to be reported in the part-time setting (Fig. 2B). Overall, respondents reporting a household child engaged in in-person school reported a mean of 6.7 (IQR 4-9) mitigation measures in place at any school attended by a household child. Those reporting only children in part-time schooling reported more mitigation measures (mean 7.3, IQR 5-10) than those reporting only children in full-time schooling (mean 6.4, IQR 4-9). There is substantial geographic heterogeneity in the number of mitigation measures reported (Fig. 1D, Fig. S3, Tables S5-S6), with households in South Dakota reporting the least (mean 4.6, IQR 2-7), and households in Vermont reporting the most (mean 8.9, IQR 8-11) mitigation measures.

We find a clear association with the number of mitigation measures implemented and the risk of COVID-19 outcomes among adult household members responding to the survey after adjustment for individual and county level factors. Each measure implemented is associated with a 9% decrease in the odds of CLI (aOR 0.91, 95% CI 0.89-0.92), a 8% decrease in the odds of loss of taste or smell (aOR 0.92, 95% CI 0.91-0.93) and an 7% decrease in the odds of a recent positive SARS-CoV-2 test (aOR 0.93, 95% CI 0.92-0.94) (Table S7). Regression treating each individual mitigation measure as having an independent effect show that report of daily symptom screening is clearly associated with greater risk reductions than the average measures (Fig. 3, Table S8), with some evidence that teacher mask mandates and cancelling extra-curricular activities are also associated with larger reductions than average. In contrast, closing cafeterias, playgrounds and use of desk shields are associated with lower risk reductions (or even risk increases); however this may reflect saturation effects as these are typically reported along with a high number of other measures. Notably, part-time in-person schooling is not associated with a decrease in the risk of COVID-19-related outcomes compared to full-time in-person schooling once we account for other mitigation measures.

**Figure 3.**
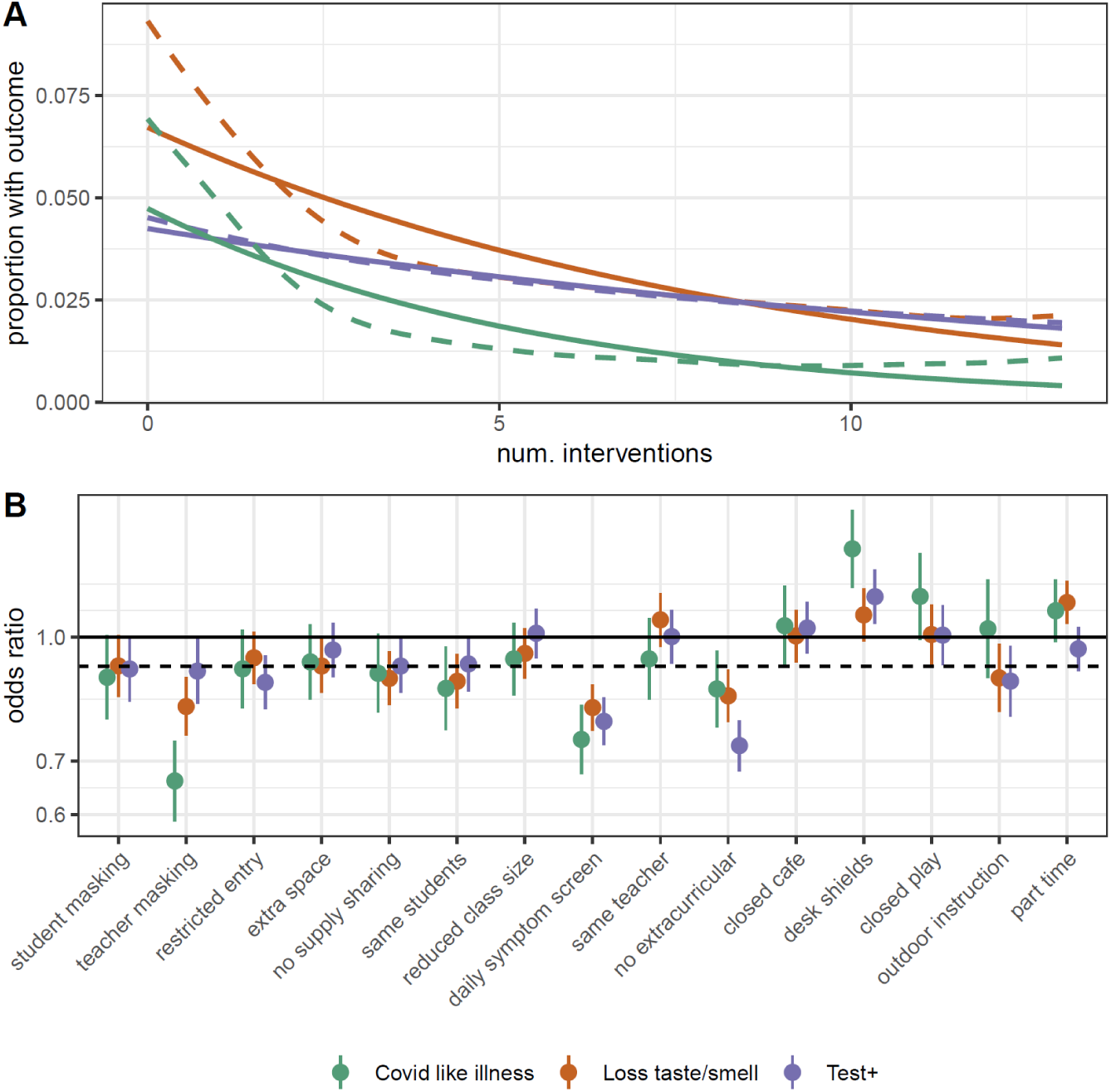
Impact of individual mitigation measures. (**A**) Relationship between number of mitigation measures and percent reporting COVID-19-related outcomes using a log-linear (solid) and spline (dashed) model. **(B)** Odds ratio of COVID-19-related outcomes by mitigation measure in multivariable model including all measures, versus the reduction due to a generic mitigation measure (dotted line).

To explore what, if any, levels of mitigation are associated with elimination of the excess risk posed by in-person schooling, we conducted analyses where we limited the in-person exposure group to schools with 0, 1-3, 4-6, 7-9 and 10 or more mitigation measures in turn (Fig. 4, Fig. S4, Tables S9-S10). We found that when we limited the in-person group to cases where 7 or more mitigation measures were in place the risks associated with in-person schooling had largely disappeared, with complete absence of increased risk with 10 or more mitigation measures. Among those reporting 7 or more mitigation measures, over 80% reported student and teacher mask mandates, restricted entry, extra space between desks and no supply sharing, and over 50% reported student cohorting, reduced class size and daily symptom screening.

**Figure 4.**
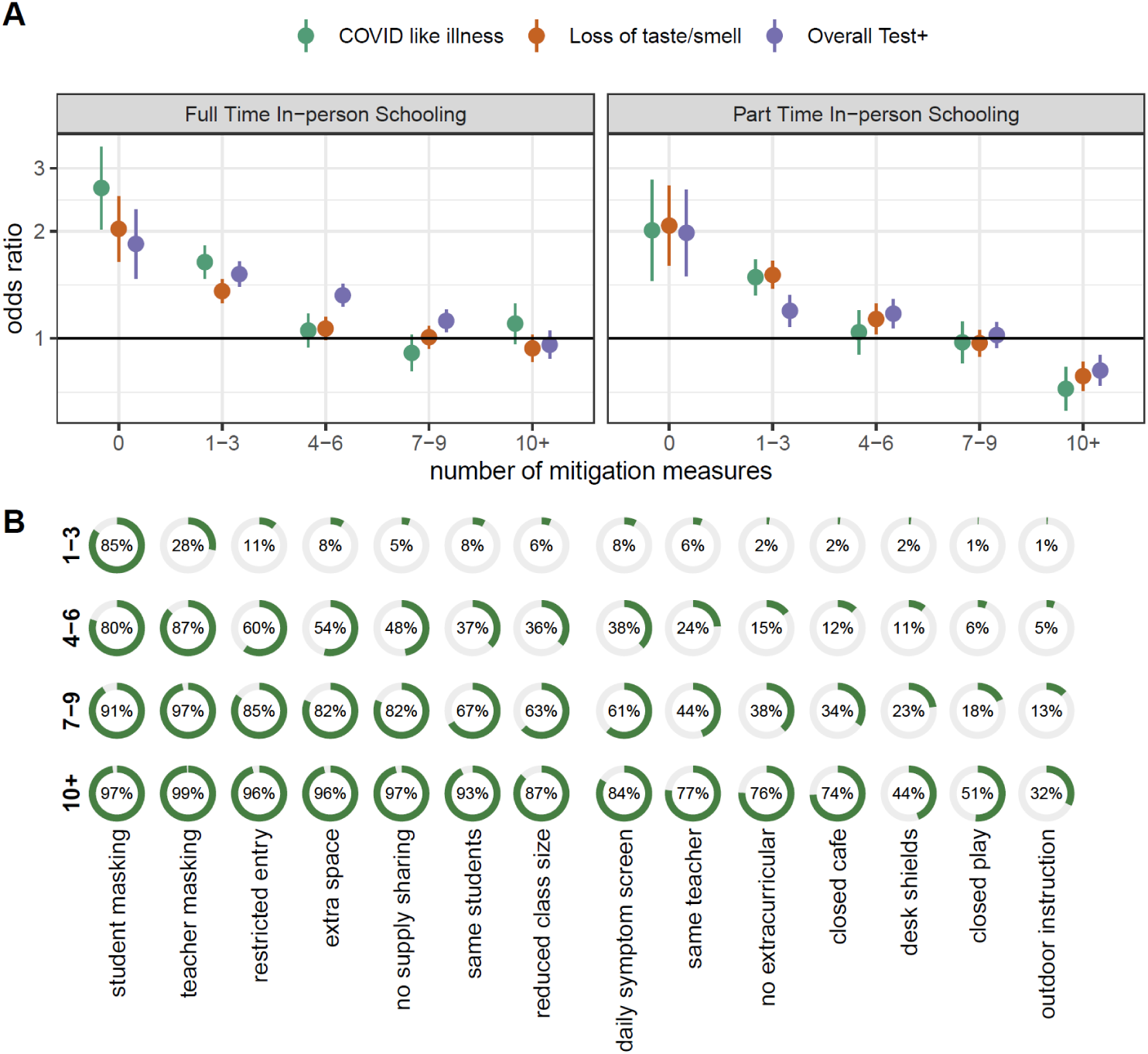
Risk of in-person schooling by strata of number of reported mitigation measures. (**A**) Estimated risk associated with full- and part-time in-person schooling by outcome and number of mitigation measures implemented, adjusted for individual and county-level covariates. (**B**) Distribution of mitigation measures by total number of measures implemented.

The results presented here show a clear association between in-person schooling and the risk of COVID-19-related outcomes in adult household members, as well as evidence that implementation of a moderate number of school-based mitigation measures is adequate to eliminate this risk. However, in-person schooling and mitigation measures are not distributed at random in the population (Fig. 1, Tables S1-S3, S5-S6, S9-S10). For instance, households with a student attending in-person schooling tend to be in counties that are a higher percentage white (Fig. S2), and contain survey respondents who are more likely to have recently eaten out or gone to a bar (Table S2). Despite our best efforts to adjust for local incidence, individual behavior and other potential confounders, it is possible that unmeasured factors are responsible for the observed associations.

To address the possibility that the association with in-person schooling could be the result of differences between urban, suburban and rural counties, local patterns of incidence, or other differences between those more and less likely to send children to school in-person we performed several stratified analyses (Fig. 5). When stratifying by counties classified by size and metro status, incidence and propensity to avoid in-person schooling, we found few systematic or statistically significant deviations from the overall estimate of the relative risk associated with full- and part-time in-person schooling. The notable exception is an apparent increase in the risk associated with in-person schooling in households with the highest propensity to have children attending in-person classes (Fig. 5C).

**Figure 5.**
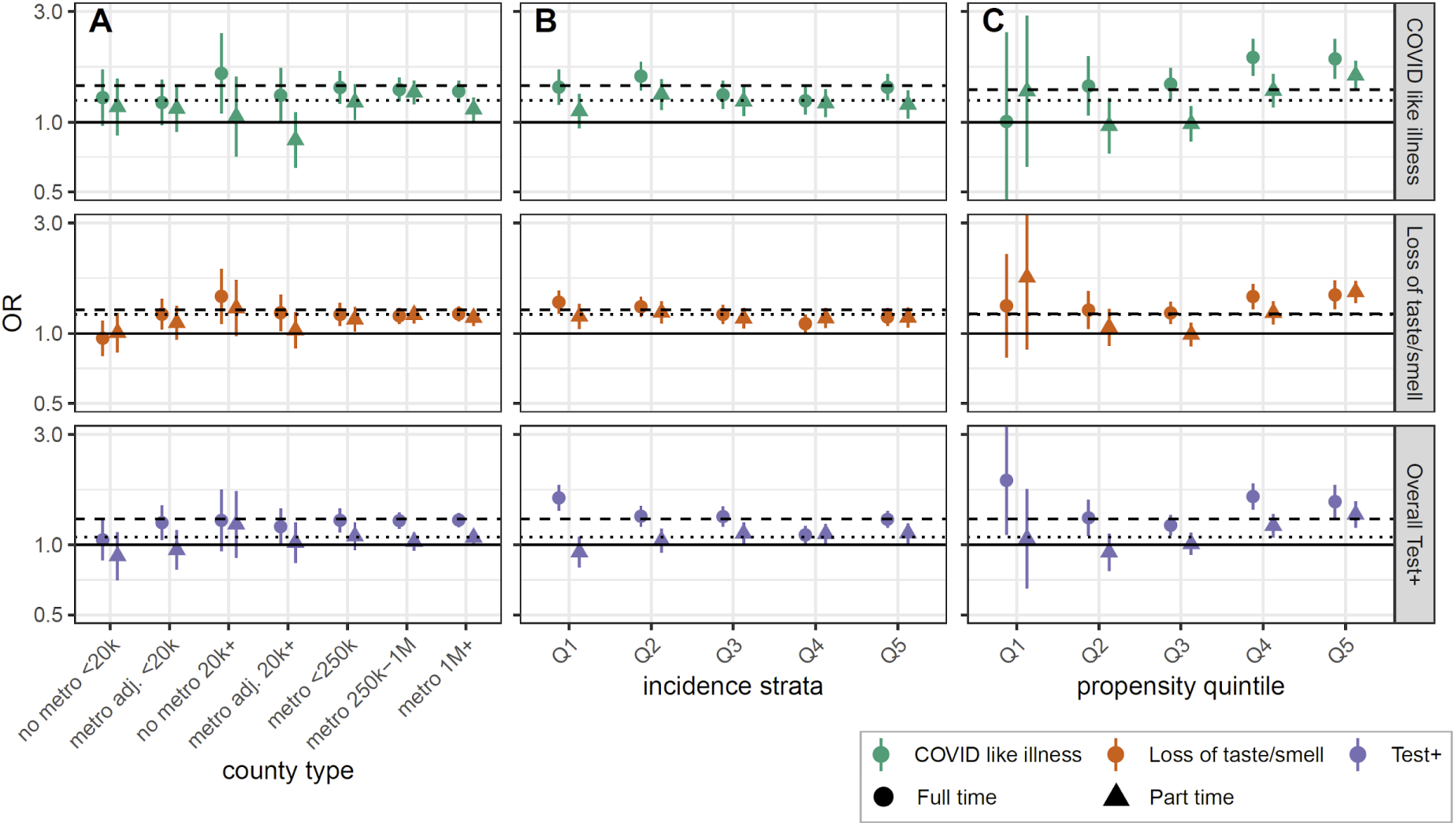
Sub-group analysis of association between in-person schooling and COVID-19-related outcomes. Estimated odds ratio of COVID-19-related outcomes from full-time (circles, dashed lines) and part-time (triangles, dotted line) in-person schooling when data is stratified by (**A**) county population size and relation to metropolitan areas (metropolitan area, non-metropolitan area, adjacent to metropolitan area), (**B**) quintile of incidence (Q1 is lowest, Q5 is highest) and (**C**) propensity to report in-person schooling (Q5 most likely to have in-person schooling, Q1 least likely). Horizontal dashed and dotted lines show overall point estimates for full-time and part-time in-person instruction, respectively.

While we were not able to specifically look at the relationship between in-person schooling, mitigation measures and risk to teachers, we were able to look at the risk associated with reporting paid work outside the home among those reporting teaching pre-K through high school as their current occupation. Teachers working outside the home work were more likely to report COVID-19-related outcomes (e.g., Test positive aOR 1.8, 95% CI 1.5-2.2; Fig. S5, Table S11), but increased risk was commensurate with the risk of working outside the home in other groups, such as those working in healthcare (aOR 1.7, 95% CI 1.5-1.9) and office work (aOR 1.6, 95% CI 1.5-1.7).

The results presented here provide strong evidence that in-person schooling poses a real risk to those living in the households of students, but that risk can be managed through commonly implemented school-based mitigation measures. However, much still remains unknown. We were unable to measure the risk posed by in-person schooling to the students themselves, nor were we able to specifically assess how different policies impact teachers and other school staff. This study also provides limited insight into the mechanisms by which in-person schooling increases risk, and it remains possible that classroom transmission plays a minor role, and other school related activities drive risk.

Furthermore, despite its size, this internet-based survey is cross-sectional, necessarily short and subject to response biases; though its size and robustness of the results allay some of these concerns. We were unable to evaluate compliance with or investment in reported mitigation measures. Though we adjust for several county-level measures of socioeconomic status and these data were not available at the individual level, such factors are known to be associated with COVID-19 risk and attitudes about in-person schooling. Additional, more formal, studies that span schools with multiple policies and approaches are critical to finding definitive answers to these questions. However, our results were robust when we looked with strata of urbanization, background COVID-19 risk, and propensity for in-person schooling (Table 5), and when we examined alternative measures of individual and household COVID-19 occurrence (Fig. S6-S8).

The debate around in-person schooling in the United States has been intense, and has exacerbated differences in approach between independent school systems and individual families nationally. The lack of coordination has provided an opportunity to learn about the risks of in-person schooling, and the degree to which mitigation measures may reduce risk. While online surveys have their unique limitations, the wide reach of the Facebook COVID-19 Symptom Survey has allowed us to gather data from households engaged in heterogeneous schooling activities throughout the country in a way few other study designs could. In analyzing these data, we find further support for the idea that in-person schooling carries with it increased COVID-19 risk to household members; but also suggests common, low cost, mitigation measures can greatly reduce this risk.

## Data Availability

Data are freely available from the CMU Delphi Research Group to researchers at universities and non-profits as detailed at Getting Data Access - Delphi Epidata API (cmu-delphi.github.io). All analytic code with dummy data sets is available at https://github.com/HopkinsIDD/inperson-schooling-covid-survey (note this code will not reproduce paper tables and figures without obtaining underlying data from CMU).

## Acknowledgements

This research is partially based on survey results from Carnegie Mellon University’s Delphi Group.

## Funding

Johns Hopkins University Discovery Award (EB, CL, EAS)

Johns Hopkins University COVID-19 Modeling and Policy Hub Award (EB, CL, EAS) Department of Health and Human Services (JL, MKG)

## Author Contributions

Conceptualization: JL, MKG, CJEM, ASA, EAS

Methodology: JL, MKG, EAS

Investigation: JL, MKG, EB, CL, KHG

Visualization: JL, MKG, KHG

Funding acquisition: JL, EAS

Project administration: JL, EAS

Supervision: JL, MKG, EAS

Writing – original draft: JL, MKG, KHG, EB, CJEM, CL, ASA, EAS

Writing – review & editing: JL, MKG, KHG, EB, CJEM, CL, ASA, EAS

## Competing interests

Authors declare they have no competing interests.

## Supplementary Materials

Methods

Table S1 – S11

Figure S1 – S9

## Supplementary Materials

### Methods

#### The Facebook Symptom Survey

The US COVID-19 Symptom Survey is a cross-sectional survey conducted daily by Carnegie Mellon University, where Facebook provides its application as a platform to recruit participants. Each day, selected users get an invitation to participate in the survey at the top of their Facebook News Feed. The survey instrument asks various questions related to health symptoms, testing, schooling, mental health, and several preventive behaviors in the context of the ongoing pandemic. It was developed by public health and survey experts, and was reviewed and approved by the Institutional Review Boards of both the University of Maryland and Carnegie Mellon University. To provide adequate geographic coverage, stratified random sampling within US states is used. To account both for systematic demographic differences between the sampling frame of Facebook users (i.e. the Facebook Active User Base (FAUB) aged 18+ living in the US) and the United States population, and for bias related to non-response and coverage, Facebook employs a two-stage weighting process ((*18*). In the first stage, inverse propensity score weighting is used to adjust for non-response bias by making the sample more representative of the FAUB. The covariates used in this step are obtained from internal Facebook data, including self-reported age, gender, geographical variables, and other Facebook user characteristics that they found to correlate well with survey responses in the past. In the second stage, post-stratification is used to balance the state-level distribution of age and gender among the Facebook population based on the Current Population Survey 2018 March Supplement March Supplement.

The resulting weights are provided as part of the microdata and can be used to adjust estimates so that the survey population is representative of the US population -- adjusting both for the differences between the US population and US Facebook users, and for the propensity of a Facebook user to take the survey in the first place.

Each time Facebook links a user to the survey instrument, it generates a unique, non-informative random ID number and sends it back to CMU. If the user completes the survey (i.e. provides valid answers to a given basic set of survey questions), CMU sends back the corresponding ID number back to Facebook, and in return receives the weight whose computation was described above. Throughout the entire process, Facebook has no access to individual survey responses to weight the data, and CMU doe s not receive any information linking the survey respondents to their Facebook information (*18*, *19*)

There had been eight waves of the COVID-19 Symptom Survey conducted, with changes to the instrument in each wave including the addition or deletion of questions (*19*, *20*). Our period of study encompasses Waves 5 to 8. Wave 5 took place from November 24 to December 18, 2020. Wave 6 took place from December 19, 2020 to January 11, 2021. From this round we excluded observations between December 24, 2020 and January 10, 2021, corresponding with the typical winter school holidays. Wave 7 took place from January 12 to February 7, 2021. Finally Wave 8 began on February 8, 2021. We include data up to February 10, 2021.

#### Case Data

Case data were obtained from data compiled by the Johns Hopkins Center for Systems Science and Engineering (JHU-CSSE) COVID-19 Dashboard (*21*) . Relevant county-level attack rates were calculated as the average 2-week incidence from 2020-11-15 to 2020-12-27 for period one and 2020-12-27 to 2021-01-31 for period two. Attack rates were calculated by dividing reported cases by estimated 2020 county populations obtained from the US Census bureau using the `tidycensus` package (https://cran.r-project.org/package=tidycensus). Adjustment for attack rates was done based on the log base 2 cases per thousands (i.e., log2([average biweekly attack rate per 1000]+1)).

#### County Level Covariates

In addition to county level attack rates, multiple county-level measures of socioeconomic status were included in adjusted analyses. These variables were derived from the 2014-2018 American Community Survey, obtained using the *tidycensus* package. The county level variables included in these analyses are:

- Total population
- Total population per square mile
- Percent of population that is white
- Percent of population that is Black
- Percent of households with income level under the appropriate poverty threshold
- Percent of households with computer
- Percent of households with broadband internet subscription
- Percent of population without health insurance
- Percent of population employed as essential workers (in agriculture, construction, wholesale, transportation, education or health)
- GINI index of income inequality
- County type (metropolitan, metropolitan-adjacent, or non-metropolitan, by county size)
- County urban-rural classification

#### Outcomes and Covariates

Due to varying relationships between the timeframe of reported behaviours and other exposures and the potential for resulting biases (e.g., a reported behaviour being the result of receiving a positive COVID-19 test, not the other way around), we considered multiple COVID-19-related outcomes. We considered three primary outcomes, as reported and experienced by the survey respondent:

- **COVID-19 like illness (CLI),** defined as a fever of at least 100 °F, along with cough, shortness of breath, or difficulty breathing in the past 24 hours.
- Experience of **loss of taste or smell** within the last 24 hours.
- Report of a **positive COVID-19 test** within the past 14 days.
We also considered the following secondary outcomes, as reported by the survey respondent, to consider household-level risk and motivation for testing:
- Any member in the household experiencing COVID-19 like illness, defined as fever of at least 100 °F, along with cough, shortness of breath, or difficulty breathing, in the past 24 hours
- Direct contact in the last 24 hours by the survey respondent with a household member who recently tested positive for COVID-19. A “direct contact” was defined as a conversation lasting more than 5 minutes with someone closer than 6 feet away or physical contact.
- Report of a positive COVID-19 test result for survey respondent within the past 14 days when testing was indicated (due to either illness or contact with someone who was ill or tested positive for COVID-19)
- Report of a positive COVID-19 test result for survey respondent within the past 14 days when testing was not indicated (testing was required by an employer, school, or while receiving other medical care *and* the individual was not ill and had not been in contact with someone who was ill or tested positive for COVID-19).

We considered the adjusted associations between each of these outcomes with reported in-person schooling practices, including reported mitigation measures in place. Responses for in-person schooling practices were provided at the household level (e.g., “Is any child in the household going to in-person classes?”). Respondents were asked whether each of the following mitigation measures was in place where their child(ren) attended in-person classes:

- Mandatory mask-wearing for students
- Mandatory mask-wearing for teachers
- Student is with the same teacher all day
- Student is with the same students all day
- Some or all outdoor instruction
- Restricted entry into school (e.g. no parents or caregivers)
- Reduced class sizes
- Closed cafeteria
- Closed playground
- Use of separators or “desk shields” in classrooms
- Extra space between desks in classrooms
- No school-based extracurricular activities (e.g. sports, clubs, after school care)
- No sharing of books and/or supplies (e.g. each student has their own set at their desk)
- Daily symptom screening for those going onto campus

Various other individual- and household-level variables were collected in the survey and used in adjusted analyses. Several variables were re-coded to account for out of range and nonsensical responses and to aid in interpretability. The survey variables used in these analyses are:

- **State** and **zip code** of residence. **County** of residence is derived from reported zip code; both state and county are used to match to county-level covariates. That is, if the reported county was not within the reported state, county-level covariates were marked as missing (n=4,362 among households with >=1 reported school-aged child).
- **Age** of survey respondent, categorized as 18-24, 25-34, 35-44, 45-54, 55-64, or 65+ years of age
- **Gender,** re-coded to whether the survey respondent identified as male or not, due to small number of non-binary or other respondents.
- **Employment status** of the survey respondent in the last 4 weeks
- Primary **occupation** of the survey respondent in the last 4 weeks, where occupations reported by less than 10,000 people (50,000 in the teacher sub-analysis) were collapsed into the “Other” category.
- Indicator for whether the respondent’s employment required paid **work outside of the home** in the last four weeks.
- **Number of kids, adults, and people 65 years or older in the household.** Negative, non-integer and extreme (≥100) values were recorded as missing. The number of kids re-coded into eight categories (0,1,2,…,6,7+), and the number of adults, people over 65 years, and total household members were re-coded into 11 categories (0,1,2,…,10+). Variables for number of known sick contacts, number of people encountered in recent social gatherings, and the number encountered during shopping were not considered due to high levels of missingness.

- **Grade indicators** for whether there is one or more child in the household in pre-kindergarten or kindergarten; in grades 1 - 5 (elementary school, classically); in grades 6 to 8 (middle school); or in grades 9 - 12 (high school). Only households with at least one school-aged child are included in analyses of household risk.
- **Masking level**, indicating self-reported frequency of mask or facial covering use in public in the last 5 days. Responses were categorized as never, rarely, sometimes, mostly, or always wearing a mask in public, or no reported time spent in public in the past 5 days.
- Indicator for whether the survey respondent reported any **out-of-state travel** in the past 5 days
- Indicators for whether survey respondent reported various **activities** in the past 24 hours, including going to work or school outside place of residence; going to a market, grocery store, or pharmacy; going to a bar, restaurant, or cafe; going to an event with more than 10 people; spending time with someone not currently staying with the survey respondent; and using public transit.
- **Covid testing indicator** for whether the survey respondent received a COVID-19 test in the past 14 days. Missing values were treated as a negative response (i.e., no test received). Only the subset of individuals who received a test in the past 14 days were used in analyses of positive COVID-19 test outcomes among survey respondents.

We also assessed the reported risk of COVID-19-related outcomes associated with education professions among all survey respondents in the two time periods (that is, not restricted to households with school-aged children). In this analysis, individuals who reported their primary occupation as “Education” were divided into two separate categories: “K-12 educator”, for respondents who reported being a preschool, kindergarten, elementary, middle, or secondary school teacher; and “Other educator”, for those who reported being a postsecondary teacher, a teacher assistant, other teacher or instructor including special education, or librarian.

#### Analysis

All analyses were conducted using quasibinomial regression accounting for survey weights described above using the `srvyr` package (https://cran.r-project.org/package=srvyr) in the R statistical language.

Analyses of the risk of in-person schooling were restricted to households with at least one school-aged child. Analyses of the effect of individual in-school mitigation measures and the number of mitigation measures were restricted to households with at least one school-aged child engaged in any in-person schooling. Analyses of the odds of a reported positive test for the survey respondent were further restricted to include only households in which the respondent had been tested for any reason in the last 14 days. Estimates of the risk of in-person schooling and the impact of individual mitigation measures were adjusted by respondent age, gender, occupation, masking behaviors, out-of-state travel, and whether they reported a visit to a bar/restaurant/cafe, to an event with more than 10 people, or whether they used public transit; by household size and number of children; and county 2-week average attack rate, population, percent white population, percent households in poverty, GINI index of income inequality, and metropolitan type.

Propensity scores were used to explore possible confounding in the relationship between in-person schooling and risk of COVID-19-related outcomes. Random forests with 500 classification trees were used to generate propensity scores for a household reporting any child engaged in in-person schooling, using the `ranger` package (*22*) in the R statistical language. Variables included for classification were: respondent age, gender, occupation, masking behaviors, out-of-state travel, reported activities outside of house; household size, number of children, and indicators for child grade level; and county 2-week average attack rate, population, population per square mile, percent white and Black population, percent without health insurance, percent essential workers, percent households in poverty, percent households with computer, percent households with broadband internet subscription, GINI index of income inequality, urban-rural classification and metropolitan type. Variable importance was calculated using the GINI index for classification, or the mean decrease in node impurity (Fig. S9).

Analyses of the risk of COVID-19-related outcome among educational professionals were conducted among all respondents with non-missing occupation. An interaction term between occupation type, including K-12 educator, and an indicator for any paid work outside the home in the last four weeks was used to evaluate the baseline risk associated with each occupation, with extra-household work, and with extra-household work within each occupation. These models were adjusted with the same variable set included in models of risk of in-person schooling, excluding the number of children in the household due to missingness.

**Table S1.**
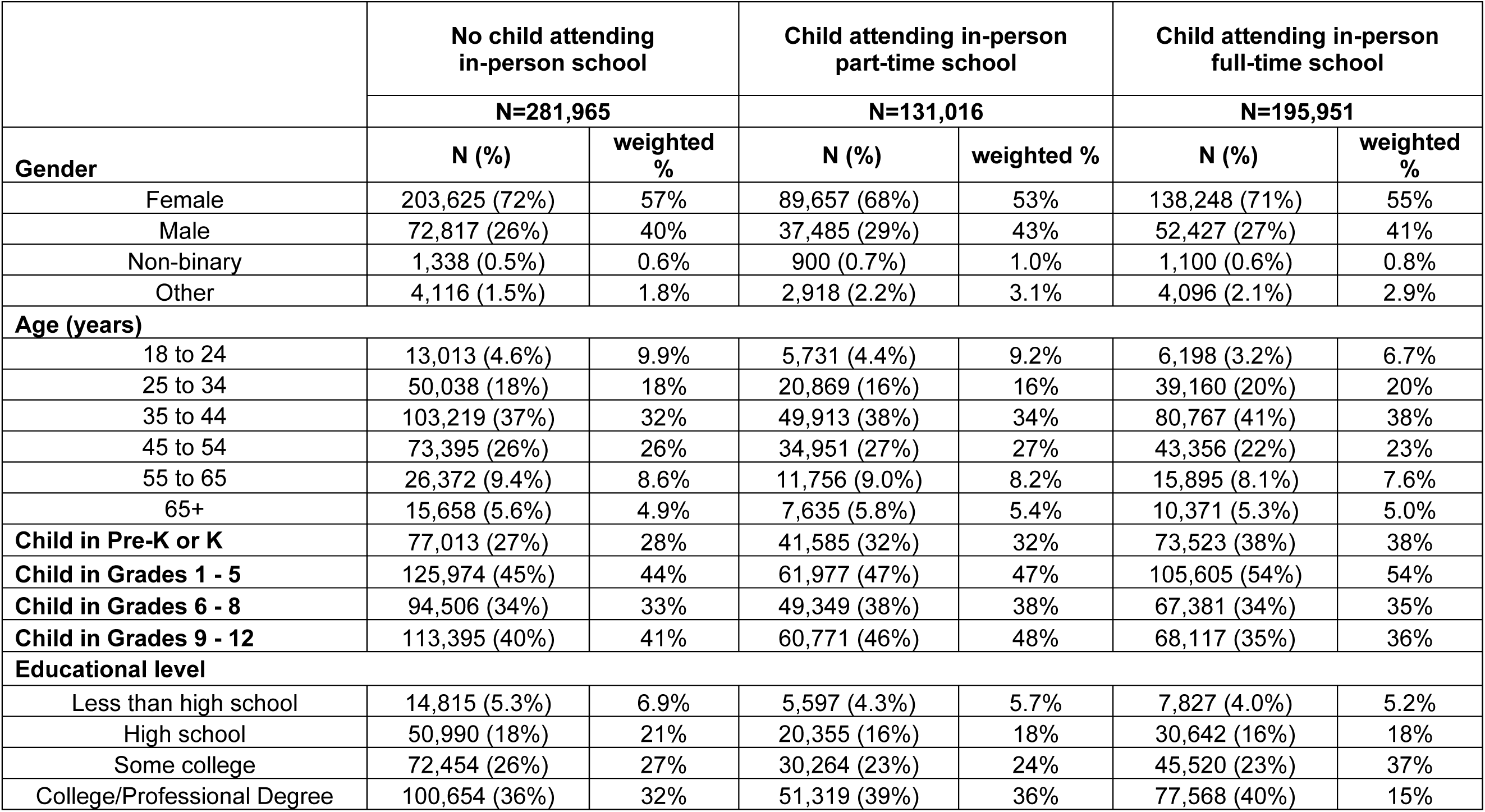
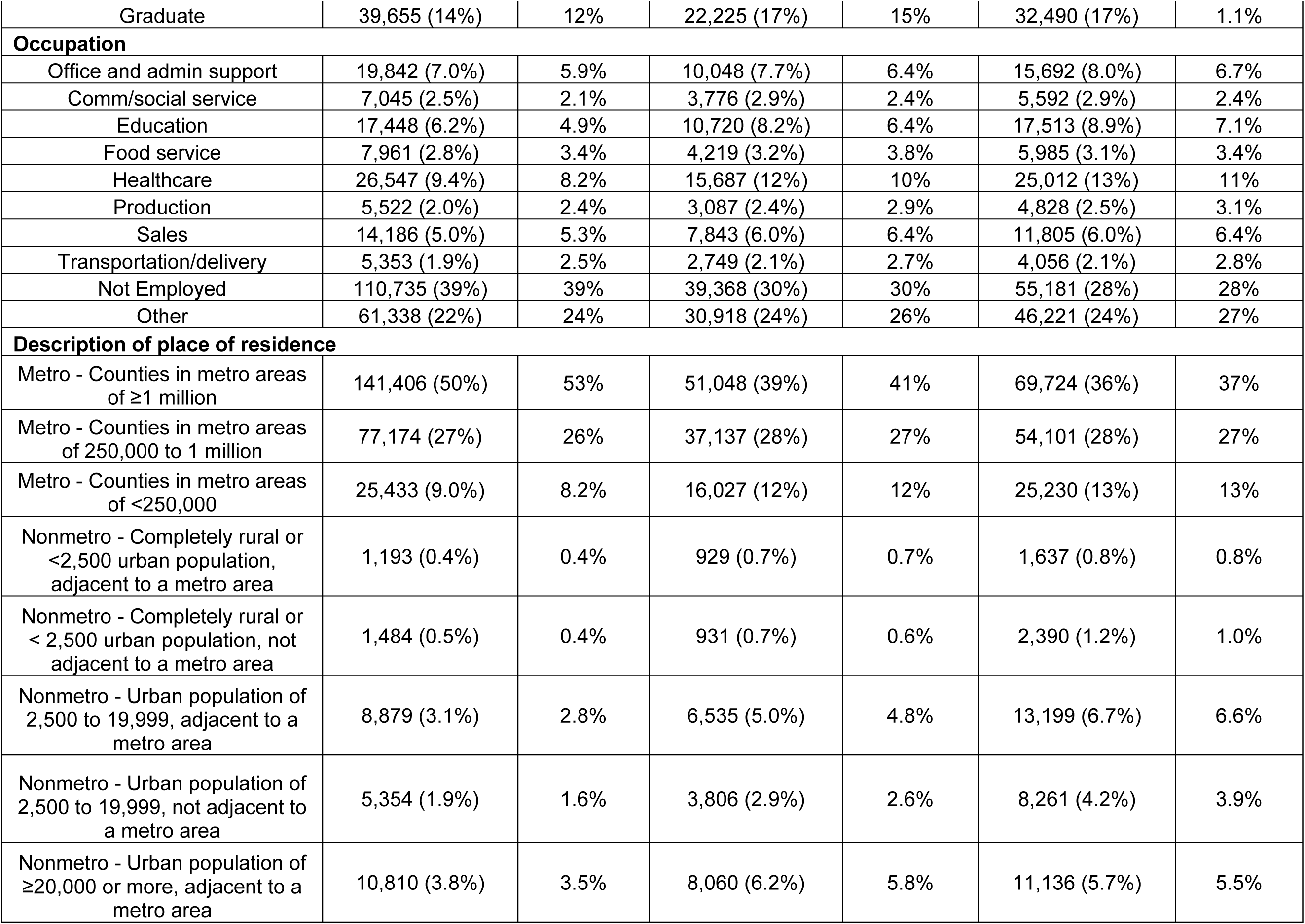
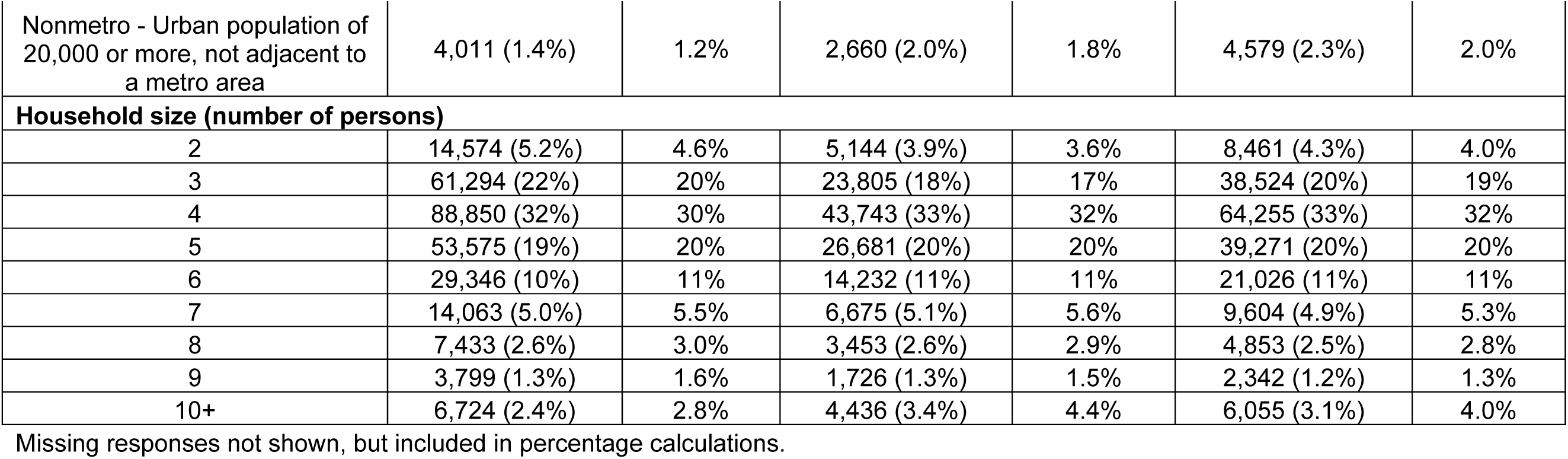
Selected sociodemographics among participants with ≥1 school-aged child in the household comparing those reporting no in-person schooling to those reporting any part-time and full-time in-person schooling; observed and survey-weighted percentages reported.

|  | No child attending in-person school |  | Child attending in-person part-time school |  | Child attending in-person full-time school |  |
| --- | --- | --- | --- | --- | --- | --- |
|  | N=281,965 |  | N=131,016 |  | N=195,951 |  |
| Gender | N (%) | weighted % | N (%) | weighted % | N (%) | weighted % |
| Female | 203,625 (72%) | 57% | 89,657 (68%) | 53% | 138,248 (71%) | 55% |
| Male | 72,817 (26%) | 40% | 37,485 (29%) | 43% | 52,427 (27%) | 41% |
| Non-binary | 1,338 (0.5%) | 0.6% | 900 (0.7%) | 1.0% | 1,100 (0.6%) | 0.8% |
| Other | 4,116 (1.5%) | 1.8% | 2,918 (2.2%) | 3.1% | 4,096 (2.1%) | 2.9% |
| **Age (years)** |  |  |  |  |  |  |
| 18 to 24 | 13,013 (4.6%) | 9.9% | 5,731 (4.4%) | 9.2% | 6,198 (3.2%) | 6.7% |
| 25 to 34 | 50,038 (18%) | 18% | 20,869 (16%) | 16% | 39,160 (20%) | 20% |
| 35 to 44 | 103,219 (37%) | 32% | 49,913 (38%) | 34% | 80,767 (41%) | 38% |
| 45 to 54 | 73,395 (26%) | 26% | 34,951 (27%) | 27% | 43,356 (22%) | 23% |
| 55 to 65 | 26,372 (9.4%) | 8.6% | 11,756 (9.0%) | 8.2% | 15,895 (8.1%) | 7.6% |
| 65+ | 15,658 (5.6%) | 4.9% | 7,635 (5.8%) | 5.4% | 10,371 (5.3%) | 5.0% |
| **Child in Pre-K or K** | 77,013 (27%) | 28% | 41,585 (32%) | 32% | 73,523 (38%) | 38% |
| **Child in Grades 1 - 5** | 125,974 (45%) | 44% | 61,977 (47%) | 47% | 105,605 (54%) | 54% |
| **Child in Grades 6 - 8** | 94,506 (34%) | 33% | 49,349 (38%) | 38% | 67,381 (34%) | 35% |
| **Child in Grades 9 - 12** | 113,395 (40%) | 41% | 60,771 (46%) | 48% | 68,117 (35%) | 36% |
| **Educational level** |  |  |  |  |  |  |
| Less than high school | 14,815 (5.3%) | 6.9% | 5,597 (4.3%) | 5.7% | 7,827 (4.0%) | 5.2% |
| High school | 50,990 (18%) | 21% | 20,355 (16%) | 18% | 30,642 (16%) | 18% |
| Some college | 72,454 (26%) | 27% | 30,264 (23%) | 24% | 45,520 (23%) | 37% |
| College/Professional Degree | 100,654 (36%) | 32% | 51,319 (39%) | 36% | 77,568 (40%) | 15% |
| Graduate | 39,655 (14%) | 12% | 22,225 (17%) | 15% | 32,490 (17%) | 1.1% |
| **Occupation** |  |  |  |  |  |  |
| Office and admin support | 19,842 (7.0%) | 5.9% | 10,048 (7.7%) | 6.4% | 15,692 (8.0%) | 6.7% |
| Comm/social service | 7,045 (2.5%) | 2.1% | 3,776 (2.9%) | 2.4% | 5,592 (2.9%) | 2.4% |
| Education | 17,448 (6.2%) | 4.9% | 10,720 (8.2%) | 6.4% | 17,513 (8.9%) | 7.1% |
| Food service | 7,961 (2.8%) | 3.4% | 4,219 (3.2%) | 3.8% | 5,985 (3.1%) | 3.4% |
| Healthcare | 26,547 (9.4%) | 8.2% | 15,687 (12%) | 10% | 25,012 (13%) | 11% |
| Production | 5,522 (2.0%) | 2.4% | 3,087 (2.4%) | 2.9% | 4,828 (2.5%) | 3.1% |
| Sales | 14,186 (5.0%) | 5.3% | 7,843 (6.0%) | 6.4% | 11,805 (6.0%) | 6.4% |
| Transportation/delivery | 5,353 (1.9%) | 2.5% | 2,749 (2.1%) | 2.7% | 4,056 (2.1%) | 2.8% |
| Not Employed | 110,735 (39%) | 39% | 39,368 (30%) | 30% | 55,181 (28%) | 28% |
| Other | 61,338 (22%) | 24% | 30,918 (24%) | 26% | 46,221 (24%) | 27% |
| **Description of place of residence** |  |  |  |  |  |  |
| Metro - Counties in metro areas of ≥1 million | 141,406 (50%) | 53% | 51,048 (39%) | 41% | 69,724 (36%) | 37% |
| Metro - Counties in metro areas of 250,000 to 1 million | 77,174 (27%) | 26% | 37,137 (28%) | 27% | 54,101 (28%) | 27% |
| Metro - Counties in metro areas of <250,000 | 25,433 (9.0%) | 8.2% | 16,027 (12%) | 12% | 25,230 (13%) | 13% |
| Nonmetro - Completely rural or <2,500 urban population, adjacent to a metro area | 1,193 (0.4%) | 0.4% | 929 (0.7%) | 0.7% | 1,637 (0.8%) | 0.8% |
| Nonmetro - Completely rural or < 2,500 urban population, not adjacent to a metro area | 1,484 (0.5%) | 0.4% | 931 (0.7%) | 0.6% | 2,390 (1.2%) | 1.0% |
| Nonmetro - Urban population of 2,500 to 19,999, adjacent to a metro area | 8,879 (3.1%) | 2.8% | 6,535 (5.0%) | 4.8% | 13,199 (6.7%) | 6.6% |
| Nonmetro - Urban population of 2,500 to 19,999, not adjacent to a metro area | 5,354 (1.9%) | 1.6% | 3,806 (2.9%) | 2.6% | 8,261 (4.2%) | 3.9% |
| Nonmetro - Urban population of ≥20,000 or more, adjacent to a metro area | 10,810 (3.8%) | 3.5% | 8,060 (6.2%) | 5.8% | 11,136 (5.7%) | 5.5% |
| Nonmetro - Urban population of 20,000 or more, not adjacent to a metro area | 4,011 (1.4%) | 1.2% | 2,660 (2.0%) | 1.8% | 4,579 (2.3%) | 2.0% |
| **Household size (number of persons)** |  |  |  |  |  |  |
| 2 | 14,574 (5.2%) | 4.6% | 5,144 (3.9%) | 3.6% | 8,461 (4.3%) | 4.0% |
| 3 | 61,294 (22%) | 20% | 23,805 (18%) | 17% | 38,524 (20%) | 19% |
| 4 | 88,850 (32%) | 30% | 43,743 (33%) | 32% | 64,255 (33%) | 32% |
| 5 | 53,575 (19%) | 20% | 26,681 (20%) | 20% | 39,271 (20%) | 20% |
| 6 | 29,346 (10%) | 11% | 14,232 (11%) | 11% | 21,026 (11%) | 11% |
| 7 | 14,063 (5.0%) | 5.5% | 6,675 (5.1%) | 5.6% | 9,604 (4.9%) | 5.3% |
| 8 | 7,433 (2.6%) | 3.0% | 3,453 (2.6%) | 2.9% | 4,853 (2.5%) | 2.8% |
| 9 | 3,799 (1.3%) | 1.6% | 1,726 (1.3%) | 1.5% | 2,342 (1.2%) | 1.3% |
| 10+ | 6,724 (2.4%) | 2.8% | 4,436 (3.4%) | 4.4% | 6,055 (3.1%) | 4.0% |

Missing responses not shown, but included in percentage calculations.

**Table S2.**
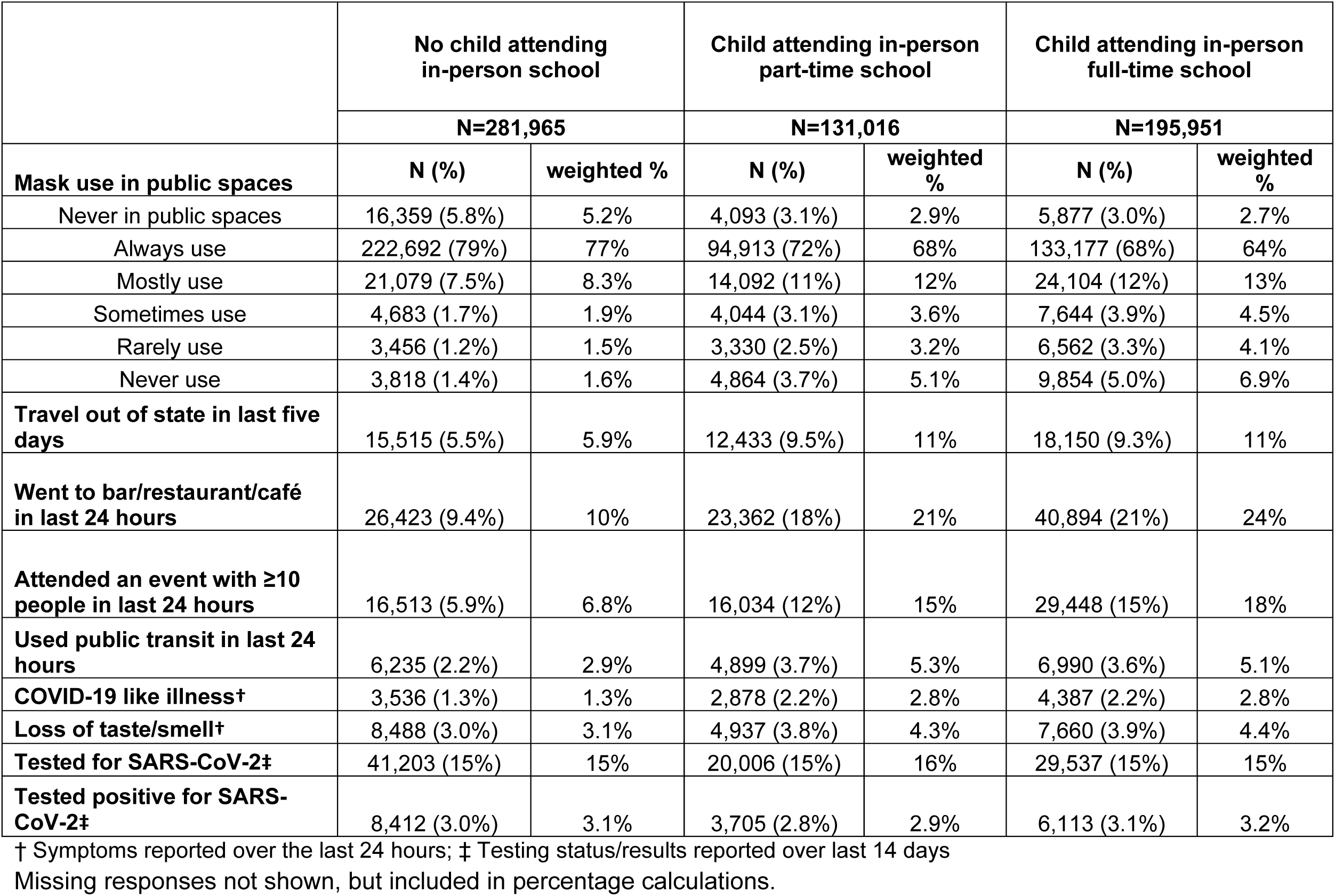
Selected behaviors relevant to COVID-19 acquisition/transmission and COVID-19-related outcomes among participants with ≥1 school-aged child in the household comparing those reporting no in-person schooling to those reporting any part-time and full-time in-person schooling; observed and survey-weighted percentages reported.

**Table S3.**
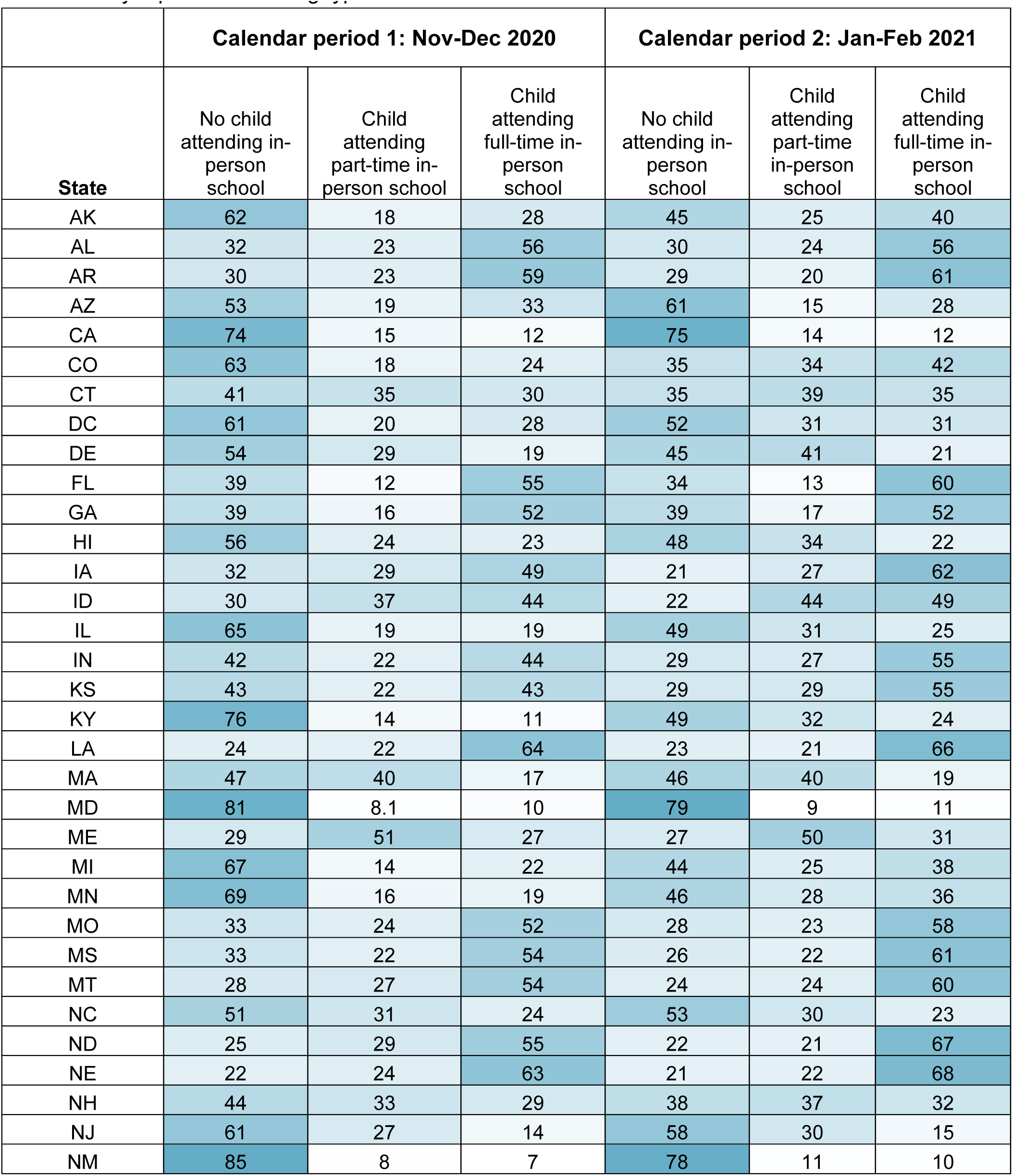
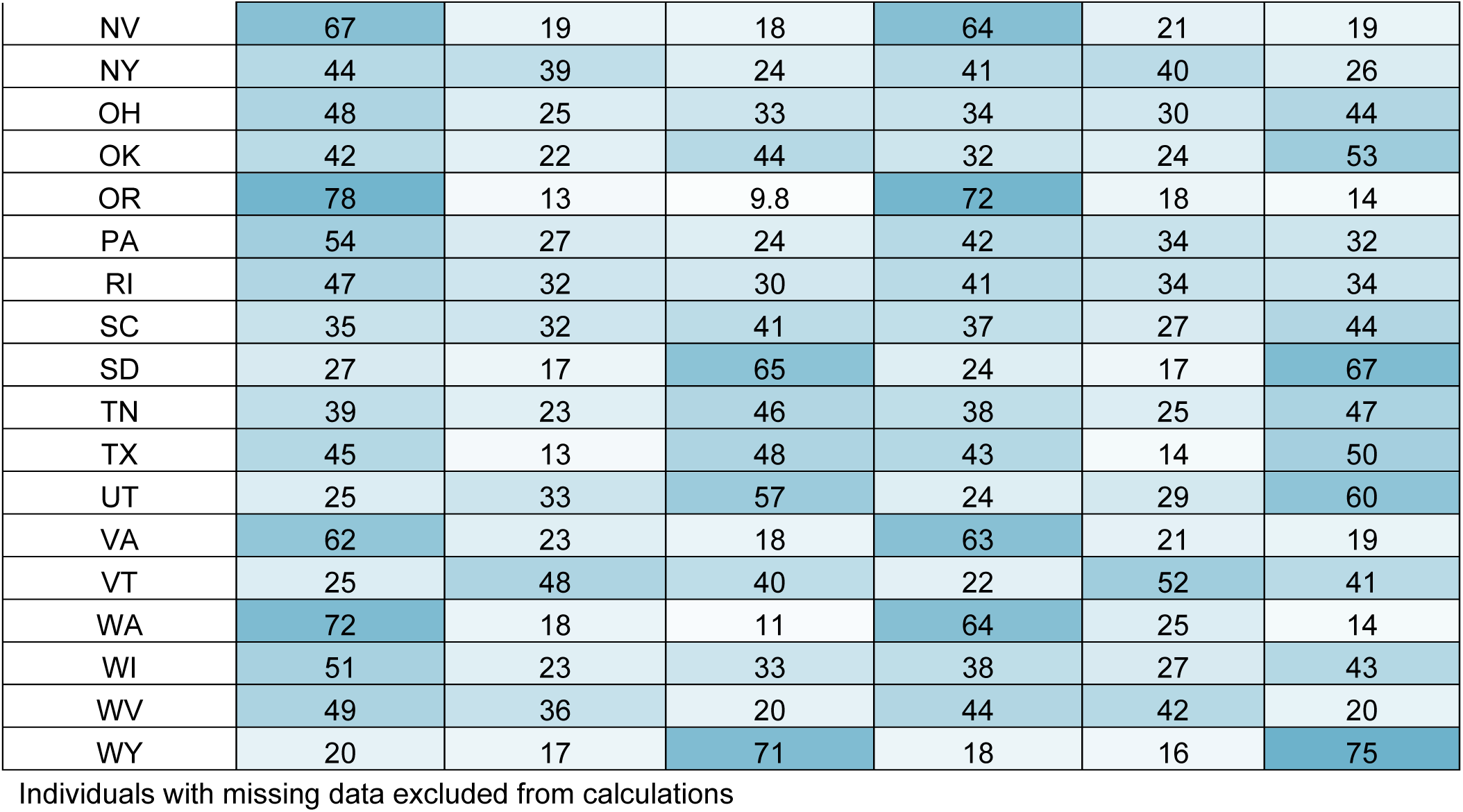
Survey-weighted percentage of participants with ≥1 school-aged child in the household by reported schooling type and state.

**Table S4.**
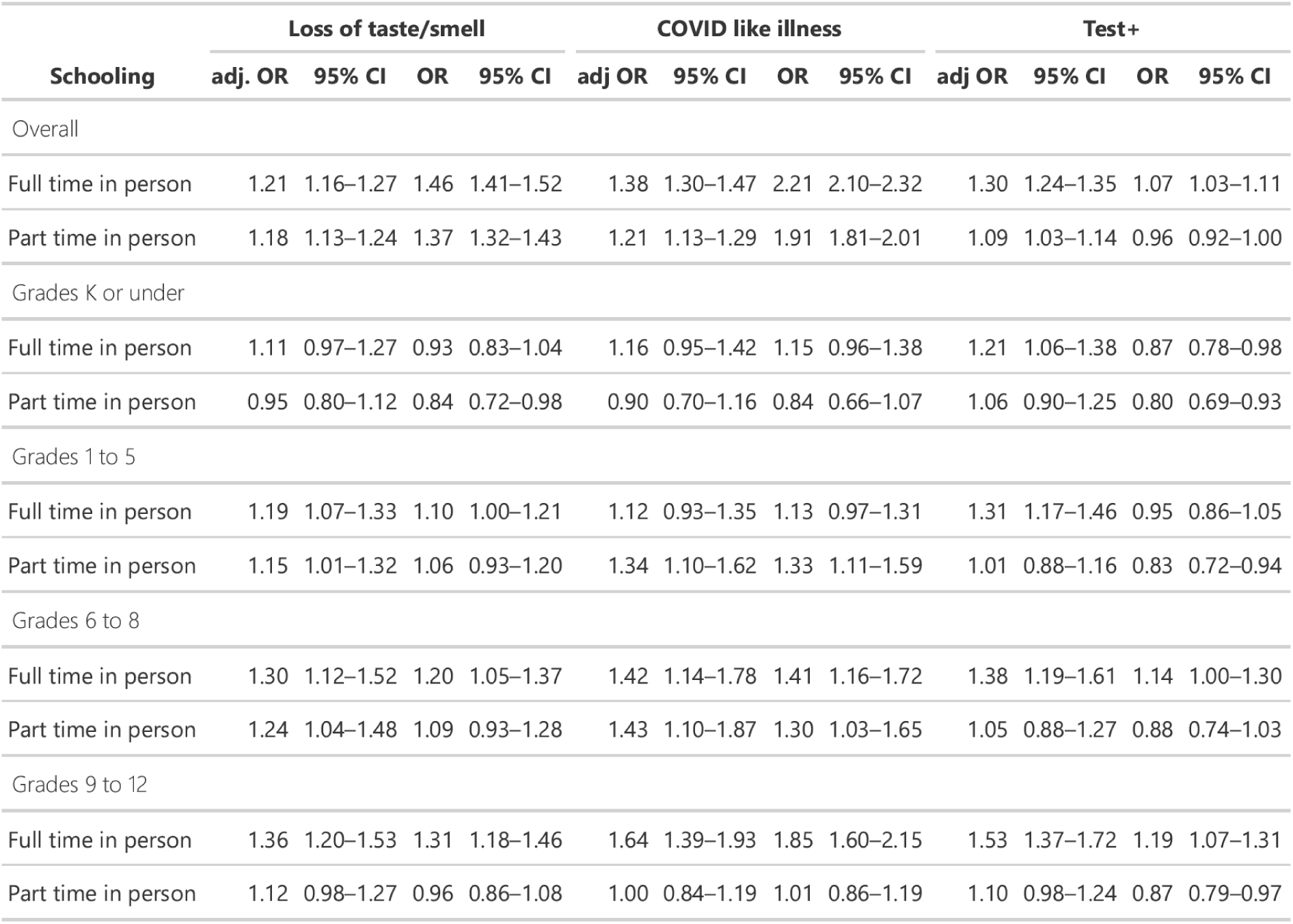
Adjusted and unadjusted odds ratio (OR) of COVID-19 among those engaged in full- and part-time in-person schooling versus those engaged in virtual or homeschooling outcomes. All analyses adjust for survey weights. See methods for adjustment factors.

**Table S5.**
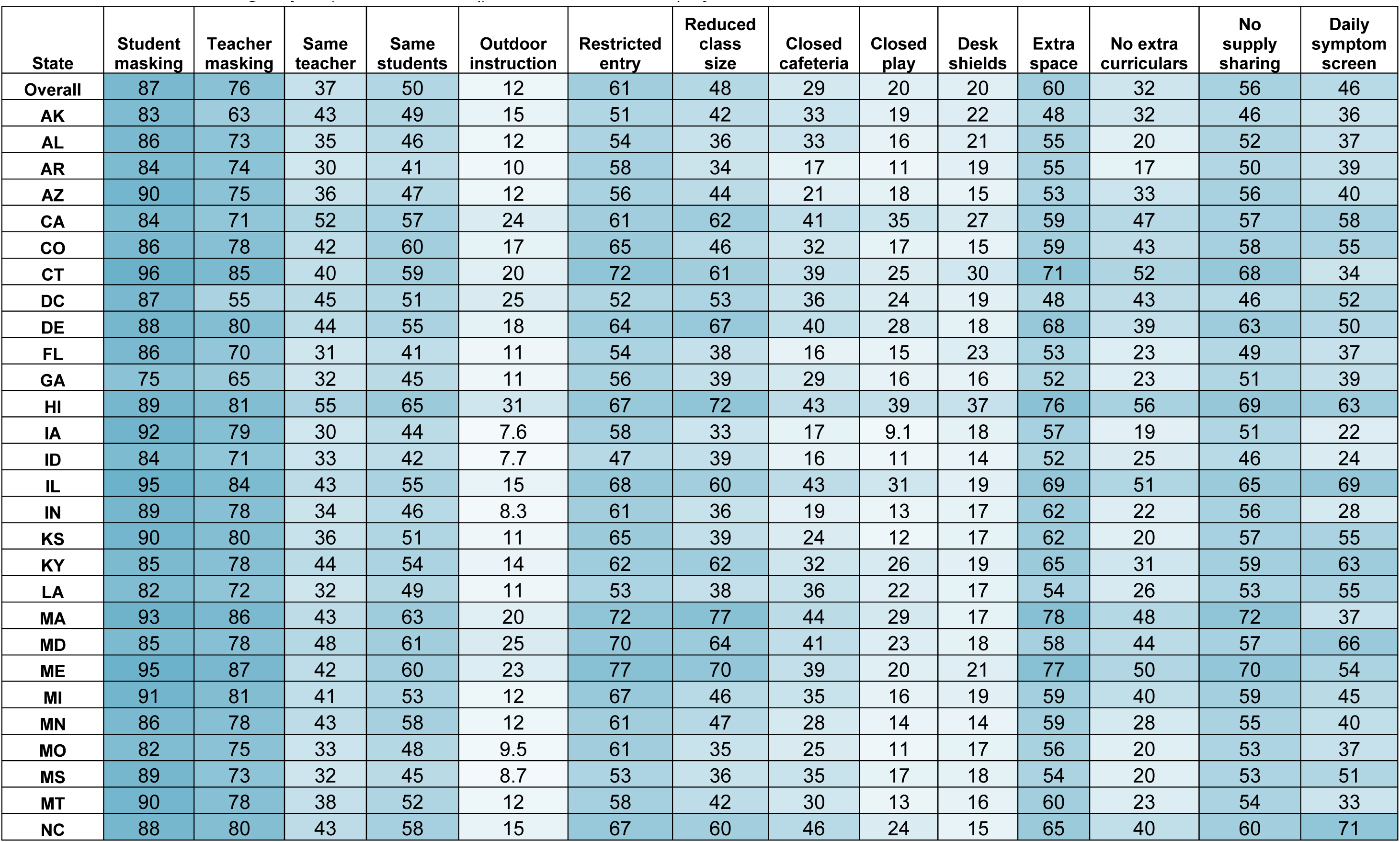
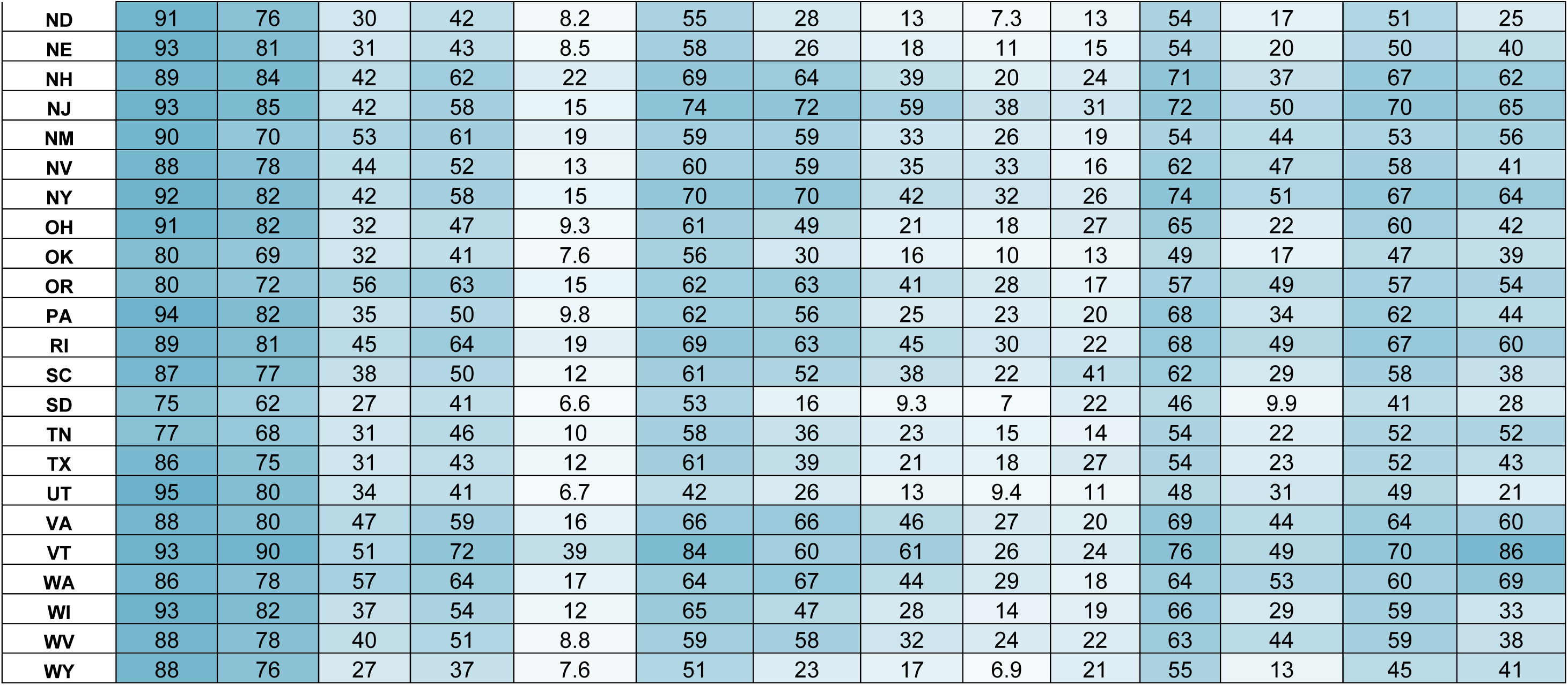
Survey-weighted percentages of participants reporting school mitigation measures among those with ≥1 school-aged child in the household attending any in-person school (part-time or full-time) by state.

**Table S6.**
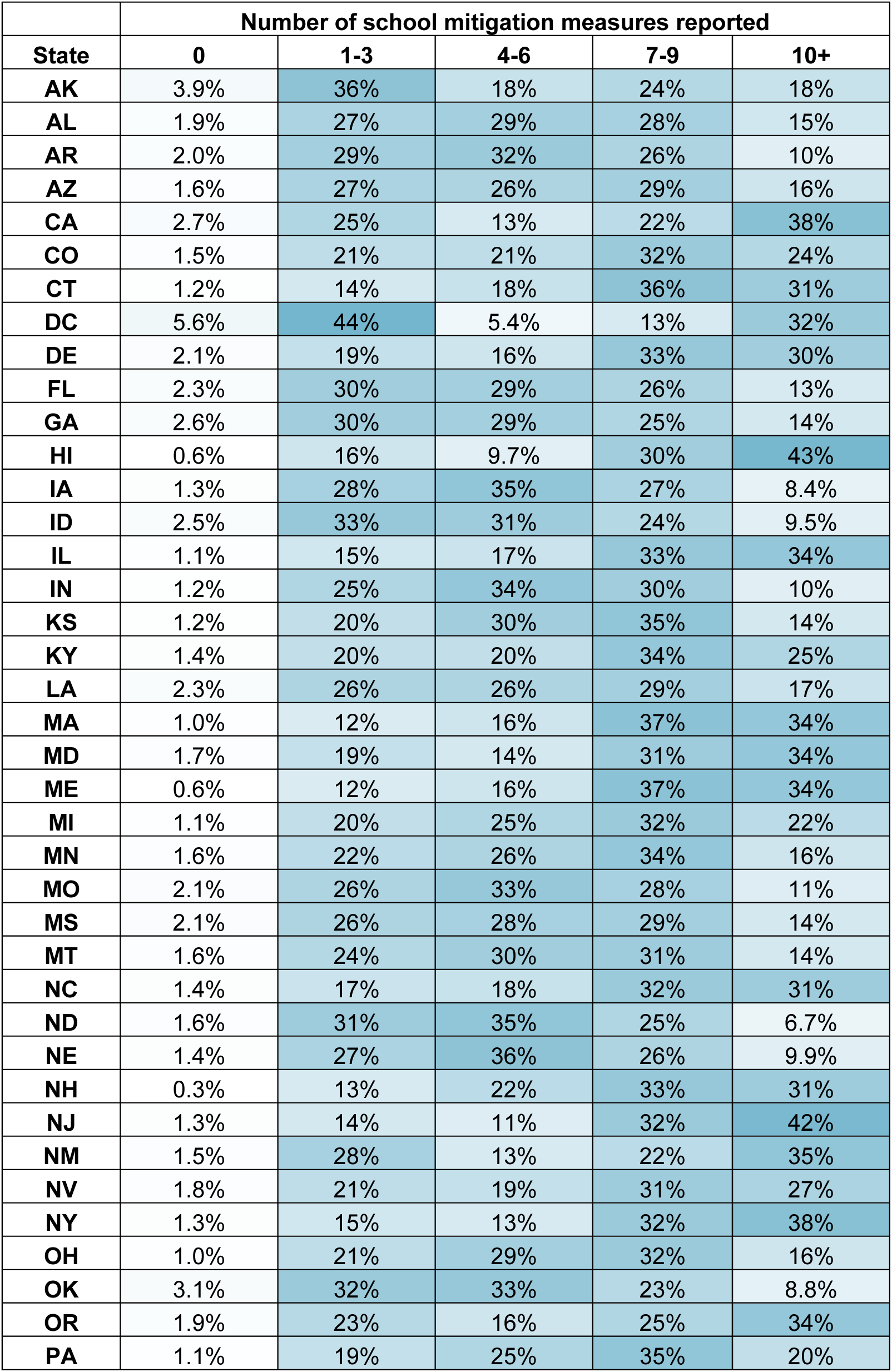
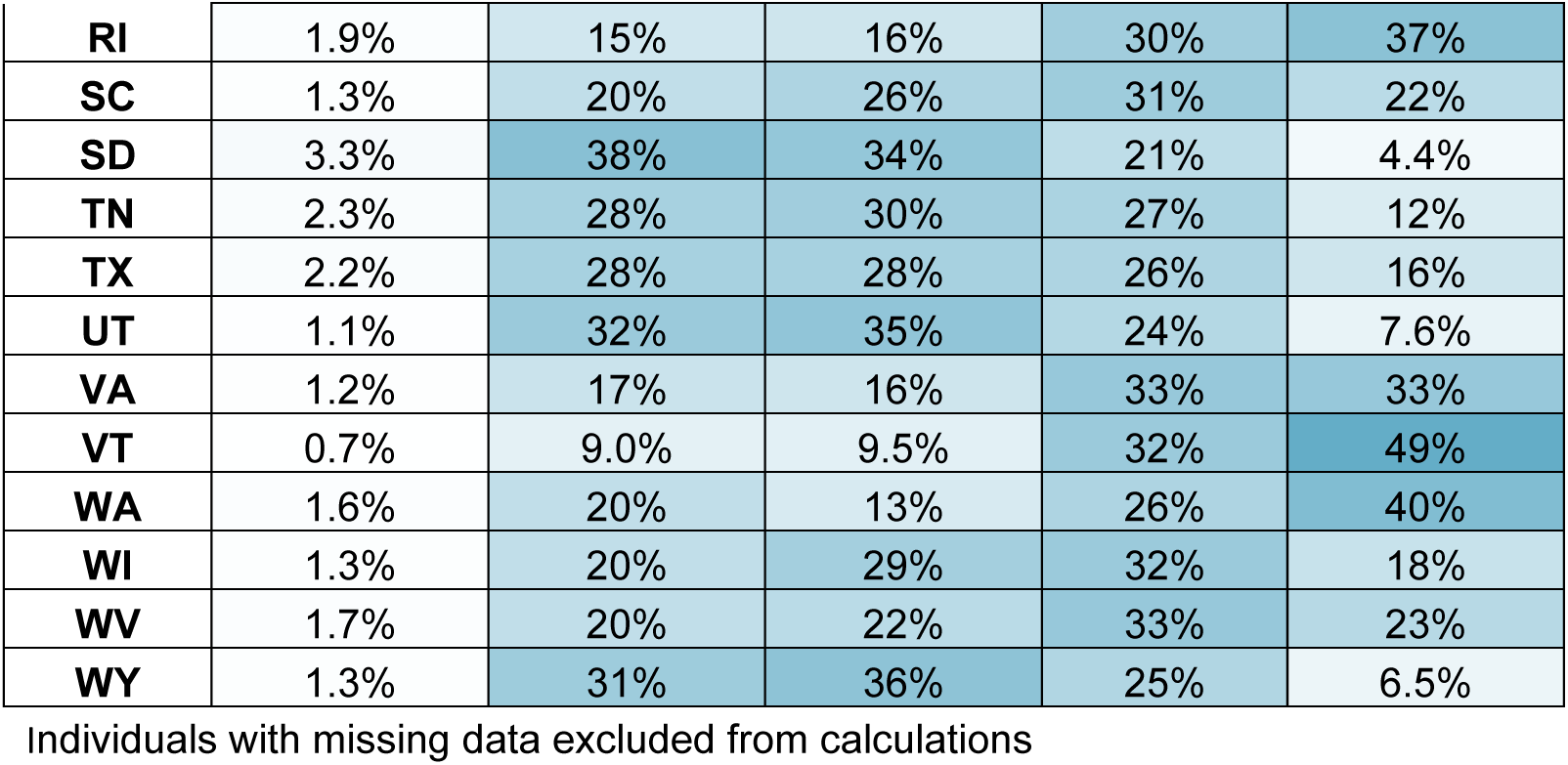
Survey-weighted percentage of participants with ≥1 school-aged child attending any in-person school (part-time or full-time) by the reported number of school mitigation measures and state.

**Table S7.**
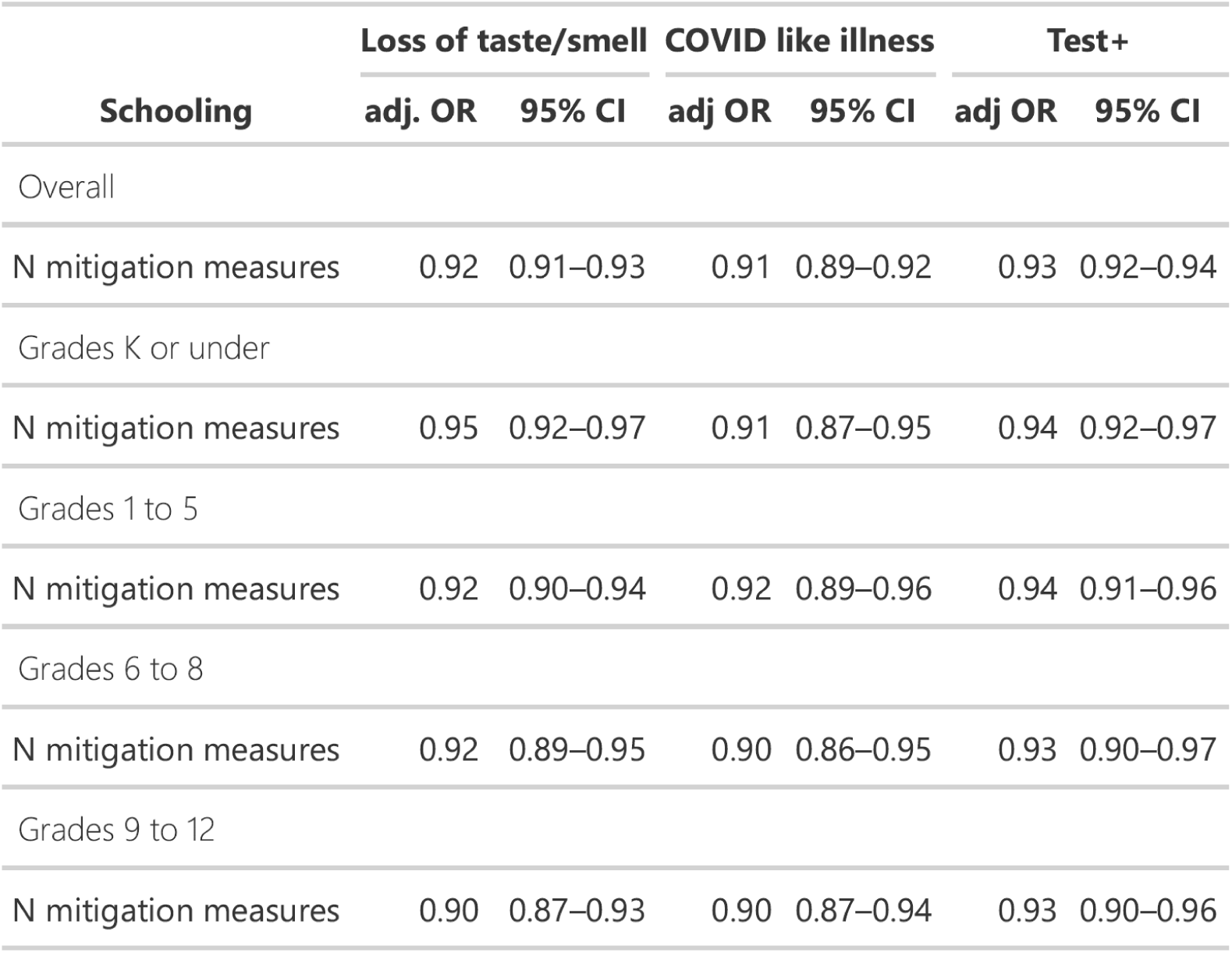
Association between risk of full-time schooling and number of mitigation measures implemented after accounting for survey design and adjusting for county and individual level covariates.

**Table S8.**
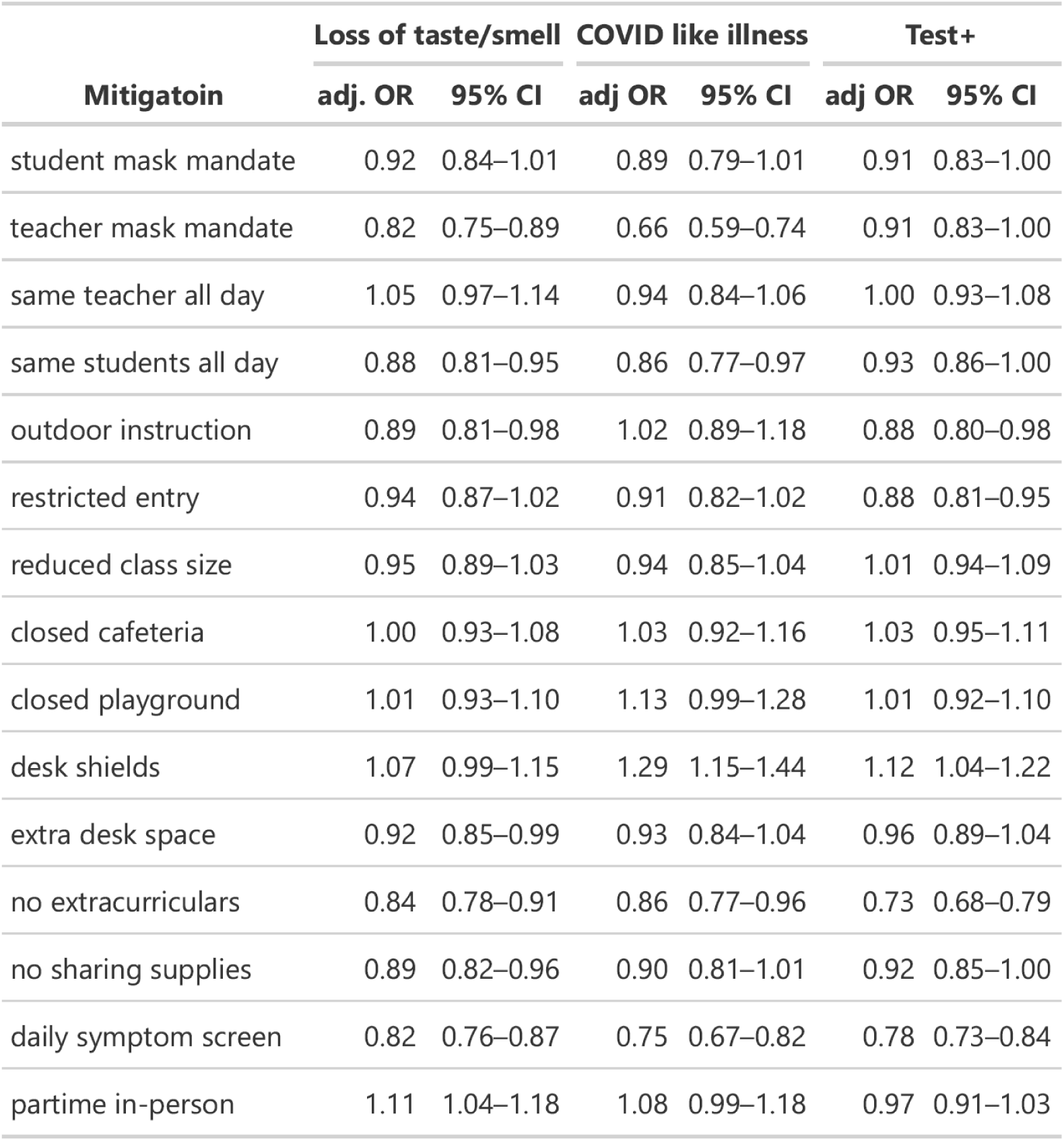
The association between mitigation measures and the off-ratio of COVID-19-related outcomes in those engaged in in-person schooling compared to those engaged in virtual- or home-schooling. Adjusted form county and individual level covariates.

**Table S9.**
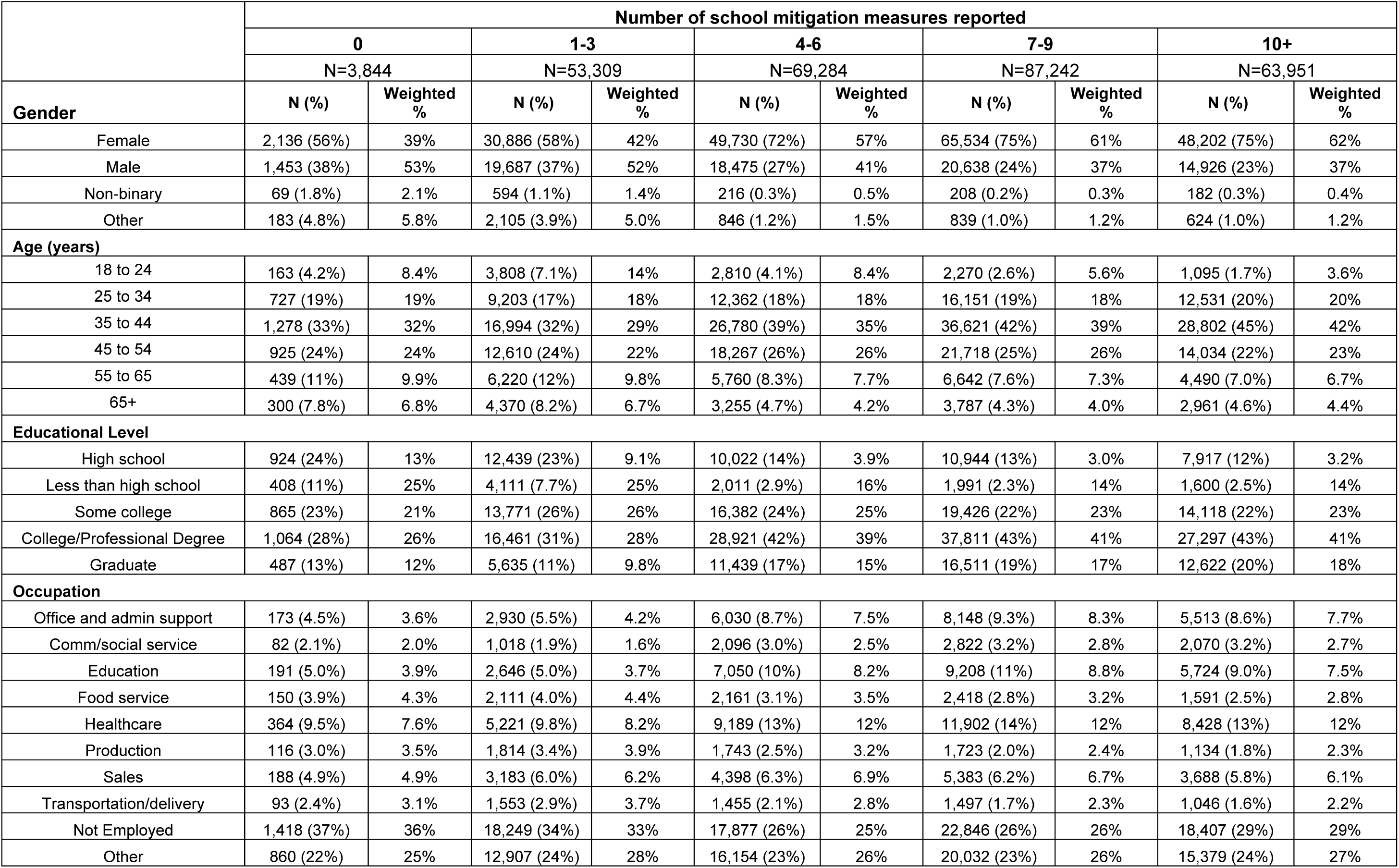
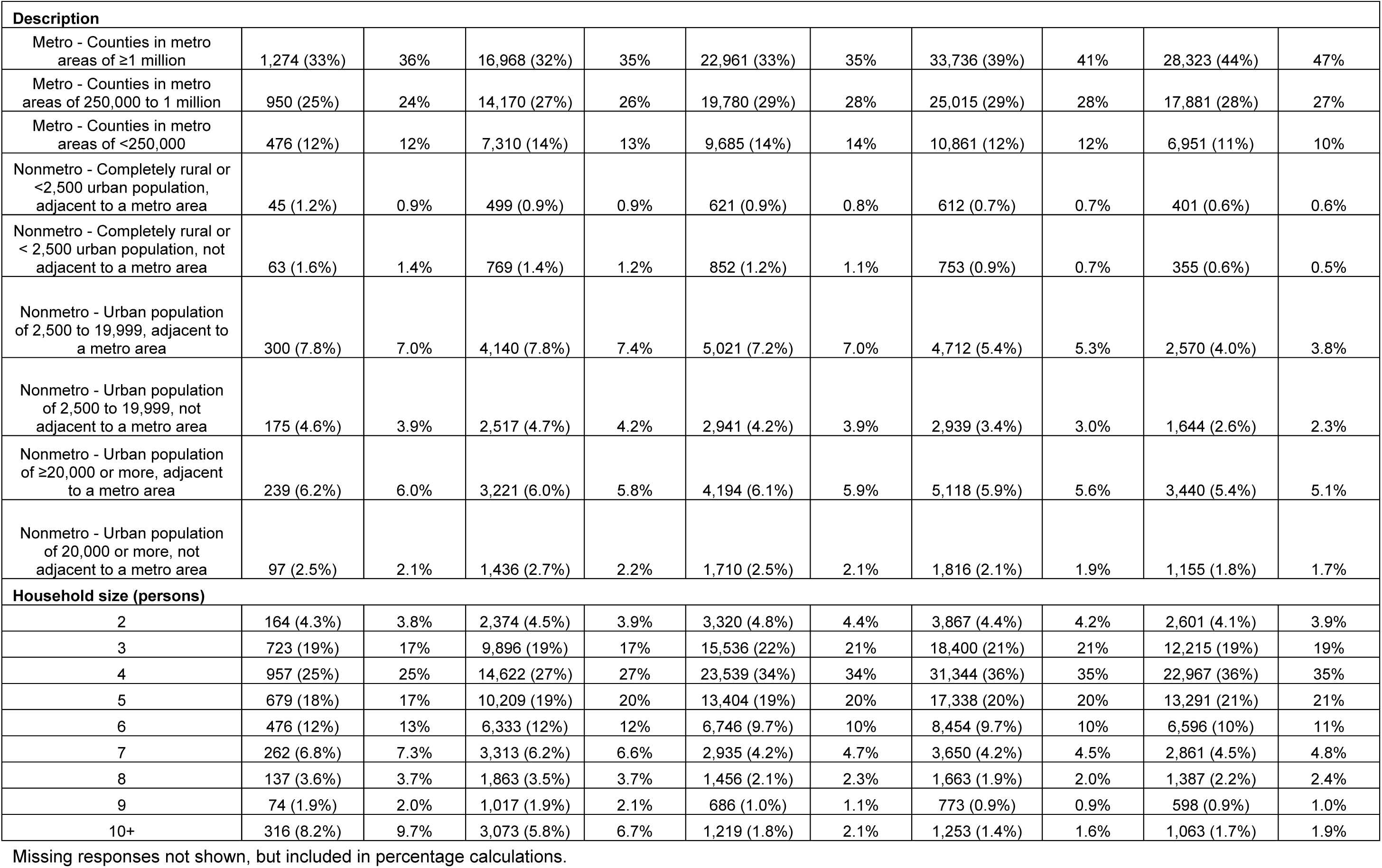
Selected demographics among participants with ≥1 school-aged child in the household attending any in-person schooling (part-time or full-time) by number of reported school mitigation measures; observed and survey-weighted percentages reported.

**Table S10.**
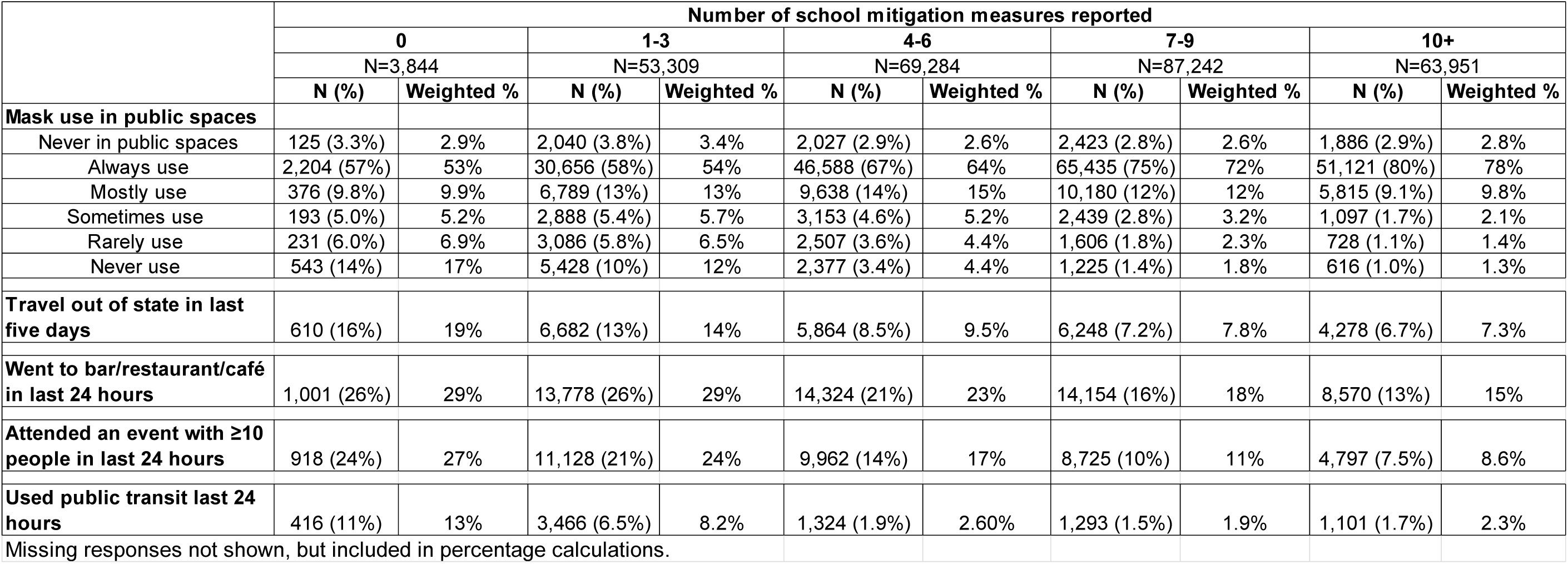
Selected behaviors relevant to COVID-19 acquisition/transmission among participants with ≥1 school-aged child in the household attending any in-person schooling (part-time or full-time) by number of reported school mitigation measures; observed and survey-weighted percentages reported.

**Table S11.**
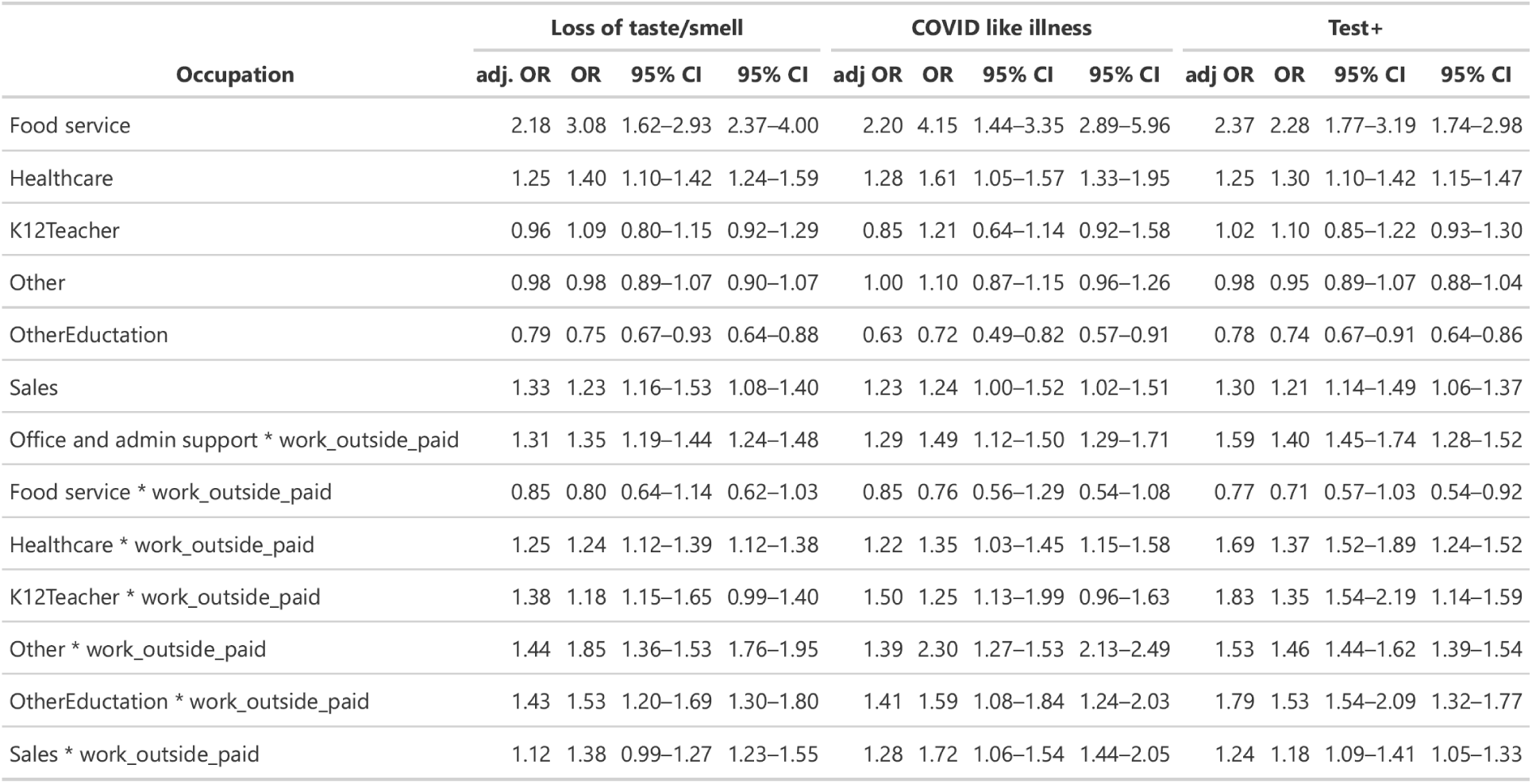
The relative odds, compared to office and administrative support staff not working outside the home for pay, of COVID-19-related outcomes among individuals who do, and do not, report paid work outside the home.

**Figure S1.**
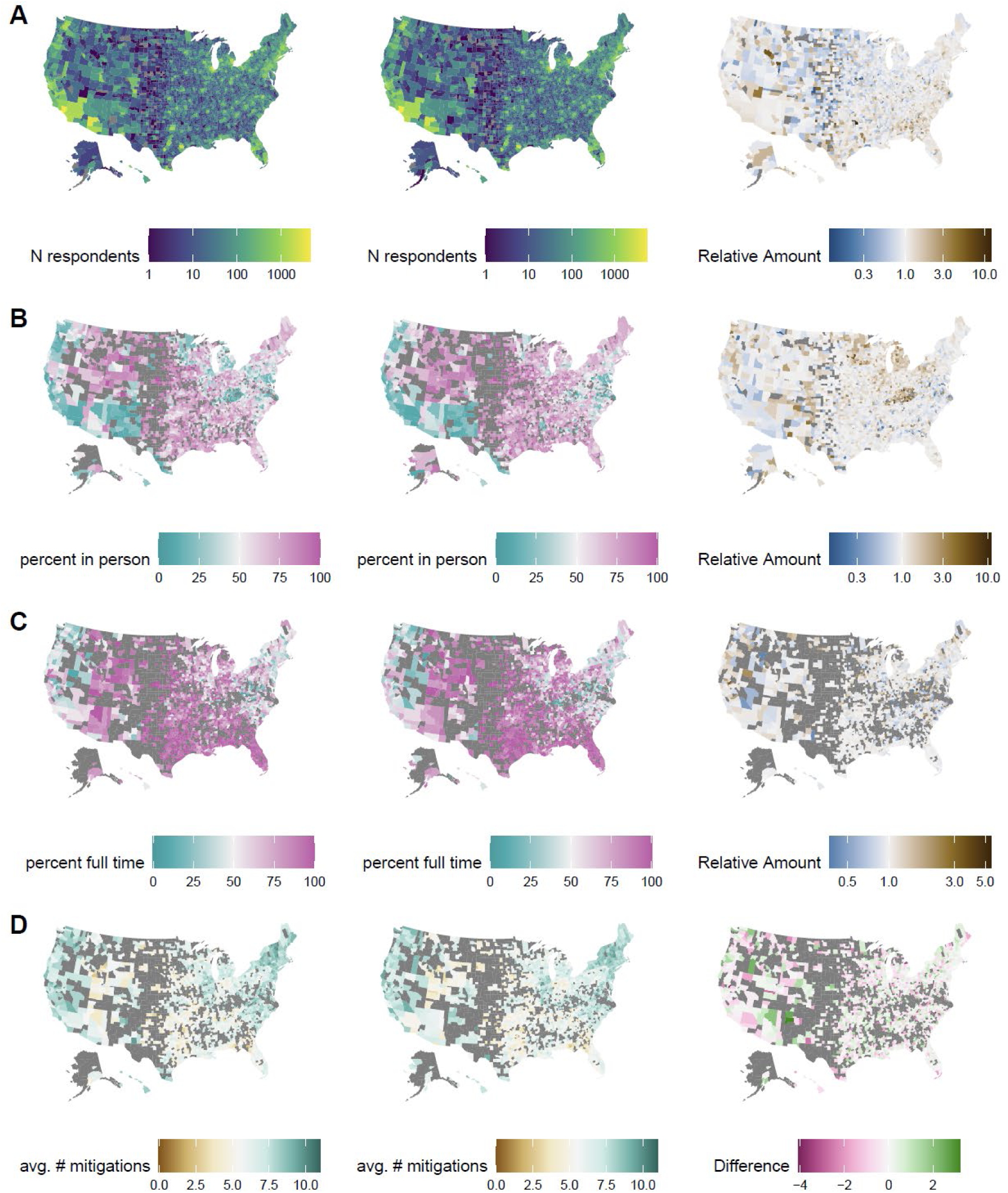
Distribution of outcomes in first period (left column), second period (center column) and change over time (right column). Results are shown for (**A**) number of survey respondents reporting ≧1 school-aged child in the household, (**B**) percent reporting in-person schooling, (**C**) percent reporting full-time in-person schooling, and (**D**) average number of in-school mitigation measures.

**Figure S2.**
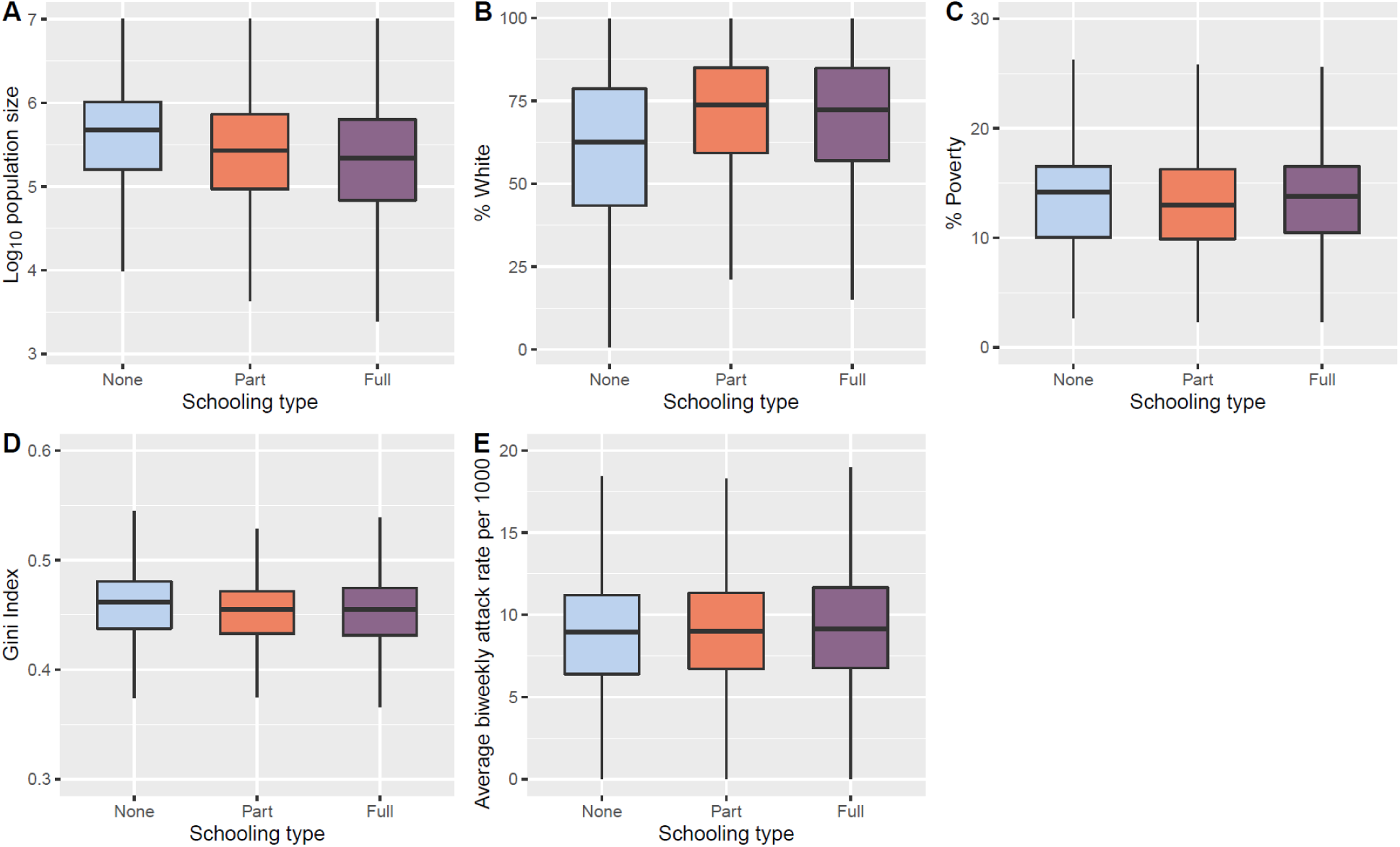
Distribution of selected county-level factors among participants reporting ≧1 school- aged child in the household by schooling type: no in-person schooling (none), part-time in-person schooling (Part), and full-time in-person schooling (Full); outlier values are excluded. Results are shown for county population size **(A)**, percentage of county population that is white **(B)** percentage with income level under the appropriate poverty threshold (**C**) Gini index of income inequality **(D)** and average biweekly SARS-CoV-2 attack rate per 1000 persons **(E)**.

**Figure S3.**
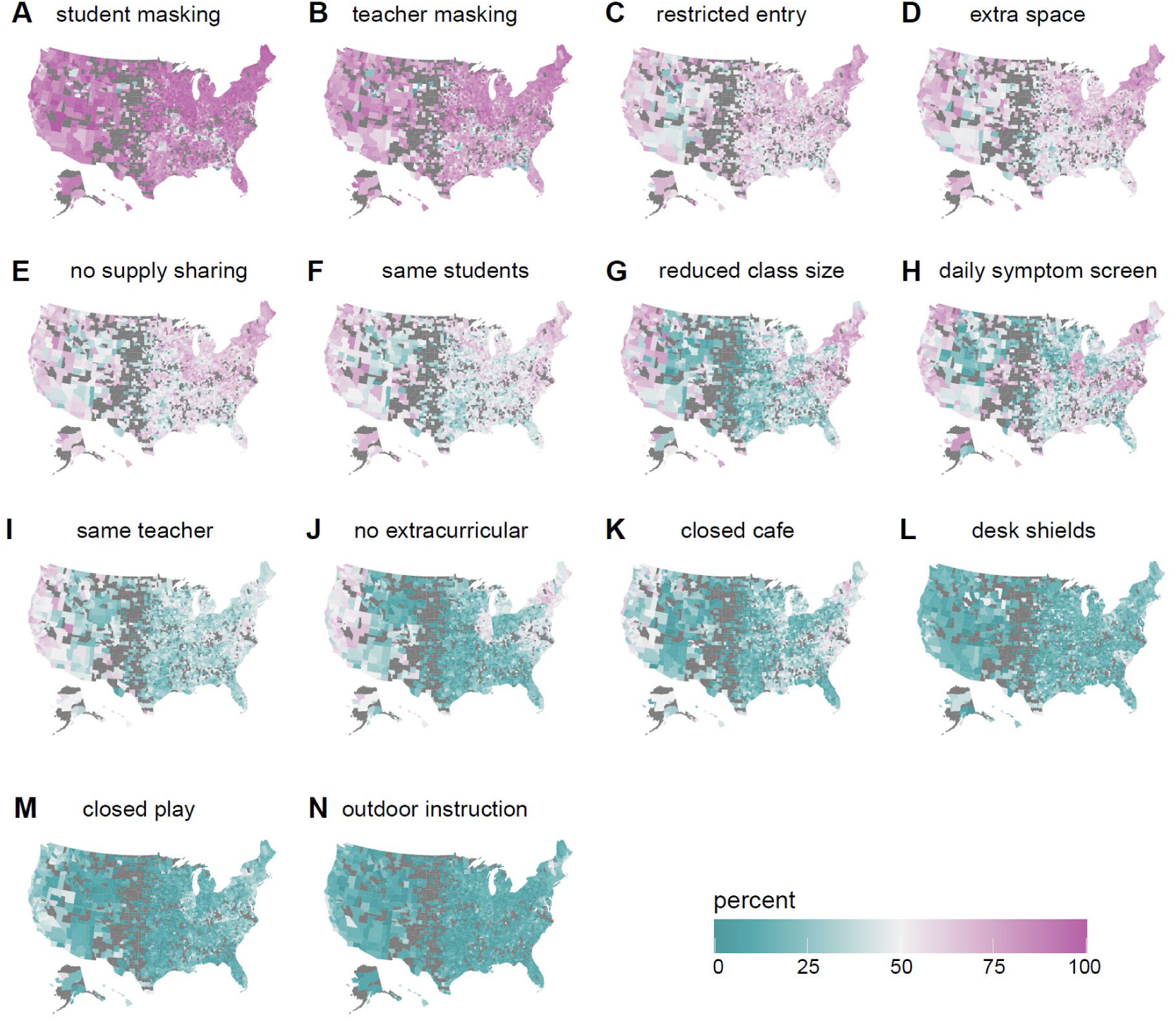
Percent of households with ≧1 child attending in-person school reporting each mitigation measure. Counties with less than 10 in-person respondents are excluded (gray).

**Figure S4.**
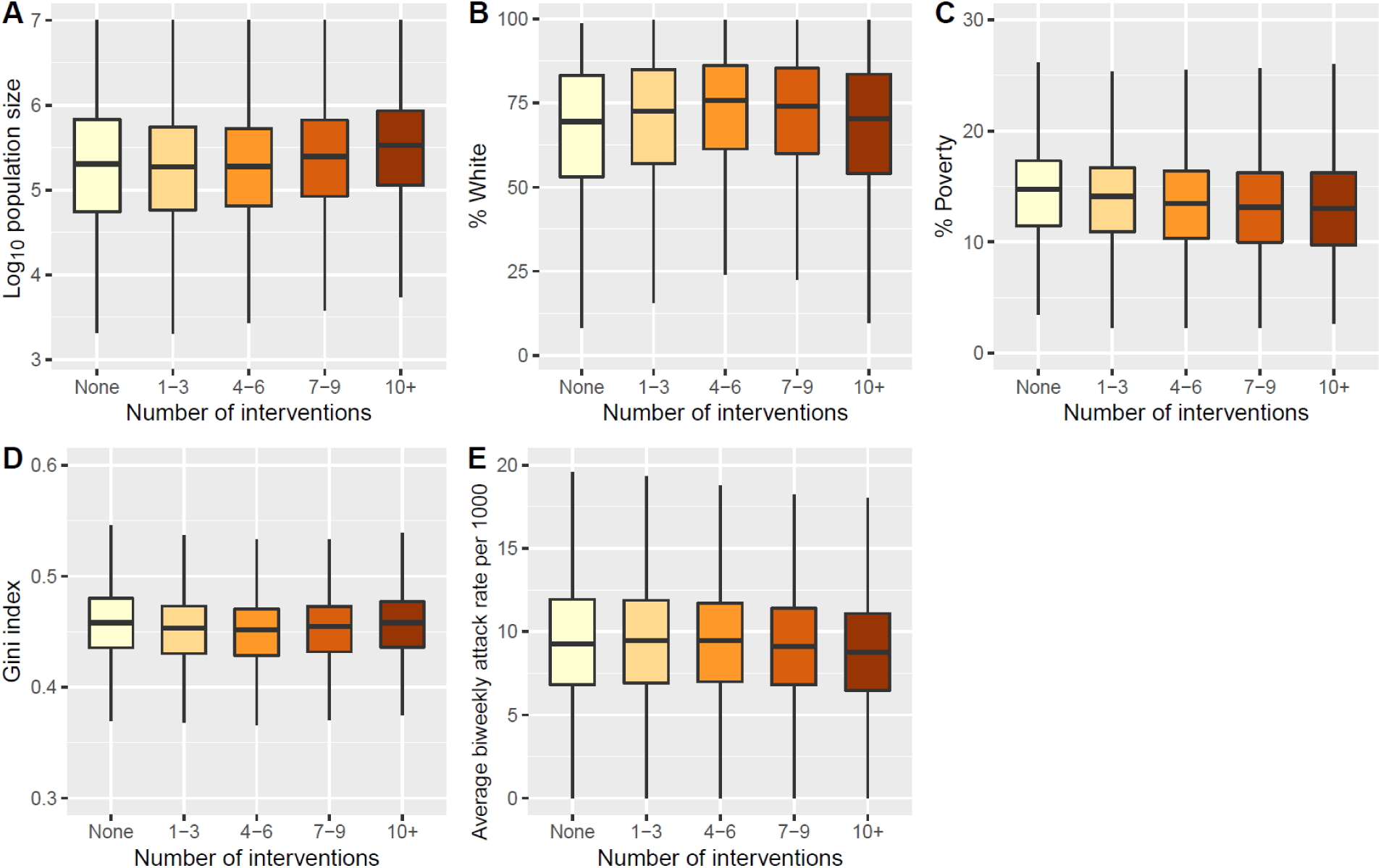
Distribution of selected county-level factors among participants reporting ≧1 school-aged child in the household attending in-person school (part-time or full-time) by the number of reported mitigation measures in the school; outlier values are excluded. Results are shown for county population size **(A)**, percentage of county population that is white **(B)**, percentage with income level under the appropriate poverty threshold (**C**) Gini index of income inequality **(D)** and average biweekly SARS-CoV-2 attack rate per 1000 persons **(E)**.

**Figure S5.**
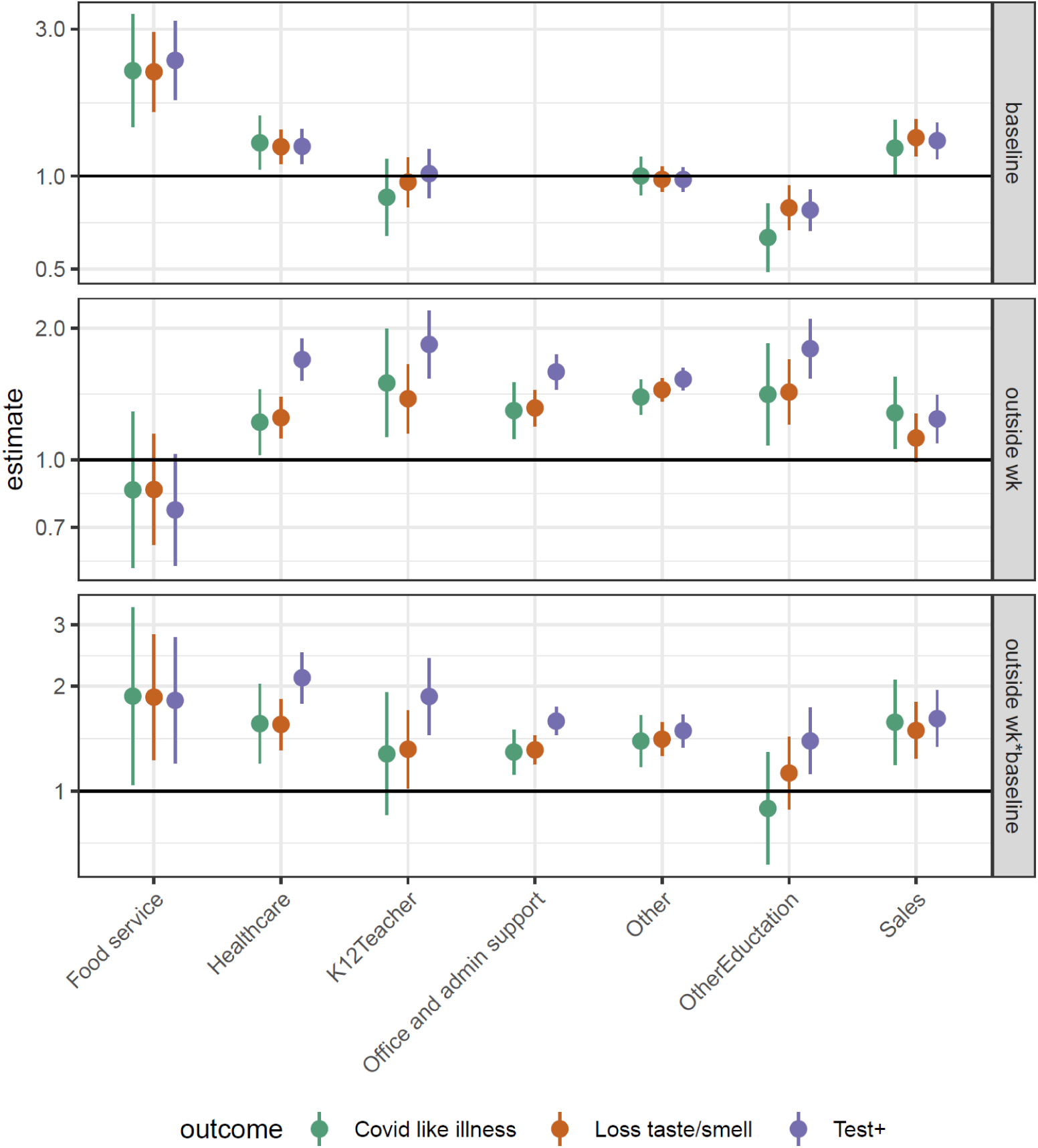
Odds ratio of COVID-19-related outcomes, contrasting office workers not reporting extra-household work for pay to those in other employment categories not reporting work for pay outside the home (top), and to those reporting work for pay outside the home (bottom). The middle row shows the odds ratio (i.e., increased risk) within each category associated with working outside the home compared to no work outside the home.

**Figure S6.**
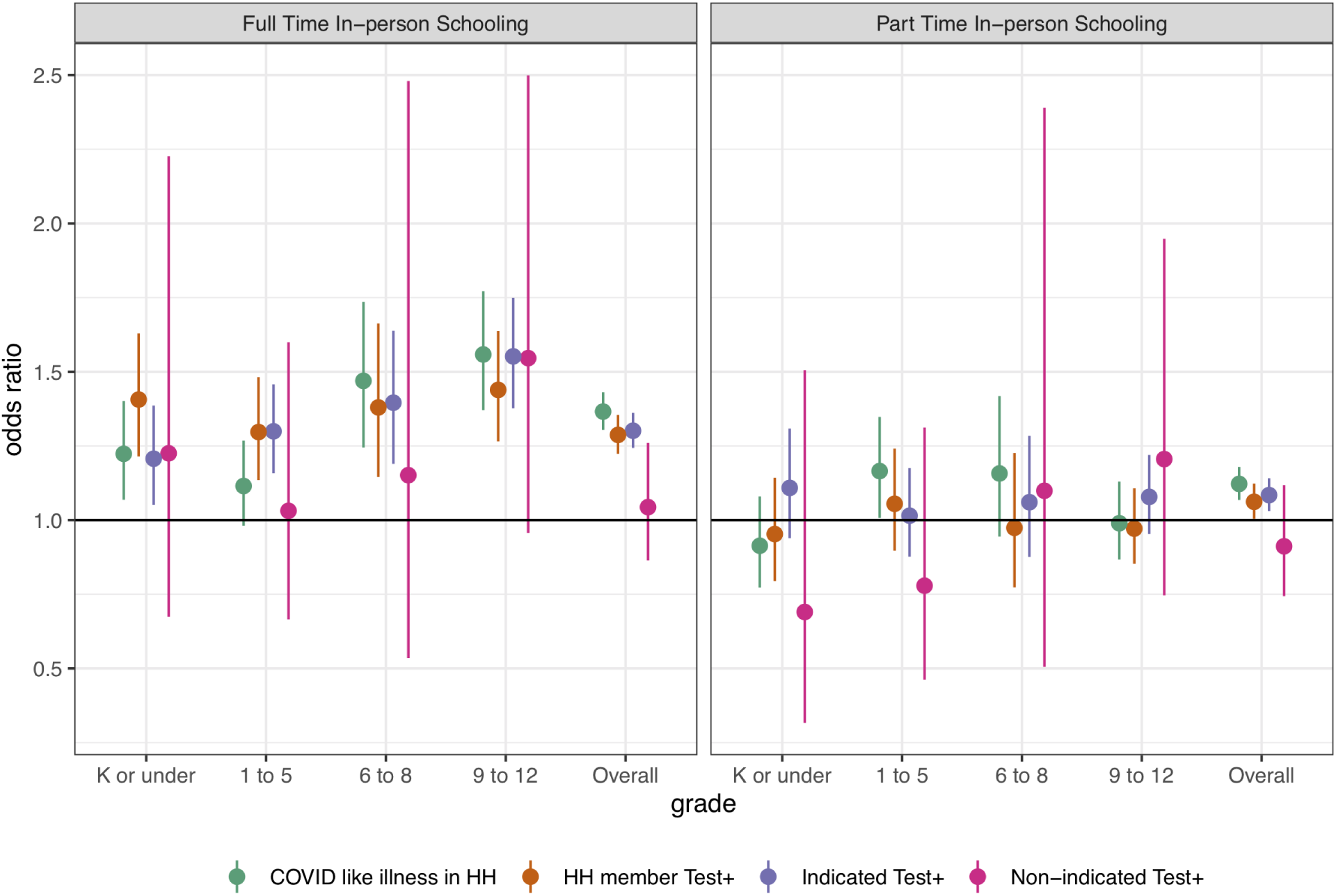
Odds ratio of secondary COVID-19-related outcomes associated with full- and part-time in-person schooling by outcome and grade level, adjusted for individual and county level covariates (but not number of mitigation measures).

**Figure S7.**
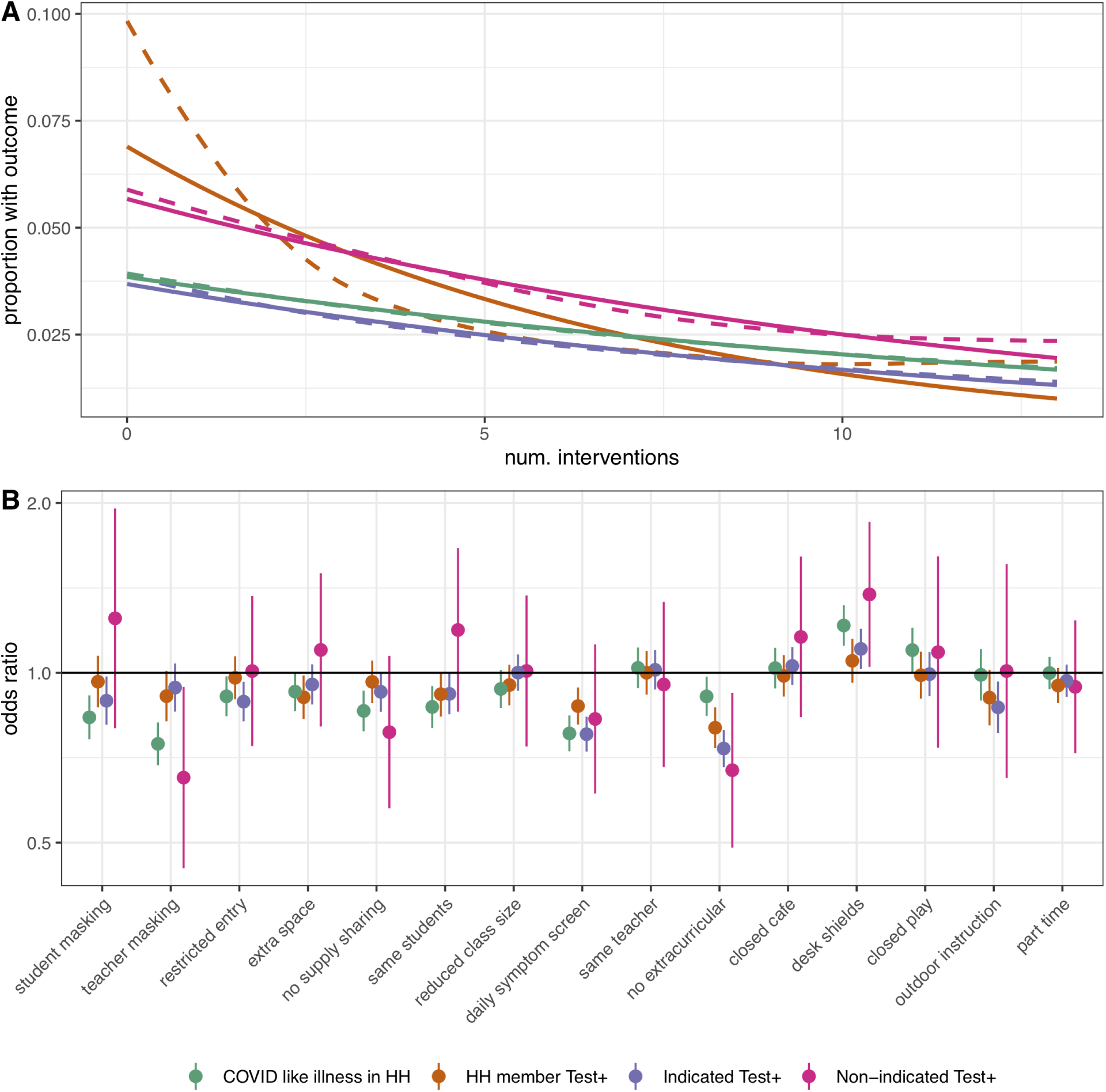
Relationship between number of mitigation measures and proportion reporting secondary COVID-19-related outcomes using a log-linear (solid) and spline (dashed) model. **(B)** Adjusted odds ratio of COVID-19-related outcomes by mitigation measure in multivariate model including all measures.

**Figure S8.**
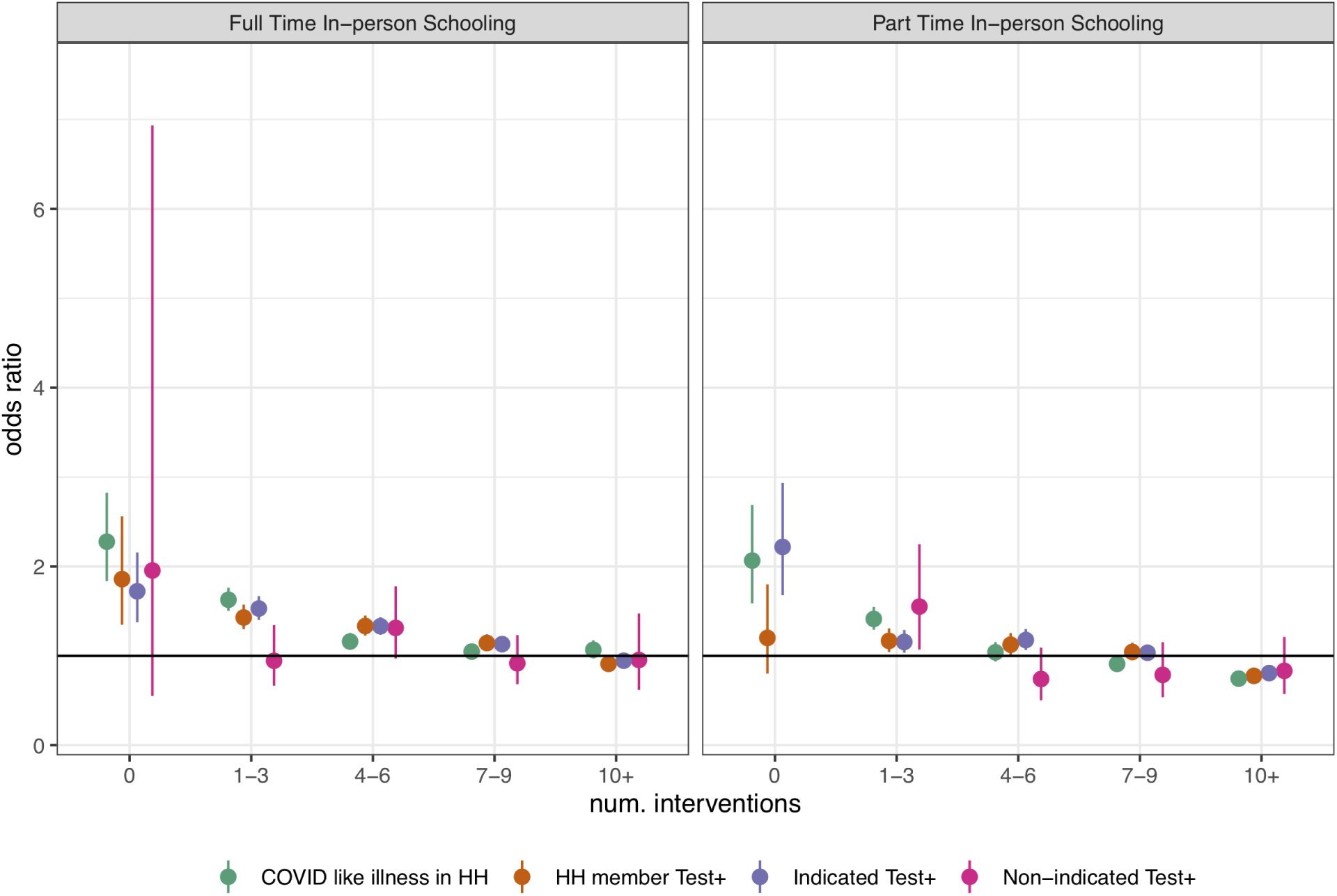
Odds ratio of secondary COVID-19-related outcomes associated with in-person full-time and part-time schooling by number of mitigation measures implemented, adjusted for individual and county-level covariates. An outlier value for the odds ratio of non-indicated positive test result among part-time in-person schooling with zero mitigation measures, for which the sample size was <30, is excluded.

**Figure S9.**
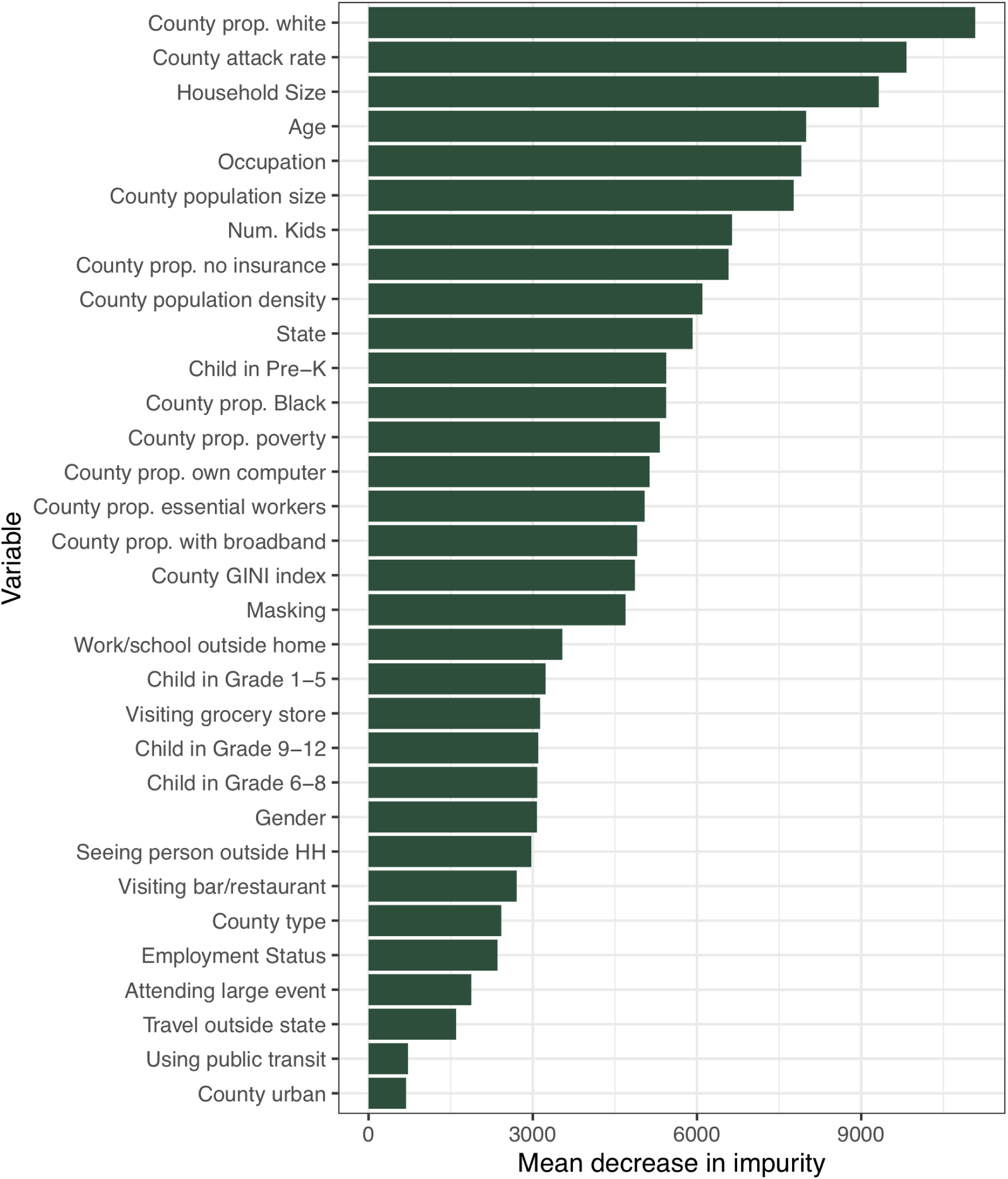
Variable importance, as mean decrease in impurity, for random forests of propensity for in-person schooling. Variables with greater influence on the propensity score have higher values.

## Notes

### Competing Interest Statement

The authors have declared no competing interest.

### Author Declarations

The original survey this analysis is based on reviewed and approved by the Institutional Review Boards of both the University of Maryland and Carnegie Mellon University

