## Supplementary Materials for "Household COVID-19 risk and in-person schooling"

**Figure S1.** Distribution of outcomes in first period (left column), second period (center column) and change over time (right column). Results are shown for (**A**) number of survey respondents reporting ≧1 school-aged child in the household, (**B**) percent reporting in-person schooling, (**C**) percent reporting full-time in-person schooling, and (**D**) average number of in-school mitigation measures.

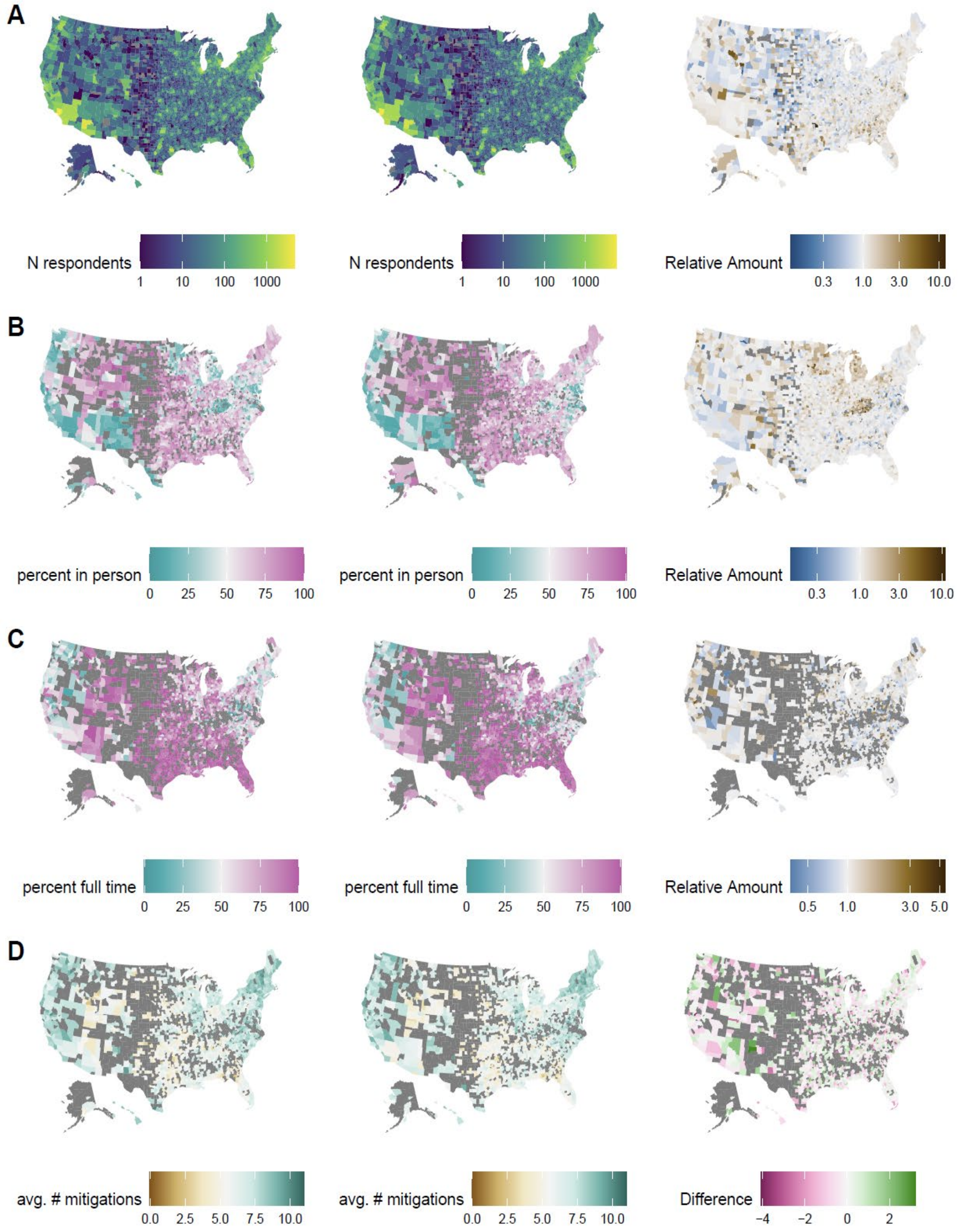

**Figure S2.** Distribution of selected county-level factors among participants reporting ≧1 school-aged child in the household by schooling type: no in-person schooling (none), part-time in-person schooling (Part), and full-time in-person schooling (Full); outlier values are excluded. Results are shown for county population size **(A)**, percentage of county population that is white **(B)** percentage with income level under the appropriate poverty threshold (**C**) Gini index of income inequality **(D)** and average biweekly SARS-CoV-2 attack rate per 1000 persons **(E)**.

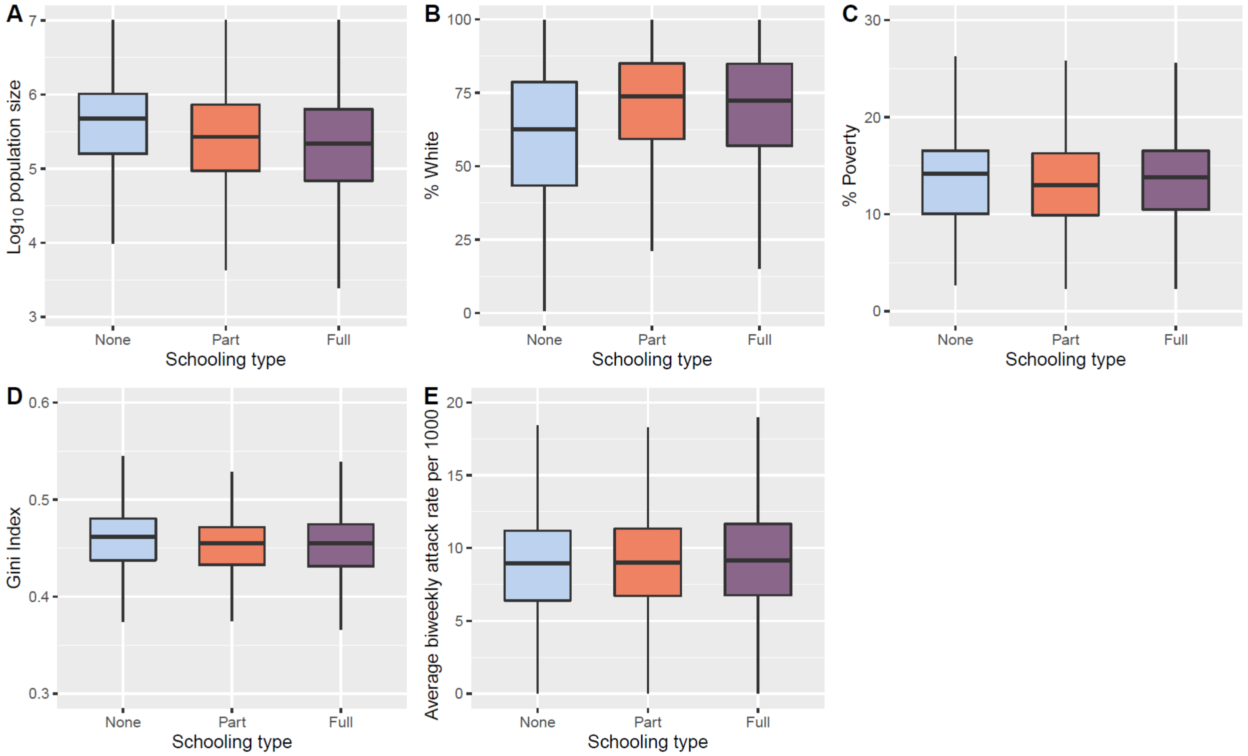

**Figure S3.** Percent of households with ≧1 child attending in-person school reporting each mitigation measure. Counties with less than 10 in-person respondents are excluded (gray).

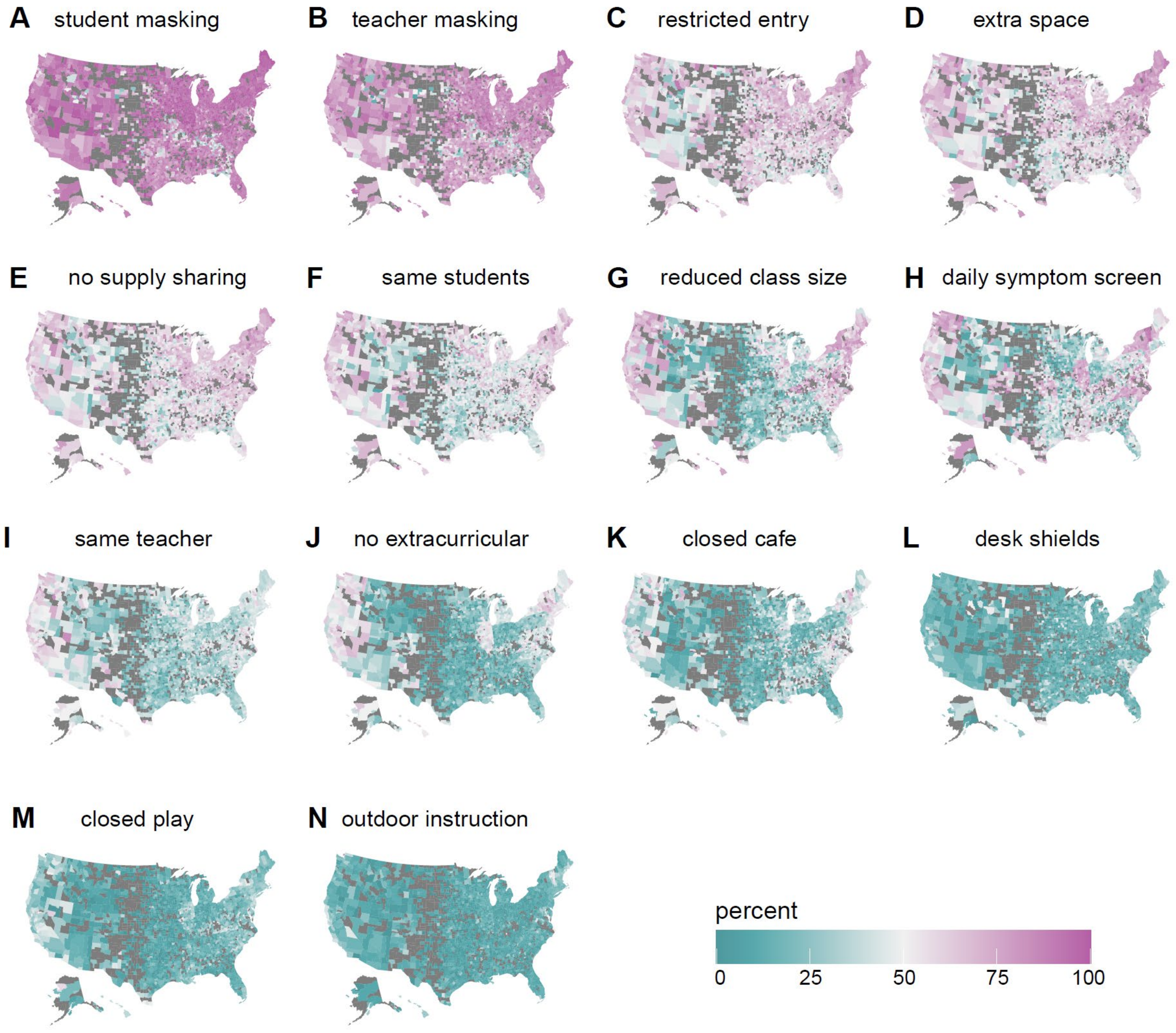

**Figure S4.** Distribution of selected county-level factors among participants reporting ≧1 school-aged child in the household attending in-person school (part-time or full-time) by the number of reported mitigation measures in the school; outlier values are excluded. Results are shown for county population size **(A)**, percentage of county population that is white **(B)**, percentage with income level under the appropriate poverty threshold (**C**) Gini index of income inequality **(D)** and average biweekly SARS-CoV-2 attack rate per 1000 persons **(E)**.

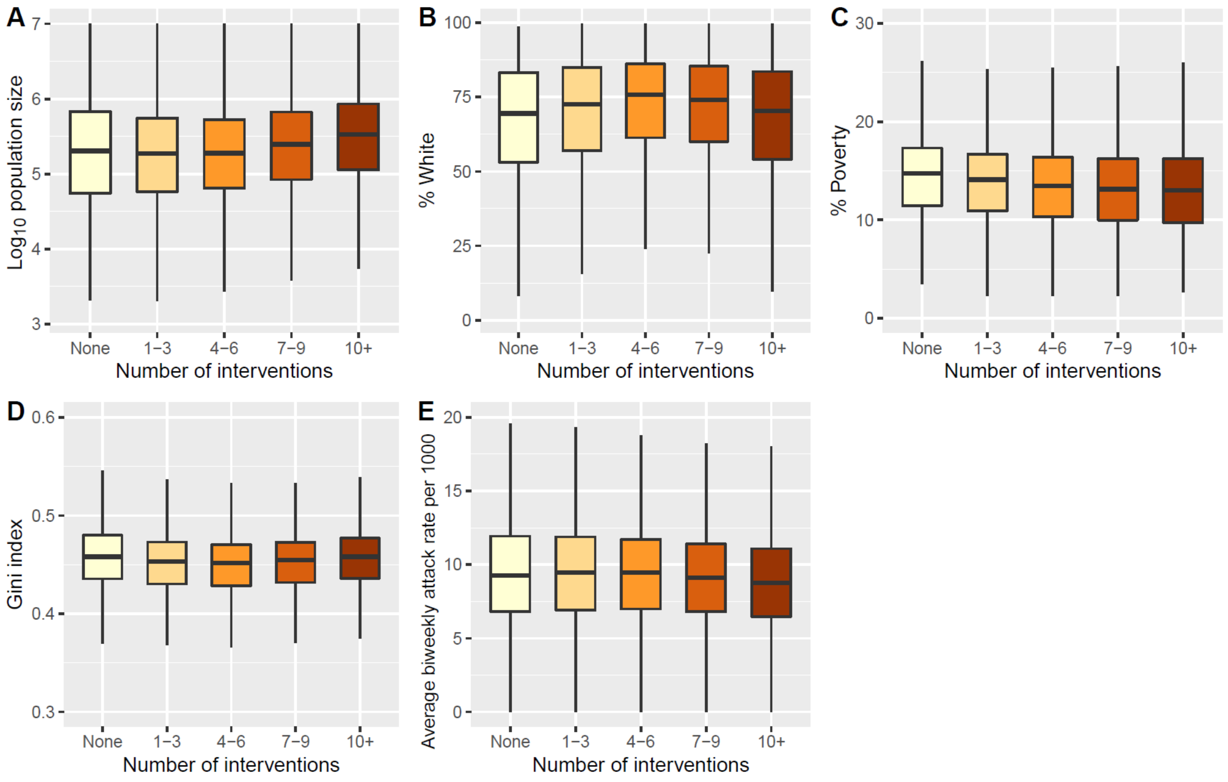

**Figure S5.** Odds ratio of COVID-19-related outcomes, contrasting office workers not reporting extra-household work for pay to those in other employment categories not reporting work for pay outside the home (top), and to those reporting work for pay outside the home (bottom). The middle row shows the odds ratio (i.e., increased risk) within each category associated with working outside the home compared to no work outside the home.

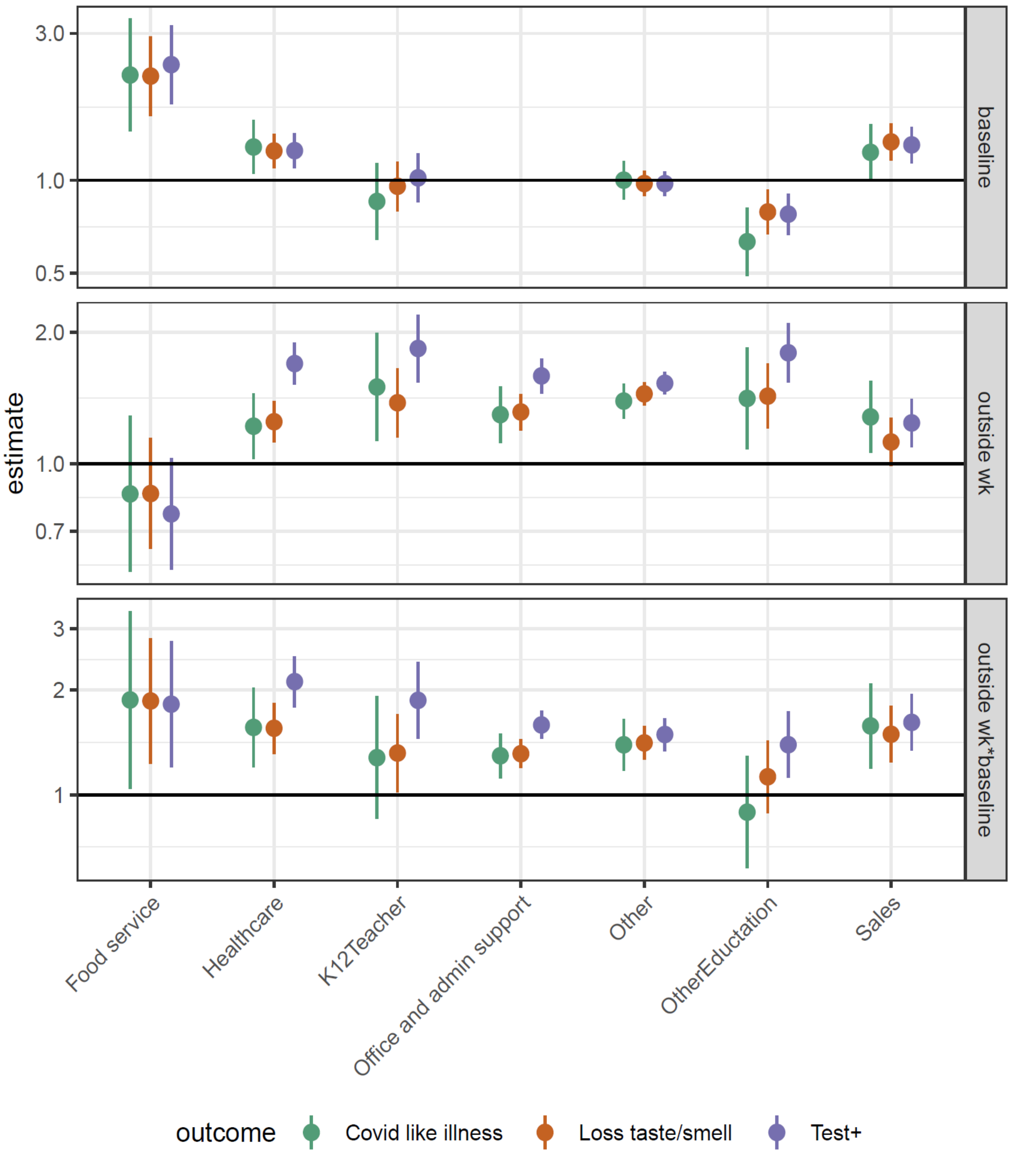

**Figure S6.** Odds ratio of secondary COVID-19-related outcomes associated with full- and part-time in-person schooling by outcome and grade level, adjusted for individual and county level covariates (but not number of mitigation measures).

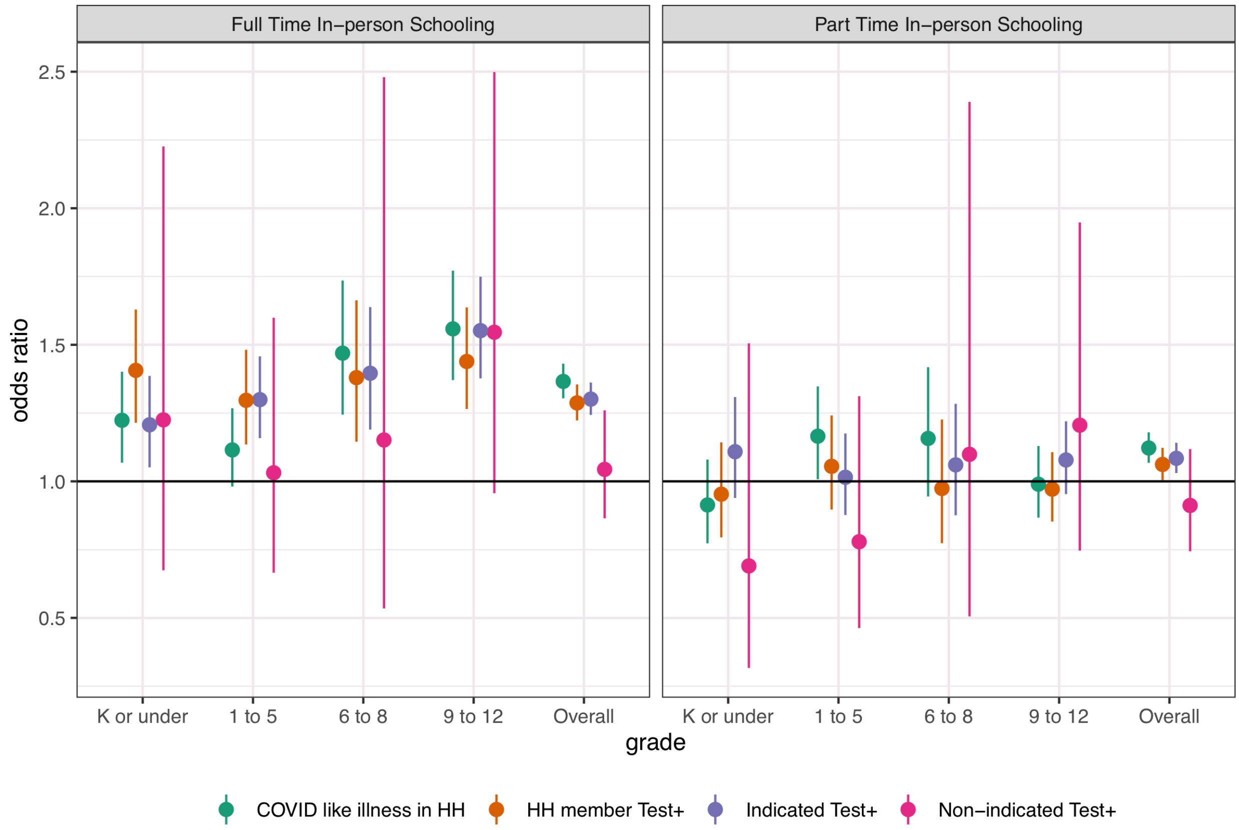

**Figure S7.** Relationship between number of mitigation measures and proportion reporting secondary COVID-19-related outcomes using a log-linear (solid) and spline (dashed) model. **(B)** Adjusted odds ratio of COVID-19-related outcomes by mitigation measure in multivariate model including all measures.

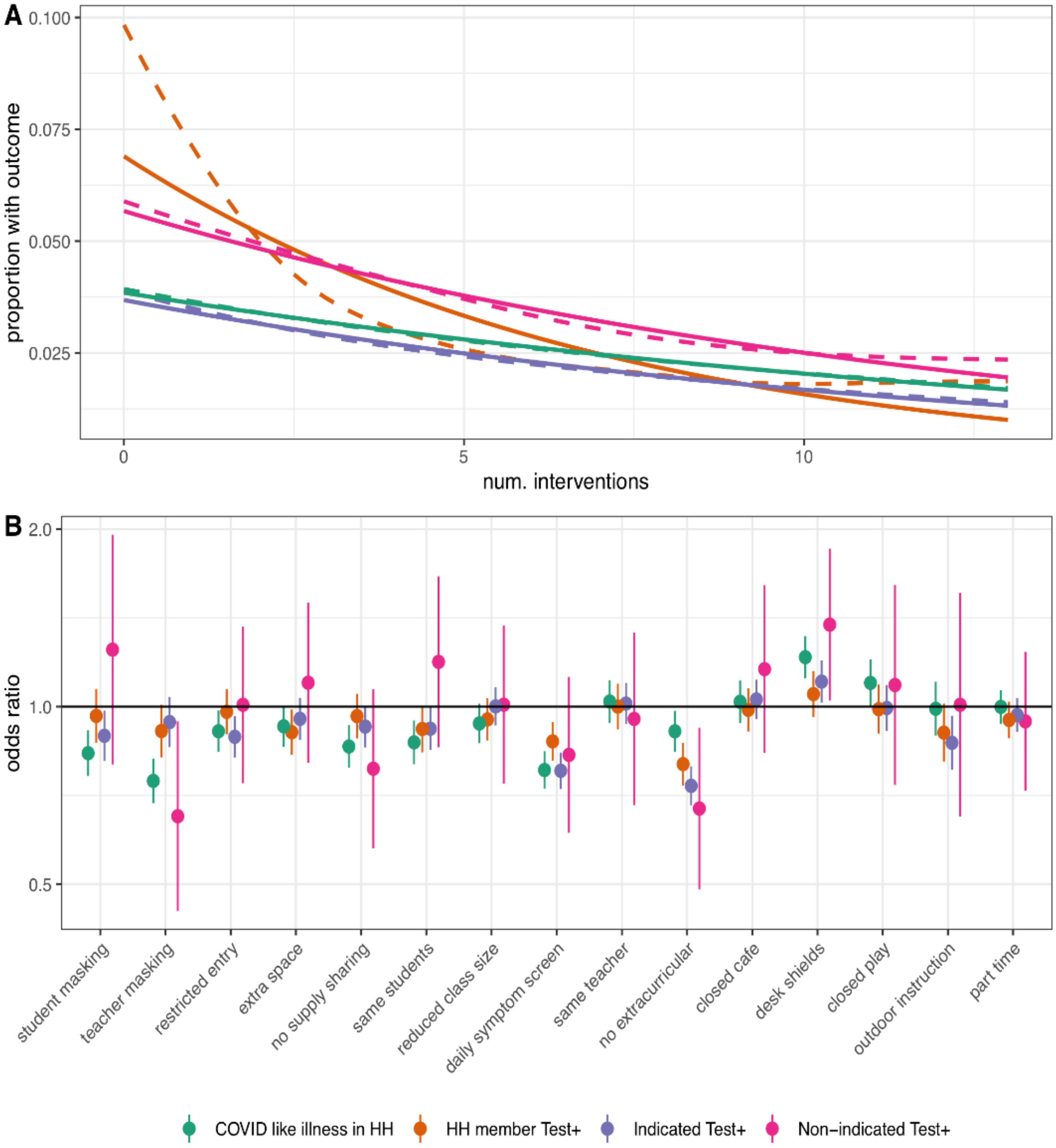

**Figure S8.** Odds ratio of secondary COVID-19-related outcomes associated with in-person full-time and part-time schooling by number of mitigation measures implemented, adjusted for individual and county-level covariates. An outlier value for the odds ratio of non-indicated positive test result among part-time in-person schooling with zero mitigation measures, for which the sample size was <30, is excluded.

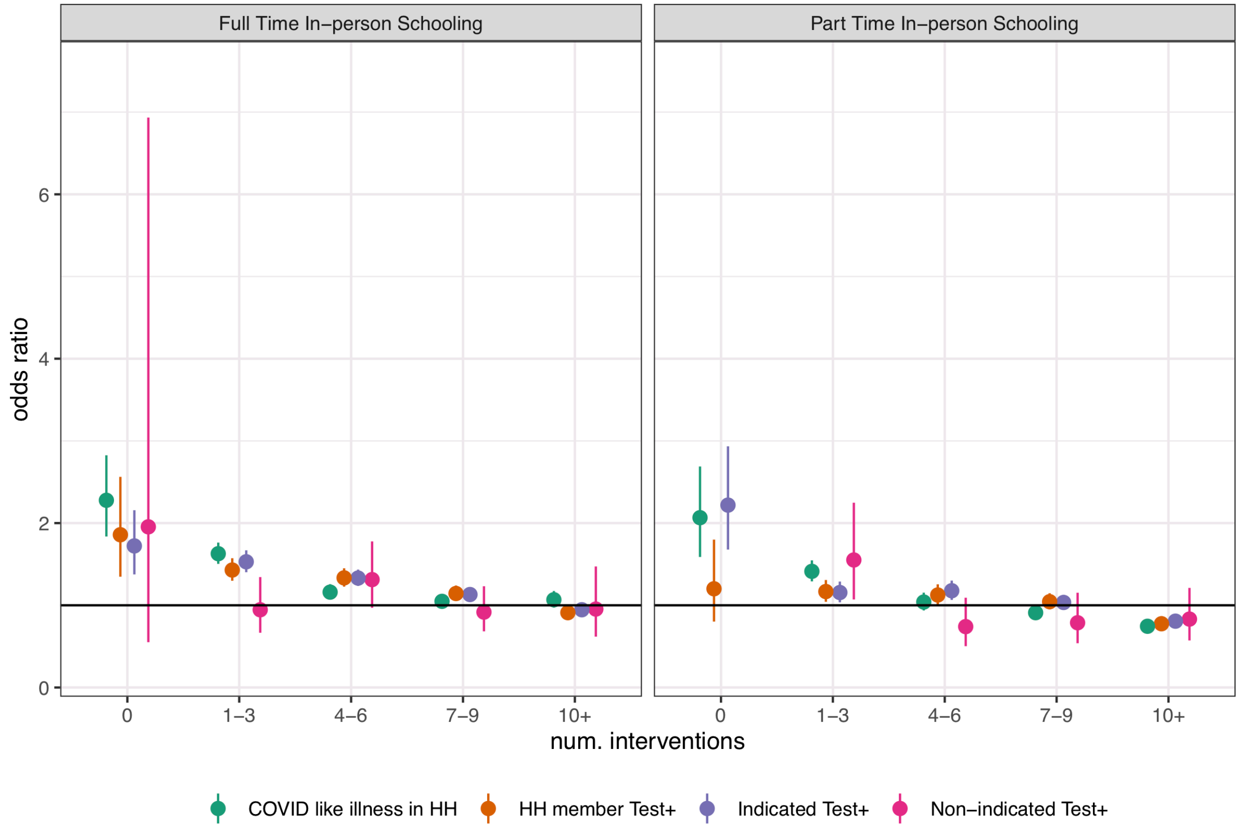

**Figure S9.** Variable importance, as mean decrease in impurity, for random forests of propensity for in-person schooling. Variables with greater influence on the propensity score have higher values.

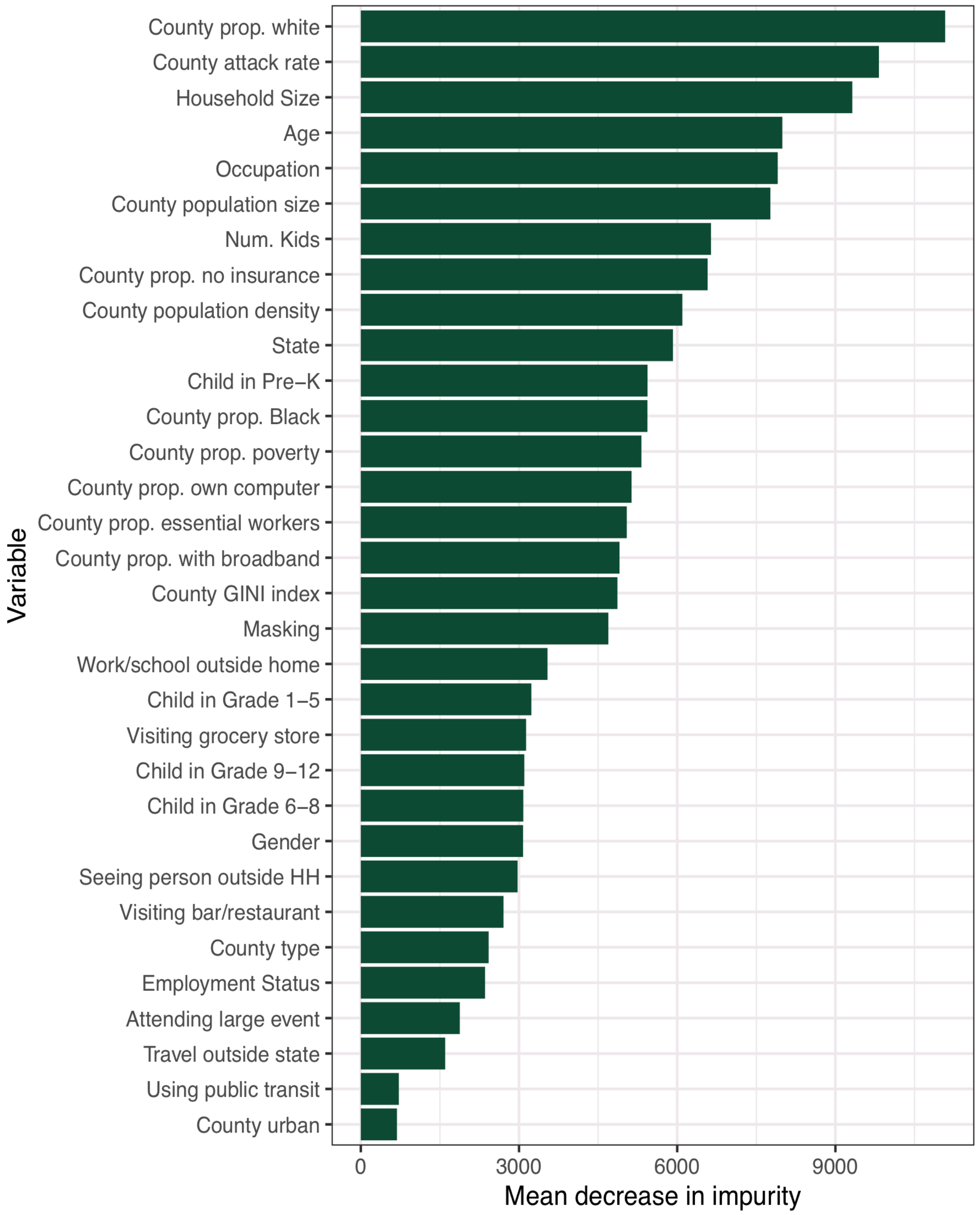

### Supplementary Tables

**Table S1.** Selected sociodemographics among participants with ≥1 school-aged child in the household comparing those reporting no in-person schooling to those reporting any part-time and full-time in-person schooling; observed and survey-weighted percentages reported.

| | No child attending in-person school | | Child attending in-person part-time school | | Child attending in-person full-time school | |
|---|---|---|---|---|---|---|
| | N=281,965 | | N=131,016 | | N=195,951 | |
| **Gender** | **N (%)** | **weighted %** | **N (%)** | **weighted %** | **N (%)** | **weighted %** |
| Female | 203,625 (72%) | 57% | 89,657 (68%) | 53% | 138,248 (71%) | 55% |
| Male | 72,817 (26%) | 40% | 37,485 (29%) | 43% | 52,427 (27%) | 41% |
| Non-binary | 1,338 (0.5%) | 0.6% | 900 (0.7%) | 1.0% | 1,100 (0.6%) | 0.8% |
| Other | 4,116 (1.5%) | 1.8% | 2,918 (2.2%) | 3.1% | 4,096 (2.1%) | 2.9% |
| **Age (years)** | | | | | | |
| 18 to 24 | 13,013 (4.6%) | 9.9% | 5,731 (4.4%) | 9.2% | 6,198 (3.2%) | 6.7% |
| 25 to 34 | 50,038 (18%) | 18% | 20,869 (16%) | 16% | 39,160 (20%) | 20% |
| 35 to 44 | 103,219 (37%) | 32% | 49,913 (38%) | 34% | 80,767 (41%) | 38% |
| 45 to 54 | 73,395 (26%) | 26% | 34,951 (27%) | 27% | 43,356 (22%) | 23% |
| 55 to 65 | 26,372 (9.4%) | 8.6% | 11,756 (9.0%) | 8.2% | 15,895 (8.1%) | 7.6% |
| 65+ | 15,658 (5.6%) | 4.9% | 7,635 (5.8%) | 5.4% | 10,371 (5.3%) | 5.0% |
| **Child in Pre-K or K** | 77,013 (27%) | 28% | 41,585 (32%) | 32% | 73,523 (38%) | 38% |
| **Child in Grades 1 - 5** | 125,974 (45%) | 44% | 61,977 (47%) | 47% | 105,605 (54%) | 54% |
| **Child in Grades 6 - 8** | 94,506 (34%) | 33% | 49,349 (38%) | 38% | 67,381 (34%) | 35% |
| **Child in Grades 9 - 12** | 113,395 (40%) | 41% | 60,771 (46%) | 48% | 68,117 (35%) | 36% |
| **Educational level** | | | | | | |
| Less than high school | 14,815 (5.3%) | 6.9% | 5,597 (4.3%) | 5.7% | 7,827 (4.0%) | 5.2% |
| High school | 50,990 (18%) | 21% | 20,355 (16%) | 18% | 30,642 (16%) | 18% |
| Some college | 72,454 (26%) | 27% | 30,264 (23%) | 24% | 45,520 (23%) | 37% |
| College/Professional Degree | 100,654 (36%) | 32% | 51,319 (39%) | 36% | 77,568 (40%) | 15% |
| Graduate | 39,655 (14%) | 12% | 22,225 (17%) | 15% | 32,490 (17%) | 1.1% |
| **Occupation** | | | | | | |
| Office and admin support | 19,842 (7.0%) | 5.9% | 10,048 (7.7%) | 6.4% | 15,692 (8.0%) | 6.7% |

| | | | | | | |
|---|---|---|---|---|---|---|
| Comm/social service | 7,045 (2.5%) | 2.1% | 3,776 (2.9%) | 2.4% | 5,592 (2.9%) | 2.4% |
| Education | 17,448 (6.2%) | 4.9% | 10,720 (8.2%) | 6.4% | 17,513 (8.9%) | 7.1% |
| Food service | 7,961 (2.8%) | 3.4% | 4,219 (3.2%) | 3.8% | 5,985 (3.1%) | 3.4% |
| Healthcare | 26,547 (9.4%) | 8.2% | 15,687 (12%) | 10% | 25,012 (13%) | 11% |
| Production | 5,522 (2.0%) | 2.4% | 3,087 (2.4%) | 2.9% | 4,828 (2.5%) | 3.1% |
| Sales | 14,186 (5.0%) | 5.3% | 7,843 (6.0%) | 6.4% | 11,805 (6.0%) | 6.4% |
| Transportation/delivery | 5,353 (1.9%) | 2.5% | 2,749 (2.1%) | 2.7% | 4,056 (2.1%) | 2.8% |
| Not Employed | 110,735 (39%) | 39% | 39,368 (30%) | 30% | 55,181 (28%) | 28% |
| Other | 61,338 (22%) | 24% | 30,918 (24%) | 26% | 46,221 (24%) | 27% |
| **Description of place of residence** | | | | | | |
| Metro - Counties in metro areas of ≥1 million | 141,406 (50%) | 53% | 51,048 (39%) | 41% | 69,724 (36%) | 37% |
| Metro - Counties in metro areas of 250,000 to 1 million | 77,174 (27%) | 26% | 37,137 (28%) | 27% | 54,101 (28%) | 27% |
| Metro - Counties in metro areas of <250,000 | 25,433 (9.0%) | 8.2% | 16,027 (12%) | 12% | 25,230 (13%) | 13% |
| Nonmetro - Completely rural or <2,500 urban population, adjacent to a metro area | 1,193 (0.4%) | 0.4% | 929 (0.7%) | 0.7% | 1,637 (0.8%) | 0.8% |
| Nonmetro - Completely rural or < 2,500 urban population, not adjacent to a metro area | 1,484 (0.5%) | 0.4% | 931 (0.7%) | 0.6% | 2,390 (1.2%) | 1.0% |
| Nonmetro - Urban population of 2,500 to 19,999, adjacent to a metro area | 8,879 (3.1%) | 2.8% | 6,535 (5.0%) | 4.8% | 13,199 (6.7%) | 6.6% |
| Nonmetro - Urban population of 2,500 to 19,999, not adjacent to a metro area | 5,354 (1.9%) | 1.6% | 3,806 (2.9%) | 2.6% | 8,261 (4.2%) | 3.9% |
| Nonmetro - Urban population of ≥20,000 or more, adjacent to a metro area | 10,810 (3.8%) | 3.5% | 8,060 (6.2%) | 5.8% | 11,136 (5.7%) | 5.5% |
| Nonmetro - Urban population of 20,000 or more, not adjacent to a metro area | 4,011 (1.4%) | 1.2% | 2,660 (2.0%) | 1.8% | 4,579 (2.3%) | 2.0% |
| **Household size (number of persons)** | | | | | | |

| | | | | | | |
|---|---|---|---|---|---|---|
| 2 | 14,574 (5.2%) | 4.6% | 5,144 (3.9%) | 3.6% | 8,461 (4.3%) | 4.0% |
| 3 | 61,294 (22%) | 20% | 23,805 (18%) | 17% | 38,524 (20%) | 19% |
| 4 | 88,850 (32%) | 30% | 43,743 (33%) | 32% | 64,255 (33%) | 32% |
| 5 | 53,575 (19%) | 20% | 26,681 (20%) | 20% | 39,271 (20%) | 20% |
| 6 | 29,346 (10%) | 11% | 14,232 (11%) | 11% | 21,026 (11%) | 11% |
| 7 | 14,063 (5.0%) | 5.5% | 6,675 (5.1%) | 5.6% | 9,604 (4.9%) | 5.3% |
| 8 | 7,433 (2.6%) | 3.0% | 3,453 (2.6%) | 2.9% | 4,853 (2.5%) | 2.8% |
| 9 | 3,799 (1.3%) | 1.6% | 1,726 (1.3%) | 1.5% | 2,342 (1.2%) | 1.3% |
| 10+ | 6,724 (2.4%) | 2.8% | 4,436 (3.4%) | 4.4% | 6,055 (3.1%) | 4.0% |

Missing responses not shown, but included in percentage calculations.

**Table S2.** Selected behaviors relevant to COVID-19 acquisition/transmission and COVID-19-related outcomes among participants with ≥1 school-aged child in the household comparing those reporting no in-person schooling to those reporting any part-time and full-time in-person schooling; observed and survey-weighted percentages reported.

| | **No child attending in-person school** | | **Child attending in-person part-time school** | | **Child attending in-person full-time school** | |
|---|---|---|---|---|---|---|
| | **N=281,965** | | **N=131,016** | | **N=195,951** | |
| **Mask use in public spaces** | **N (%)** | **weighted %** | **N (%)** | **weighted %** | **N (%)** | **weighted %** |
| Never in public spaces | 16,359 (5.8%) | 5.2% | 4,093 (3.1%) | 2.9% | 5,877 (3.0%) | 2.7% |
| Always use | 222,692 (79%) | 77% | 94,913 (72%) | 68% | 133,177 (68%) | 64% |
| Mostly use | 21,079 (7.5%) | 8.3% | 14,092 (11%) | 12% | 24,104 (12%) | 13% |
| Sometimes use | 4,683 (1.7%) | 1.9% | 4,044 (3.1%) | 3.6% | 7,644 (3.9%) | 4.5% |
| Rarely use | 3,456 (1.2%) | 1.5% | 3,330 (2.5%) | 3.2% | 6,562 (3.3%) | 4.1% |
| Never use | 3,818 (1.4%) | 1.6% | 4,864 (3.7%) | 5.1% | 9,854 (5.0%) | 6.9% |
| **Travel out of state in last five days** | 15,515 (5.5%) | 5.9% | 12,433 (9.5%) | 11% | 18,150 (9.3%) | 11% |
| **Went to bar/restaurant/café in last 24 hours** | 26,423 (9.4%) | 10% | 23,362 (18%) | 21% | 40,894 (21%) | 24% |
| **Attended an event with ≥10 people in last 24 hours** | 16,513 (5.9%) | 6.8% | 16,034 (12%) | 15% | 29,448 (15%) | 18% |
| **Used public transit in last 24 hours** | 6,235 (2.2%) | 2.9% | 4,899 (3.7%) | 5.3% | 6,990 (3.6%) | 5.1% |
| **COVID-19 like illness†** | 3,536 (1.3%) | 1.3% | 2,878 (2.2%) | 2.8% | 4,387 (2.2%) | 2.8% |
| **Loss of taste/smell†** | 8,488 (3.0%) | 3.1% | 4,937 (3.8%) | 4.3% | 7,660 (3.9%) | 4.4% |
| **Tested for SARS-CoV-2‡** | 41,203 (15%) | 15% | 20,006 (15%) | 16% | 29,537 (15%) | 15% |
| **Tested positive for SARS-CoV-2‡** | 8,412 (3.0%) | 3.1% | 3,705 (2.8%) | 2.9% | 6,113 (3.1%) | 3.2% |

† Symptoms reported over the last 24 hours; ‡ Testing status/results reported over last 14 days

Missing responses not shown, but included in percentage calculations.

**Table S3.** Survey-weighted percentage of participants with ≥1 school-aged child in the household by reported schooling type and state.

| State | Calendar period 1: Nov-Dec 2020 | | | Calendar period 2: Jan-Feb 2021 | | |
|---|---|---|---|---|---|---|
| | No child attending in-person school | Child attending part-time in-person school | Child attending full-time in-person school | No child attending in-person school | Child attending part-time in-person school | Child attending full-time in-person school |
| AK | 62 | 18 | 28 | 45 | 25 | 40 |
| AL | 32 | 23 | 56 | 30 | 24 | 56 |
| AR | 30 | 23 | 59 | 29 | 20 | 61 |
| AZ | 53 | 19 | 33 | 61 | 15 | 28 |
| CA | 74 | 15 | 12 | 75 | 14 | 12 |
| CO | 63 | 18 | 24 | 35 | 34 | 42 |
| CT | 41 | 35 | 30 | 35 | 39 | 35 |
| DC | 61 | 20 | 28 | 52 | 31 | 31 |
| DE | 54 | 29 | 19 | 45 | 41 | 21 |
| FL | 39 | 12 | 55 | 34 | 13 | 60 |
| GA | 39 | 16 | 52 | 39 | 17 | 52 |
| HI | 56 | 24 | 23 | 48 | 34 | 22 |
| IA | 32 | 29 | 49 | 21 | 27 | 62 |
| ID | 30 | 37 | 44 | 22 | 44 | 49 |
| IL | 65 | 19 | 19 | 49 | 31 | 25 |
| IN | 42 | 22 | 44 | 29 | 27 | 55 |
| KS | 43 | 22 | 43 | 29 | 29 | 55 |
| KY | 76 | 14 | 11 | 49 | 32 | 24 |
| LA | 24 | 22 | 64 | 23 | 21 | 66 |
| MA | 47 | 40 | 17 | 46 | 40 | 19 |
| MD | 81 | 8.1 | 10 | 79 | 9 | 11 |
| ME | 29 | 51 | 27 | 27 | 50 | 31 |
| MI | 67 | 14 | 22 | 44 | 25 | 38 |
| MN | 69 | 16 | 19 | 46 | 28 | 36 |
| MO | 33 | 24 | 52 | 28 | 23 | 58 |
| MS | 33 | 22 | 54 | 26 | 22 | 61 |
| MT | 28 | 27 | 54 | 24 | 24 | 60 |
| NC | 51 | 31 | 24 | 53 | 30 | 23 |
| ND | 25 | 29 | 55 | 22 | 21 | 67 |
| NE | 22 | 24 | 63 | 21 | 22 | 68 |
| NH | 44 | 33 | 29 | 38 | 37 | 32 |
| NJ | 61 | 27 | 14 | 58 | 30 | 15 |
| NM | 85 | 8 | 7 | 78 | 11 | 10 |
| NV | 67 | 19 | 18 | 64 | 21 | 19 |

| | | | | | | |
|---|---|---|---|---|---|---|
| NY | 44 | 39 | 24 | 41 | 40 | 26 |
| OH | 48 | 25 | 33 | 34 | 30 | 44 |
| OK | 42 | 22 | 44 | 32 | 24 | 53 |
| OR | 78 | 13 | 9.8 | 72 | 18 | 14 |
| PA | 54 | 27 | 24 | 42 | 34 | 32 |
| RI | 47 | 32 | 30 | 41 | 34 | 34 |
| SC | 35 | 32 | 41 | 37 | 27 | 44 |
| SD | 27 | 17 | 65 | 24 | 17 | 67 |
| TN | 39 | 23 | 46 | 38 | 25 | 47 |
| TX | 45 | 13 | 48 | 43 | 14 | 50 |
| UT | 25 | 33 | 57 | 24 | 29 | 60 |
| VA | 62 | 23 | 18 | 63 | 21 | 19 |
| VT | 25 | 48 | 40 | 22 | 52 | 41 |
| WA | 72 | 18 | 11 | 64 | 25 | 14 |
| WI | 51 | 23 | 33 | 38 | 27 | 43 |
| WV | 49 | 36 | 20 | 44 | 42 | 20 |
| WY | 20 | 17 | 71 | 18 | 16 | 75 |

Individuals with missing data excluded from calculations

**Table S4.** Adjusted and unadjusted odds ratio (OR) of COVID-19 among those engaged in full- and part-time in-person schooling versus those engaged in virtual or homeschooling outcomes. All analyses adjust for survey weights. See methods for adjustment factors.

| Schooling | Loss of taste/smell | | | | COVID like illness | | | | Test+ | | | |
|---|---|---|---|---|---|---|---|---|---|---|---|---|
| | adj. OR | 95% CI | OR | 95% CI | adj OR | 95% CI | OR | 95% CI | adj OR | 95% CI | OR | 95% CI |
| Overall | | | | | | | | | | | | |
| Full time in person | 1.21 | 1.16–1.27 | 1.46 | 1.41–1.52 | 1.38 | 1.30–1.47 | 2.21 | 2.10–2.32 | 1.30 | 1.24–1.35 | 1.07 | 1.03–1.11 |
| Part time in person | 1.18 | 1.13–1.24 | 1.37 | 1.32–1.43 | 1.21 | 1.13–1.29 | 1.91 | 1.81–2.01 | 1.09 | 1.03–1.14 | 0.96 | 0.92–1.00 |
| Grades K or under | | | | | | | | | | | | |
| Full time in person | 1.11 | 0.97–1.27 | 0.93 | 0.83–1.04 | 1.16 | 0.95–1.42 | 1.15 | 0.96–1.38 | 1.21 | 1.06–1.38 | 0.87 | 0.78–0.98 |
| Part time in person | 0.95 | 0.80–1.12 | 0.84 | 0.72–0.98 | 0.90 | 0.70–1.16 | 0.84 | 0.66–1.07 | 1.06 | 0.90–1.25 | 0.80 | 0.69–0.93 |
| Grades 1 to 5 | | | | | | | | | | | | |
| Full time in person | 1.19 | 1.07–1.33 | 1.10 | 1.00–1.21 | 1.12 | 0.93–1.35 | 1.13 | 0.97–1.31 | 1.31 | 1.17–1.46 | 0.95 | 0.86–1.05 |
| Part time in person | 1.15 | 1.01–1.32 | 1.06 | 0.93–1.20 | 1.34 | 1.10–1.62 | 1.33 | 1.11–1.59 | 1.01 | 0.88–1.16 | 0.83 | 0.72–0.94 |
| Grades 6 to 8 | | | | | | | | | | | | |
| Full time in person | 1.30 | 1.12–1.52 | 1.20 | 1.05–1.37 | 1.42 | 1.14–1.78 | 1.41 | 1.16–1.72 | 1.38 | 1.19–1.61 | 1.14 | 1.00–1.30 |
| Part time in person | 1.24 | 1.04–1.48 | 1.09 | 0.93–1.28 | 1.43 | 1.10–1.87 | 1.30 | 1.03–1.65 | 1.05 | 0.88–1.27 | 0.88 | 0.74–1.03 |
| Grades 9 to 12 | | | | | | | | | | | | |
| Full time in person | 1.36 | 1.20–1.53 | 1.31 | 1.18–1.46 | 1.64 | 1.39–1.93 | 1.85 | 1.60–2.15 | 1.53 | 1.37–1.72 | 1.19 | 1.07–1.31 |
| Part time in person | 1.12 | 0.98–1.27 | 0.96 | 0.86–1.08 | 1.00 | 0.84–1.19 | 1.01 | 0.86–1.19 | 1.10 | 0.98–1.24 | 0.87 | 0.79–0.97 |

**Table S5.** Survey-weighted percentages of participants reporting school mitigation measures among those with ≥1 school-aged child in the household attending any in-person school (part-time or full-time) by state.

| State | Student masking | Teacher masking | Same teacher | Same students | Outdoor instruction | Restricted entry | Reduced class size | Closed cafeteria | Closed play | Desk shields | Extra space | No extra curriculars | No supply sharing | Daily symptom screen |
|---|---|---|---|---|---|---|---|---|---|---|---|---|---|---|
| **Overall** | 87 | 76 | 37 | 50 | 12 | 61 | 48 | 29 | 20 | 20 | 60 | 32 | 56 | 46 |
| **AK** | 83 | 63 | 43 | 49 | 15 | 51 | 42 | 33 | 19 | 22 | 48 | 32 | 46 | 36 |
| **AL** | 86 | 73 | 35 | 46 | 12 | 54 | 36 | 33 | 16 | 21 | 55 | 20 | 52 | 37 |
| **AR** | 84 | 74 | 30 | 41 | 10 | 58 | 34 | 17 | 11 | 19 | 55 | 17 | 50 | 39 |
| **AZ** | 90 | 75 | 36 | 47 | 12 | 56 | 44 | 21 | 18 | 15 | 53 | 33 | 56 | 40 |
| **CA** | 84 | 71 | 52 | 57 | 24 | 61 | 62 | 41 | 35 | 27 | 59 | 47 | 57 | 58 |
| **CO** | 86 | 78 | 42 | 60 | 17 | 65 | 46 | 32 | 17 | 15 | 59 | 43 | 58 | 55 |
| **CT** | 96 | 85 | 40 | 59 | 20 | 72 | 61 | 39 | 25 | 30 | 71 | 52 | 68 | 34 |
| **DC** | 87 | 55 | 45 | 51 | 25 | 52 | 53 | 36 | 24 | 19 | 48 | 43 | 46 | 52 |
| **DE** | 88 | 80 | 44 | 55 | 18 | 64 | 67 | 40 | 28 | 18 | 68 | 39 | 63 | 50 |
| **FL** | 86 | 70 | 31 | 41 | 11 | 54 | 38 | 16 | 15 | 23 | 53 | 23 | 49 | 37 |
| **GA** | 75 | 65 | 32 | 45 | 11 | 56 | 39 | 29 | 16 | 16 | 52 | 23 | 51 | 39 |
| **HI** | 89 | 81 | 55 | 65 | 31 | 67 | 72 | 43 | 39 | 37 | 76 | 56 | 69 | 63 |
| **IA** | 92 | 79 | 30 | 44 | 7.6 | 58 | 33 | 17 | 9.1 | 18 | 57 | 19 | 51 | 22 |
| **ID** | 84 | 71 | 33 | 42 | 7.7 | 47 | 39 | 16 | 11 | 14 | 52 | 25 | 46 | 24 |
| **IL** | 95 | 84 | 43 | 55 | 15 | 68 | 60 | 43 | 31 | 19 | 69 | 51 | 65 | 69 |
| **IN** | 89 | 78 | 34 | 46 | 8.3 | 61 | 36 | 19 | 13 | 17 | 62 | 22 | 56 | 28 |
| **KS** | 90 | 80 | 36 | 51 | 11 | 65 | 39 | 24 | 12 | 17 | 62 | 20 | 57 | 55 |
| **KY** | 85 | 78 | 44 | 54 | 14 | 62 | 62 | 32 | 26 | 19 | 65 | 31 | 59 | 63 |
| **LA** | 82 | 72 | 32 | 49 | 11 | 53 | 38 | 36 | 22 | 17 | 54 | 26 | 53 | 55 |
| **MA** | 93 | 86 | 43 | 63 | 20 | 72 | 77 | 44 | 29 | 17 | 78 | 48 | 72 | 37 |
| **MD** | 85 | 78 | 48 | 61 | 25 | 70 | 64 | 41 | 23 | 18 | 58 | 44 | 57 | 66 |
| **ME** | 95 | 87 | 42 | 60 | 23 | 77 | 70 | 39 | 20 | 21 | 77 | 50 | 70 | 54 |
| **MI** | 91 | 81 | 41 | 53 | 12 | 67 | 46 | 35 | 16 | 19 | 59 | 40 | 59 | 45 |
| **MN** | 86 | 78 | 43 | 58 | 12 | 61 | 47 | 28 | 14 | 14 | 59 | 28 | 55 | 40 |
| **MO** | 82 | 75 | 33 | 48 | 9.5 | 61 | 35 | 25 | 11 | 17 | 56 | 20 | 53 | 37 |
| **MS** | 89 | 73 | 32 | 45 | 8.7 | 53 | 36 | 35 | 17 | 18 | 54 | 20 | 53 | 51 |
| **MT** | 90 | 78 | 38 | 52 | 12 | 58 | 42 | 30 | 13 | 16 | 60 | 23 | 54 | 33 |
| **NC** | 88 | 80 | 43 | 58 | 15 | 67 | 60 | 46 | 24 | 15 | 65 | 40 | 60 | 71 |
| **ND** | 91 | 76 | 30 | 42 | 8.2 | 55 | 28 | 13 | 7.3 | 13 | 54 | 17 | 51 | 25 |

| | | | | | | | | | | | | | | |
|---|---|---|---|---|---|---|---|---|---|---|---|---|---|---|
| **NE** | 93 | 81 | 31 | 43 | 8.5 | 58 | 26 | 18 | 11 | 15 | 54 | 20 | 50 | 40 |
| **NH** | 89 | 84 | 42 | 62 | 22 | 69 | 64 | 39 | 20 | 24 | 71 | 37 | 67 | 62 |
| **NJ** | 93 | 85 | 42 | 58 | 15 | 74 | 72 | 59 | 38 | 31 | 72 | 50 | 70 | 65 |
| **NM** | 90 | 70 | 53 | 61 | 19 | 59 | 59 | 33 | 26 | 19 | 54 | 44 | 53 | 56 |
| **NV** | 88 | 78 | 44 | 52 | 13 | 60 | 59 | 35 | 33 | 16 | 62 | 47 | 58 | 41 |
| **NY** | 92 | 82 | 42 | 58 | 15 | 70 | 70 | 42 | 32 | 26 | 74 | 51 | 67 | 64 |
| **OH** | 91 | 82 | 32 | 47 | 9.3 | 61 | 49 | 21 | 18 | 27 | 65 | 22 | 60 | 42 |
| **OK** | 80 | 69 | 32 | 41 | 7.6 | 56 | 30 | 16 | 10 | 13 | 49 | 17 | 47 | 39 |
| **OR** | 80 | 72 | 56 | 63 | 15 | 62 | 63 | 41 | 28 | 17 | 57 | 49 | 57 | 54 |
| **PA** | 94 | 82 | 35 | 50 | 9.8 | 62 | 56 | 25 | 23 | 20 | 68 | 34 | 62 | 44 |
| **RI** | 89 | 81 | 45 | 64 | 19 | 69 | 63 | 45 | 30 | 22 | 68 | 49 | 67 | 60 |
| **SC** | 87 | 77 | 38 | 50 | 12 | 61 | 52 | 38 | 22 | 41 | 62 | 29 | 58 | 38 |
| **SD** | 75 | 62 | 27 | 41 | 6.6 | 53 | 16 | 9.3 | 7 | 22 | 46 | 9.9 | 41 | 28 |
| **TN** | 77 | 68 | 31 | 46 | 10 | 58 | 36 | 23 | 15 | 14 | 54 | 22 | 52 | 52 |
| **TX** | 86 | 75 | 31 | 43 | 12 | 61 | 39 | 21 | 18 | 27 | 54 | 23 | 52 | 43 |
| **UT** | 95 | 80 | 34 | 41 | 6.7 | 42 | 26 | 13 | 9.4 | 11 | 48 | 31 | 49 | 21 |
| **VA** | 88 | 80 | 47 | 59 | 16 | 66 | 66 | 46 | 27 | 20 | 69 | 44 | 64 | 60 |
| **VT** | 93 | 90 | 51 | 72 | 39 | 84 | 60 | 61 | 26 | 24 | 76 | 49 | 70 | 86 |
| **WA** | 86 | 78 | 57 | 64 | 17 | 64 | 67 | 44 | 29 | 18 | 64 | 53 | 60 | 69 |
| **WI** | 93 | 82 | 37 | 54 | 12 | 65 | 47 | 28 | 14 | 19 | 66 | 29 | 59 | 33 |
| **WV** | 88 | 78 | 40 | 51 | 8.8 | 59 | 58 | 32 | 24 | 22 | 63 | 44 | 59 | 38 |
| **WY** | 88 | 76 | 27 | 37 | 7.6 | 51 | 23 | 17 | 6.9 | 21 | 55 | 13 | 45 | 41 |

**Table S6.** Survey-weighted percentage of participants with ≥1 school-aged child attending any in-person school (part-time or full-time) by the reported number of school mitigation measures and state.

| | Number of school mitigation measures reported | | | | |
|---|---|---|---|---|---|
| **State** | **0** | **1-3** | **4-6** | **7-9** | **10+** |
| **AK** | 3.9% | 36% | 18% | 24% | 18% |
| **AL** | 1.9% | 27% | 29% | 28% | 15% |
| **AR** | 2.0% | 29% | 32% | 26% | 10% |
| **AZ** | 1.6% | 27% | 26% | 29% | 16% |
| **CA** | 2.7% | 25% | 13% | 22% | 38% |
| **CO** | 1.5% | 21% | 21% | 32% | 24% |
| **CT** | 1.2% | 14% | 18% | 36% | 31% |
| **DC** | 5.6% | 44% | 5.4% | 13% | 32% |
| **DE** | 2.1% | 19% | 16% | 33% | 30% |
| **FL** | 2.3% | 30% | 29% | 26% | 13% |
| **GA** | 2.6% | 30% | 29% | 25% | 14% |
| **HI** | 0.6% | 16% | 9.7% | 30% | 43% |
| **IA** | 1.3% | 28% | 35% | 27% | 8.4% |
| **ID** | 2.5% | 33% | 31% | 24% | 9.5% |
| **IL** | 1.1% | 15% | 17% | 33% | 34% |
| **IN** | 1.2% | 25% | 34% | 30% | 10% |
| **KS** | 1.2% | 20% | 30% | 35% | 14% |
| **KY** | 1.4% | 20% | 20% | 34% | 25% |
| **LA** | 2.3% | 26% | 26% | 29% | 17% |
| **MA** | 1.0% | 12% | 16% | 37% | 34% |
| **MD** | 1.7% | 19% | 14% | 31% | 34% |
| **ME** | 0.6% | 12% | 16% | 37% | 34% |
| **MI** | 1.1% | 20% | 25% | 32% | 22% |
| **MN** | 1.6% | 22% | 26% | 34% | 16% |
| **MO** | 2.1% | 26% | 33% | 28% | 11% |
| **MS** | 2.1% | 26% | 28% | 29% | 14% |
| **MT** | 1.6% | 24% | 30% | 31% | 14% |
| **NC** | 1.4% | 17% | 18% | 32% | 31% |
| **ND** | 1.6% | 31% | 35% | 25% | 6.7% |
| **NE** | 1.4% | 27% | 36% | 26% | 9.9% |
| **NH** | 0.3% | 13% | 22% | 33% | 31% |
| **NJ** | 1.3% | 14% | 11% | 32% | 42% |
| **NM** | 1.5% | 28% | 13% | 22% | 35% |
| **NV** | 1.8% | 21% | 19% | 31% | 27% |
| **NY** | 1.3% | 15% | 13% | 32% | 38% |
| **OH** | 1.0% | 21% | 29% | 32% | 16% |
| **OK** | 3.1% | 32% | 33% | 23% | 8.8% |
| **OR** | 1.9% | 23% | 16% | 25% | 34% |
| **PA** | 1.1% | 19% | 25% | 35% | 20% |
| **RI** | 1.9% | 15% | 16% | 30% | 37% |

| | | | | | |
|---|---|---|---|---|---|
| **SC** | 1.3% | 20% | 26% | 31% | 22% |
| **SD** | 3.3% | 38% | 34% | 21% | 4.4% |
| **TN** | 2.3% | 28% | 30% | 27% | 12% |
| **TX** | 2.2% | 28% | 28% | 26% | 16% |
| **UT** | 1.1% | 32% | 35% | 24% | 7.6% |
| **VA** | 1.2% | 17% | 16% | 33% | 33% |
| **VT** | 0.7% | 9.0% | 9.5% | 32% | 49% |
| **WA** | 1.6% | 20% | 13% | 26% | 40% |
| **WI** | 1.3% | 20% | 29% | 32% | 18% |
| **WV** | 1.7% | 20% | 22% | 33% | 23% |
| **WY** | 1.3% | 31% | 36% | 25% | 6.5% |

Individuals with missing data excluded from calculations

**Table S7.** Association between risk of full-time schooling and number of mitigation measures implemented after accounting for survey design and adjusting for county and individual level covariates.

| | Loss of taste/smell | | COVID like illness | | Test+ | |
|---|---|---|---|---|---|---|
| **Schooling** | **adj. OR** | **95% CI** | **adj OR** | **95% CI** | **adj OR** | **95% CI** |
| Overall | | | | | | |
| N mitigation measures | 0.92 | 0.91–0.93 | 0.91 | 0.89–0.92 | 0.93 | 0.92–0.94 |
| Grades K or under | | | | | | |
| N mitigation measures | 0.95 | 0.92–0.97 | 0.91 | 0.87–0.95 | 0.94 | 0.92–0.97 |
| Grades 1 to 5 | | | | | | |
| N mitigation measures | 0.92 | 0.90–0.94 | 0.92 | 0.89–0.96 | 0.94 | 0.91–0.96 |
| Grades 6 to 8 | | | | | | |
| N mitigation measures | 0.92 | 0.89–0.95 | 0.90 | 0.86–0.95 | 0.93 | 0.90–0.97 |
| Grades 9 to 12 | | | | | | |
| N mitigation measures | 0.90 | 0.87–0.93 | 0.90 | 0.87–0.94 | 0.93 | 0.90–0.96 |

**Table S8.** The association between mitigation measures and the off-ratio of COVID-19-related outcomes in those engaged in in-person schooling compared to those engaged in virtual- or home-schooling. Adjusted form county and individual level covariates.

| | Loss of taste/smell | | COVID like illness | | Test+ | |
|---|---|---|---|---|---|---|
| **Mitigatoin** | **adj. OR** | **95% CI** | **adj OR** | **95% CI** | **adj OR** | **95% CI** |
| student mask mandate | 0.92 | 0.84–1.01 | 0.89 | 0.79–1.01 | 0.91 | 0.83–1.00 |
| teacher mask mandate | 0.82 | 0.75–0.89 | 0.66 | 0.59–0.74 | 0.91 | 0.83–1.00 |
| same teacher all day | 1.05 | 0.97–1.14 | 0.94 | 0.84–1.06 | 1.00 | 0.93–1.08 |
| same students all day | 0.88 | 0.81–0.95 | 0.86 | 0.77–0.97 | 0.93 | 0.86–1.00 |
| outdoor instruction | 0.89 | 0.81–0.98 | 1.02 | 0.89–1.18 | 0.88 | 0.80–0.98 |
| restricted entry | 0.94 | 0.87–1.02 | 0.91 | 0.82–1.02 | 0.88 | 0.81–0.95 |
| reduced class size | 0.95 | 0.89–1.03 | 0.94 | 0.85–1.04 | 1.01 | 0.94–1.09 |
| closed cafeteria | 1.00 | 0.93–1.08 | 1.03 | 0.92–1.16 | 1.03 | 0.95–1.11 |
| closed playground | 1.01 | 0.93–1.10 | 1.13 | 0.99–1.28 | 1.01 | 0.92–1.10 |
| desk shields | 1.07 | 0.99–1.15 | 1.29 | 1.15–1.44 | 1.12 | 1.04–1.22 |
| extra desk space | 0.92 | 0.85–0.99 | 0.93 | 0.84–1.04 | 0.96 | 0.89–1.04 |
| no extracurriculars | 0.84 | 0.78–0.91 | 0.86 | 0.77–0.96 | 0.73 | 0.68–0.79 |
| no sharing supplies | 0.89 | 0.82–0.96 | 0.90 | 0.81–1.01 | 0.92 | 0.85–1.00 |
| daily symptom screen | 0.82 | 0.76–0.87 | 0.75 | 0.67–0.82 | 0.78 | 0.73–0.84 |
| partime in-person | 1.11 | 1.04–1.18 | 1.08 | 0.99–1.18 | 0.97 | 0.91–1.03 |

**Table S9.** Selected demographics among participants with ≥1 school-aged child in the household attending any in-person schooling (part-time or full-time) by number of reported school mitigation measures; observed and survey-weighted percentages reported.

| | Number of school mitigation measures reported | | | | | | | | | |
|---|---|---|---|---|---|---|---|---|---|---|
| | **0** | | **1-3** | | **4-6** | | **7-9** | | **10+** | |
| | N=3,844 | | N=53,309 | | N=69,284 | | N=87,242 | | N=63,951 | |
| **Gender** | **N (%)** | **Weighted %** | **N (%)** | **Weighted %** | **N (%)** | **Weighted %** | **N (%)** | **Weighted %** | **N (%)** | **Weighted %** |
| Female | 2,136 (56%) | 39% | 30,886 (58%) | 42% | 49,730 (72%) | 57% | 65,534 (75%) | 61% | 48,202 (75%) | 62% |
| Male | 1,453 (38%) | 53% | 19,687 (37%) | 52% | 18,475 (27%) | 41% | 20,638 (24%) | 37% | 14,926 (23%) | 37% |
| Non-binary | 69 (1.8%) | 2.1% | 594 (1.1%) | 1.4% | 216 (0.3%) | 0.5% | 208 (0.2%) | 0.3% | 182 (0.3%) | 0.4% |
| Other | 183 (4.8%) | 5.8% | 2,105 (3.9%) | 5.0% | 846 (1.2%) | 1.5% | 839 (1.0%) | 1.2% | 624 (1.0%) | 1.2% |
| **Age (years)** | | | | | | | | | | |
| 18 to 24 | 163 (4.2%) | 8.4% | 3,808 (7.1%) | 14% | 2,810 (4.1%) | 8.4% | 2,270 (2.6%) | 5.6% | 1,095 (1.7%) | 3.6% |
| 25 to 34 | 727 (19%) | 19% | 9,203 (17%) | 18% | 12,362 (18%) | 18% | 16,151 (19%) | 18% | 12,531 (20%) | 20% |
| 35 to 44 | 1,278 (33%) | 32% | 16,994 (32%) | 29% | 26,780 (39%) | 35% | 36,621 (42%) | 39% | 28,802 (45%) | 42% |
| 45 to 54 | 925 (24%) | 24% | 12,610 (24%) | 22% | 18,267 (26%) | 26% | 21,718 (25%) | 26% | 14,034 (22%) | 23% |
| 55 to 65 | 439 (11%) | 9.9% | 6,220 (12%) | 9.8% | 5,760 (8.3%) | 7.7% | 6,642 (7.6%) | 7.3% | 4,490 (7.0%) | 6.7% |
| 65+ | 300 (7.8%) | 6.8% | 4,370 (8.2%) | 6.7% | 3,255 (4.7%) | 4.2% | 3,787 (4.3%) | 4.0% | 2,961 (4.6%) | 4.4% |
| **Educational Level** | | | | | | | | | | |
| High school | 924 (24%) | 13% | 12,439 (23%) | 9.1% | 10,022 (14%) | 3.9% | 10,944 (13%) | 3.0% | 7,917 (12%) | 3.2% |
| Less than high school | 408 (11%) | 25% | 4,111 (7.7%) | 25% | 2,011 (2.9%) | 16% | 1,991 (2.3%) | 14% | 1,600 (2.5%) | 14% |
| Some college | 865 (23%) | 21% | 13,771 (26%) | 26% | 16,382 (24%) | 25% | 19,426 (22%) | 23% | 14,118 (22%) | 23% |
| College/Professional Degree | 1,064 (28%) | 26% | 16,461 (31%) | 28% | 28,921 (42%) | 39% | 37,811 (43%) | 41% | 27,297 (43%) | 41% |
| Graduate | 487 (13%) | 12% | 5,635 (11%) | 9.8% | 11,439 (17%) | 15% | 16,511 (19%) | 17% | 12,622 (20%) | 18% |
| **Occupation** | | | | | | | | | | |
| Office and admin support | 173 (4.5%) | 3.6% | 2,930 (5.5%) | 4.2% | 6,030 (8.7%) | 7.5% | 8,148 (9.3%) | 8.3% | 5,513 (8.6%) | 7.7% |
| Comm/social service | 82 (2.1%) | 2.0% | 1,018 (1.9%) | 1.6% | 2,096 (3.0%) | 2.5% | 2,822 (3.2%) | 2.8% | 2,070 (3.2%) | 2.7% |
| Education | 191 (5.0%) | 3.9% | 2,646 (5.0%) | 3.7% | 7,050 (10%) | 8.2% | 9,208 (11%) | 8.8% | 5,724 (9.0%) | 7.5% |
| Food service | 150 (3.9%) | 4.3% | 2,111 (4.0%) | 4.4% | 2,161 (3.1%) | 3.5% | 2,418 (2.8%) | 3.2% | 1,591 (2.5%) | 2.8% |
| Healthcare | 364 (9.5%) | 7.6% | 5,221 (9.8%) | 8.2% | 9,189 (13%) | 12% | 11,902 (14%) | 12% | 8,428 (13%) | 12% |
| Production | 116 (3.0%) | 3.5% | 1,814 (3.4%) | 3.9% | 1,743 (2.5%) | 3.2% | 1,723 (2.0%) | 2.4% | 1,134 (1.8%) | 2.3% |
| Sales | 188 (4.9%) | 4.9% | 3,183 (6.0%) | 6.2% | 4,398 (6.3%) | 6.9% | 5,383 (6.2%) | 6.7% | 3,688 (5.8%) | 6.1% |
| Transportation/delivery | 93 (2.4%) | 3.1% | 1,553 (2.9%) | 3.7% | 1,455 (2.1%) | 2.8% | 1,497 (1.7%) | 2.3% | 1,046 (1.6%) | 2.2% |
| Not Employed | 1,418 (37%) | 36% | 18,249 (34%) | 33% | 17,877 (26%) | 25% | 22,846 (26%) | 26% | 18,407 (29%) | 29% |
| Other | 860 (22%) | 25% | 12,907 (24%) | 28% | 16,154 (23%) | 26% | 20,032 (23%) | 26% | 15,379 (24%) | 27% |

| | | | | | | | | | | |
|---|---|---|---|---|---|---|---|---|---|---|
| **Description** | | | | | | | | | | |
| Metro - Counties in metro areas of ≥1 million | 1,274 (33%) | 36% | 16,968 (32%) | 35% | 22,961 (33%) | 35% | 33,736 (39%) | 41% | 28,323 (44%) | 47% |
| Metro - Counties in metro areas of 250,000 to 1 million | 950 (25%) | 24% | 14,170 (27%) | 26% | 19,780 (29%) | 28% | 25,015 (29%) | 28% | 17,881 (28%) | 27% |
| Metro - Counties in metro areas of <250,000 | 476 (12%) | 12% | 7,310 (14%) | 13% | 9,685 (14%) | 14% | 10,861 (12%) | 12% | 6,951 (11%) | 10% |
| Nonmetro - Completely rural or <2,500 urban population, adjacent to a metro area | 45 (1.2%) | 0.9% | 499 (0.9%) | 0.9% | 621 (0.9%) | 0.8% | 612 (0.7%) | 0.7% | 401 (0.6%) | 0.6% |
| Nonmetro - Completely rural or < 2,500 urban population, not adjacent to a metro area | 63 (1.6%) | 1.4% | 769 (1.4%) | 1.2% | 852 (1.2%) | 1.1% | 753 (0.9%) | 0.7% | 355 (0.6%) | 0.5% |
| Nonmetro - Urban population of 2,500 to 19,999, adjacent to a metro area | 300 (7.8%) | 7.0% | 4,140 (7.8%) | 7.4% | 5,021 (7.2%) | 7.0% | 4,712 (5.4%) | 5.3% | 2,570 (4.0%) | 3.8% |
| Nonmetro - Urban population of 2,500 to 19,999, not adjacent to a metro area | 175 (4.6%) | 3.9% | 2,517 (4.7%) | 4.2% | 2,941 (4.2%) | 3.9% | 2,939 (3.4%) | 3.0% | 1,644 (2.6%) | 2.3% |
| Nonmetro - Urban population of ≥20,000 or more, adjacent to a metro area | 239 (6.2%) | 6.0% | 3,221 (6.0%) | 5.8% | 4,194 (6.1%) | 5.9% | 5,118 (5.9%) | 5.6% | 3,440 (5.4%) | 5.1% |
| Nonmetro - Urban population of 20,000 or more, not adjacent to a metro area | 97 (2.5%) | 2.1% | 1,436 (2.7%) | 2.2% | 1,710 (2.5%) | 2.1% | 1,816 (2.1%) | 1.9% | 1,155 (1.8%) | 1.7% |
| **Household size (persons)** | | | | | | | | | | |
| 2 | 164 (4.3%) | 3.8% | 2,374 (4.5%) | 3.9% | 3,320 (4.8%) | 4.4% | 3,867 (4.4%) | 4.2% | 2,601 (4.1%) | 3.9% |
| 3 | 723 (19%) | 17% | 9,896 (19%) | 17% | 15,536 (22%) | 21% | 18,400 (21%) | 21% | 12,215 (19%) | 19% |
| 4 | 957 (25%) | 25% | 14,622 (27%) | 27% | 23,539 (34%) | 34% | 31,344 (36%) | 35% | 22,967 (36%) | 35% |
| 5 | 679 (18%) | 17% | 10,209 (19%) | 20% | 13,404 (19%) | 20% | 17,338 (20%) | 20% | 13,291 (21%) | 21% |
| 6 | 476 (12%) | 13% | 6,333 (12%) | 12% | 6,746 (9.7%) | 10% | 8,454 (9.7%) | 10% | 6,596 (10%) | 11% |
| 7 | 262 (6.8%) | 7.3% | 3,313 (6.2%) | 6.6% | 2,935 (4.2%) | 4.7% | 3,650 (4.2%) | 4.5% | 2,861 (4.5%) | 4.8% |
| 8 | 137 (3.6%) | 3.7% | 1,863 (3.5%) | 3.7% | 1,456 (2.1%) | 2.3% | 1,663 (1.9%) | 2.0% | 1,387 (2.2%) | 2.4% |
| 9 | 74 (1.9%) | 2.0% | 1,017 (1.9%) | 2.1% | 686 (1.0%) | 1.1% | 773 (0.9%) | 0.9% | 598 (0.9%) | 1.0% |
| 10+ | 316 (8.2%) | 9.7% | 3,073 (5.8%) | 6.7% | 1,219 (1.8%) | 2.1% | 1,253 (1.4%) | 1.6% | 1,063 (1.7%) | 1.9% |

Missing responses not shown, but included in percentage calculations.

**Table S10.** Selected behaviors relevant to COVID-19 acquisition/transmission among participants with ≥1 school-aged child in the household attending any in-person schooling (part-time or full-time) by number of reported school mitigation measures; observed and survey-weighted percentages reported.

| | Number of school mitigation measures reported | | | | | | | | | |
|---|---|---|---|---|---|---|---|---|---|---|
| | 0 | | 1-3 | | 4-6 | | 7-9 | | 10+ | |
| | N=3,844 | | N=53,309 | | N=69,284 | | N=87,242 | | N=63,951 | |
| | N (%) | Weighted % | N (%) | Weighted % | N (%) | Weighted % | N (%) | Weighted % | N (%) | Weighted % |
| **Mask use in public spaces** | | | | | | | | | | |
| Never in public spaces | 125 (3.3%) | 2.9% | 2,040 (3.8%) | 3.4% | 2,027 (2.9%) | 2.6% | 2,423 (2.8%) | 2.6% | 1,886 (2.9%) | 2.8% |
| Always use | 2,204 (57%) | 53% | 30,656 (58%) | 54% | 46,588 (67%) | 64% | 65,435 (75%) | 72% | 51,121 (80%) | 78% |
| Mostly use | 376 (9.8%) | 9.9% | 6,789 (13%) | 13% | 9,638 (14%) | 15% | 10,180 (12%) | 12% | 5,815 (9.1%) | 9.8% |
| Sometimes use | 193 (5.0%) | 5.2% | 2,888 (5.4%) | 5.7% | 3,153 (4.6%) | 5.2% | 2,439 (2.8%) | 3.2% | 1,097 (1.7%) | 2.1% |
| Rarely use | 231 (6.0%) | 6.9% | 3,086 (5.8%) | 6.5% | 2,507 (3.6%) | 4.4% | 1,606 (1.8%) | 2.3% | 728 (1.1%) | 1.4% |
| Never use | 543 (14%) | 17% | 5,428 (10%) | 12% | 2,377 (3.4%) | 4.4% | 1,225 (1.4%) | 1.8% | 616 (1.0%) | 1.3% |
| **Travel out of state in last five days** | 610 (16%) | 19% | 6,682 (13%) | 14% | 5,864 (8.5%) | 9.5% | 6,248 (7.2%) | 7.8% | 4,278 (6.7%) | 7.3% |
| **Went to bar/restaurant/café in last 24 hours** | 1,001 (26%) | 29% | 13,778 (26%) | 29% | 14,324 (21%) | 23% | 14,154 (16%) | 18% | 8,570 (13%) | 15% |
| **Attended an event with ≥10 people in last 24 hours** | 918 (24%) | 27% | 11,128 (21%) | 24% | 9,962 (14%) | 17% | 8,725 (10%) | 11% | 4,797 (7.5%) | 8.6% |
| **Used public transit last 24 hours** | 416 (11%) | 13% | 3,466 (6.5%) | 8.2% | 1,324 (1.9%) | 2.60% | 1,293 (1.5%) | 1.9% | 1,101 (1.7%) | 2.3% |

Missing responses not shown, but included in percentage calculations.

**Table S11.** The relative odds, compared to office and administrative support staff not working outside the home for pay, of COVID-19-related outcomes among individuals who do, and do not, report paid work outside the home.

| | Loss of taste/smell | | | | COVID like illness | | | | Test+ | | | |
|---|---|---|---|---|---|---|---|---|---|---|---|---|
| **Occupation** | **adj. OR** | **OR** | **95% CI** | **95% CI** | **adj OR** | **OR** | **95% CI** | **95% CI** | **adj OR** | **OR** | **95% CI** | **95% CI** |
| Food service | 2.18 | 3.08 | 1.62–2.93 | 2.37–4.00 | 2.20 | 4.15 | 1.44–3.35 | 2.89–5.96 | 2.37 | 2.28 | 1.77–3.19 | 1.74–2.98 |
| Healthcare | 1.25 | 1.40 | 1.10–1.42 | 1.24–1.59 | 1.28 | 1.61 | 1.05–1.57 | 1.33–1.95 | 1.25 | 1.30 | 1.10–1.42 | 1.15–1.47 |
| K12Teacher | 0.96 | 1.09 | 0.80–1.15 | 0.92–1.29 | 0.85 | 1.21 | 0.64–1.14 | 0.92–1.58 | 1.02 | 1.10 | 0.85–1.22 | 0.93–1.30 |
| Other | 0.98 | 0.98 | 0.89–1.07 | 0.90–1.07 | 1.00 | 1.10 | 0.87–1.15 | 0.96–1.26 | 0.98 | 0.95 | 0.89–1.07 | 0.88–1.04 |
| OtherEductation | 0.79 | 0.75 | 0.67–0.93 | 0.64–0.88 | 0.63 | 0.72 | 0.49–0.82 | 0.57–0.91 | 0.78 | 0.74 | 0.67–0.91 | 0.64–0.86 |
| Sales | 1.33 | 1.23 | 1.16–1.53 | 1.08–1.40 | 1.23 | 1.24 | 1.00–1.52 | 1.02–1.51 | 1.30 | 1.21 | 1.14–1.49 | 1.06–1.37 |
| Office and admin support * work_outside_paid | 1.31 | 1.35 | 1.19–1.44 | 1.24–1.48 | 1.29 | 1.49 | 1.12–1.50 | 1.29–1.71 | 1.59 | 1.40 | 1.45–1.74 | 1.28–1.52 |
| Food service * work_outside_paid | 0.85 | 0.80 | 0.64–1.14 | 0.62–1.03 | 0.85 | 0.76 | 0.56–1.29 | 0.54–1.08 | 0.77 | 0.71 | 0.57–1.03 | 0.54–0.92 |
| Healthcare * work_outside_paid | 1.25 | 1.24 | 1.12–1.39 | 1.12–1.38 | 1.22 | 1.35 | 1.03–1.45 | 1.15–1.58 | 1.69 | 1.37 | 1.52–1.89 | 1.24–1.52 |
| K12Teacher * work_outside_paid | 1.38 | 1.18 | 1.15–1.65 | 0.99–1.40 | 1.50 | 1.25 | 1.13–1.99 | 0.96–1.63 | 1.83 | 1.35 | 1.54–2.19 | 1.14–1.59 |
| Other * work_outside_paid | 1.44 | 1.85 | 1.36–1.53 | 1.76–1.95 | 1.39 | 2.30 | 1.27–1.53 | 2.13–2.49 | 1.53 | 1.46 | 1.44–1.62 | 1.39–1.54 |
| OtherEductation * work_outside_paid | 1.43 | 1.53 | 1.20–1.69 | 1.30–1.80 | 1.41 | 1.59 | 1.08–1.84 | 1.24–2.03 | 1.79 | 1.53 | 1.54–2.09 | 1.32–1.77 |
| Sales * work_outside_paid | 1.12 | 1.38 | 0.99–1.27 | 1.23–1.55 | 1.28 | 1.72 | 1.06–1.54 | 1.44–2.05 | 1.24 | 1.18 | 1.09–1.41 | 1.05–1.33 |
